# Approaches to early intervention for common mental health problems in young people: a systematic review

**DOI:** 10.1101/2025.02.07.25321864

**Authors:** Rebecca Appleton, Phoebe Barnett, Connor Clarke, Jialin Yang, Sadiya Begum, Julian Edbrooke-Childs, Isobel Emptage, Una Foye, Jessica Griffiths, Isabel Hanson, Nima Cas Hunt, Ruby Jarvis, Maeve McAuliffe, Emma Maynard, Lizzie Mitchell, Irina Mostafa, Tamara Pemovska, Rob Saunders, Kylee Trevillion, Polly Waite, Brynmor Lloyd-Evans, Sonia Johnson

**Affiliations:** NIHR Policy Research Unit in Mental Health, Division of Psychiatry, University College London; Department of Applied Clinical Research and Epidemiology, Division of Psychiatry, University College London; Institute of Psychiatry, Psychology and Neuroscience, King’s College London; Anna Freud Centre, University College London; Mental Health Nursing Department, Institute of Psychiatry, Psychology and Neuroscience, King’s College London; NIHR Policy Research Unit in Mental Health, Institute of Psychiatry, Psychology and Neuroscience, King’s College London; Nuffield Department of Primary Care, University of Oxford; Florence Nightingale Faculty of Nursing Midwifery & Palliative Care, King’s College London; Centre for Evidence and Implementation, London, UK; CORE Data Lab, Centre for Outcomes Research and Effectiveness, Research Department of Clinical, Educational and Health Psychology, University College London; Department of Experimental Psychology, University of Oxford; North London NHS Foundation Trust, London

## Abstract

Effective early support for children and young people is a high priority. Early intervention approaches for young people with psychosis or eating disorders have substantial supporting evidence, but well-established approaches to delivering a prompt, effective response to young people presenting with early symptoms of anxiety and depression are lacking. We conducted a systematic review of outcomes of early interventions or approaches for young people (between 11 and 25 years) with initial symptoms of depression, anxiety and other common mental health difficulties. Five bibliographic and two grey literature databases were searched for papers relating to youth, early intervention and common mental health problems. We conducted a narrative synthesis of models and assessed quality using CASP checklists. We included 38 studies (43 publications): of these, randomised controlled trials were high quality but other studies tended to lack control groups and be of lower quality. Approaches broadly aimed provide a more comprehensive and effective early response to symptom onset, with primary goals falling into one of: 1) Making care more comprehensive and joined up, 2) Increasing speed or ease of access to support, or 3) Providing targeted support for specific needs in addition to anxiety and depression. Some evidence indicates that these approaches may facilitate access and reduce waiting times in the short-term, whilst decreasing burden on other mental health or emergency services. Significant improvements in mental health and wellbeing compared to controls were also reported across most studies with comparator groups, alongside high acceptability. Overall, models of early intervention for depression and anxiety show promise in improving access, experience and outcomes of care for young people. However, high heterogeneity of interventions and outcomes measured limits certainty. More robust controlled studies are needed, alongside comprehensive details of support received by young people through the intervention, and evidence of what works for whom in which settings.

## Introduction

Around two-thirds of mental health problems in adulthood have their onset between the ages of 14 and 25 (de Girolamo et al., 2019; Kessler et al., 2005; Solmi et al., 2022), and around one in five children and young people aged 8 to 25 years have a probable mental health disorder (Newlove-Delgado et al., 2022). Evidence suggests that rates of anxiety and depression are increasing in children and young people (Newlove-Delgado et al., 2022), particularly since the Covid-19 pandemic (Deighton et al., 2019; Goodwin et al., 2020; Racine et al., 2021; World Health Organization, 2022). This rise has also been more marked in young women compared to young men (Newlove-Delgado et al., 2022).

Early intervention models in mental health care have been designed to provide support at the onset of an individual’s symptoms, aiming to prevent problems from becoming more severe and long-term (Davey & McGorry, 2019). Such efforts are crucial as research indicates that depression or anxiety diagnoses during adolescence are associated with school difficulties, and poorer academic and socioeconomic outcomes, such as unemployment in adulthood (Clayborne et al., 2019; Essau et al., 2014; Pollard et al., 2023; Smith et al., 2021; van Poortvliet, 2024). In addition, poorer psychosocial outcomes such as low social support, loneliness, poorer family relationships, higher stress and alcohol/drug abuse have been identified in adults who experienced anxiety or depression in their youth (Clayborne et al., 2019; Essau et al., 2014; Pollard et al., 2023).

There has been an international commitment to increase capacity of services to ensure all young people can access timely support for their mental health (NHS, 2019; World Health Organization, 2024). However, it has been widely reported that existing services for children and young people are unable to meet the demand for mental health support (McGorry et al., 2022; National Audit Office, 2023; Rainer & Abdinasir, 2023) and young people are the age group least likely to receive support for their mental health (Babajide et al., 2020; Roche et al., 2020). There are also inequalities in accessing support, with young people from minoritised ethnic or LGBTQ+ groups experiencing further barriers (Edbrooke-Childs & Patalay, 2019; Green & Dorison, 2020). Thus, it appears that opportunities are often missed to intervene promptly at the onset of symptoms and prevent further deterioration in children’s and young people’s mental health and social outcomes (Knapp et al., 2016).

There is substantial evidence to suggest that early intervention models have potential to improve outcomes for illnesses including psychosis (Killackey & Yung, 2007; McGorry, 2015) and eating disorders (Koreshe et al., 2023). However, there has been less research identifying effective approaches to early intervention in young people following the onset of symptoms for common mental health conditions such as anxiety and depression (Malla et al., 2016).

This review aimed to establish what services and approaches to providing early intervention support have been tested to improve the prognosis for young people (between 11 and 25 years) presenting with early symptoms of one or more common mental health problems. By common mental health problems, we are referring to depression, any anxiety disorder (e.g. generalised anxiety disorder, obsessive compulsive disorder, social anxiety), post-traumatic stress disorder (PTSD), or general psychological distress (Kendrick & Pilling, 2012). While there are evidence-based treatments for depression and anxiety (NICE, 2019, 2022), the focus of our review is not to establish the effectiveness of single interventions for common mental health problems, but to understand the evidence base regarding service models with more than one component that are intended to facilitate the early provision of care for young people with common mental health problems, i.e. early intervention models. We therefore addressed the following questions:

Primary:

What services or approaches to delivering early intervention mental health care are effective in:

1. Improving access to, and reducing waiting times for mental health support for young people with symptoms of anxiety and depression and other common mental health difficulties?
2. Improving mental health symptoms for young people with recent onset of anxiety, depressive and other common mental health difficulties?
3. Improving social outcomes or functioning, wellbeing, or quality of life in young people showing early signs of anxiety, depressive and other common mental health difficulties?

Secondary:

4. How cost effective are services or approaches to early intervention for young people with early signs of anxiety, depressive and other common mental health difficulties?
5. What methods/approaches are used by effective models of early intervention for young people with early signs of anxiety, depressive and other common mental health difficulties?
6. How acceptable do young people with early signs of anxiety, depression and other common mental health difficulties find early intervention mental health services?

This study was conducted by the NIHR Policy Research Unit in Mental Health, which conducts research to inform mental health policy in the UK, to inform policy making in this area. The above research questions were agreed with policy makers from the Department of Health and Social Care.

## Materials and Methods

This review is reported according to PRISMA guidelines (Page et al., 2021) and has been registered with PROSPERO (CRD42024529019). We involved young people with lived experience (LM, SB, NH) in all stages of the review, including developing the protocol, screening papers for inclusion, data synthesis and interpretation of the findings.

### Search strategy

We searched five databases (MEDLINE, EMBASE, PsycINFO, CINAHL and Web of Science Core Collection) using both subject heading and textword searches from inception to the 31^st^ January 2024. We conducted an update of this search on the 4^th^ December 2024. We searched for research relating to:

1. children, young people, young adults, youth, adolescents
2. early intervention, improving access
3. anxiety, depression, common mental health problems

We also searched grey literature databases HMIC and PsycExtra and the websites of relevant charities or non-profit organisations (e.g. youth mental health charities) for non-peer reviewed evidence which met our inclusion criteria. Forward and backward citation searching of any included papers was conducted using ‘Citation chaser’ (Haddaway et al., 2022). Appendix 1 provides full details of the search strategy.

### Study selection

All references were imported into Endnote for de-duplication and Covidence for title and abstract and full text screening. All title and abstract and full text screening was conducted independently by two members of the research team (CC, RA, JY, SB, NH, IM, TP, RJ, KT, UF, JG, PB). Any disagreements were resolved through team discussion.

### Inclusion/exclusion criteria

All inclusion and exclusion criteria are shown below in Table 1:

**Table 1:** Inclusion criteria for the review.

|  | Inclusion | Exclusion |
| --- | --- | --- |
| Population | Young people between 11-25 years (or mean sample age between 11 and 25 years) seeking support with first onset common mental health problems such as symptoms of any anxiety disorder (e.g. generalised anxiety disorder, obsessive compulsive disorder, social anxiety), depression, PTSD, or general psychological distress. Symptoms were not required to meet diagnostic thresholds. | >10% young people presenting with symptoms of psychosis, bipolar disorder, schizophrenia or with neurodevelopmental disorders (autism/ADHD) or eating disorders as their primary problem or without a co-occurring common mental health problem.<br>Studies with a mixed sample (over 20% of the sample with diagnoses other than common mental health problems) with data on outcomes for those with common mental health problems not presented separately.<br>Mixed age group sample including over 25s or under 11s, without data for those in the 11-25 age range presented separately, or if the mean age was unspecified. |
| Intervention | Any services and approaches to care which aim to either improve access to mental health care, provide early intervention support, or both. This can include face-to-face or telemental health care. We defined early intervention as any approaches to improve access or engagement with services soon after onset of symptoms for young people | 'Treatment as usual' and evaluation of general mental health services for children and young people which did not have the aim of improving access to treatment or providing early intervention.<br>Digital interventions without human interaction e.g. apps.<br>Any models or interventions primarily aimed at primary prevention of mental |
|  | presenting with symptoms of an anxiety, depressive or other common mental health problem.<br>Eligible forms of support involved complex interventions (e.g. care delivered by more than one person or team, or multi-component interventions). For example, a youth counselling service was included if it had multiple elements e.g. peer support, outreach work, social interventions etc. and/or had more than one type of mental health professional working at the service. We were interested only in studies on approaches designed to be improvements on current standard mental health services in their locality. | health problems, or at treatment of youth with extensive experience of mental health services.<br>Stand-alone single-component interventions which did not form part of a more complex intervention with multiple components aiming to improve access or provide early intervention mental health support.<br>Studies of interventions developed to treat only cohorts with PTSD after a single specific traumatic event e.g. a certain natural disaster. |
| Setting | Community-based services (including social services and services based in the voluntary sector)<br>Outpatient services<br>Primary care-based services<br>Services delivered online, for example telemental health (providing they met the intervention inclusion criteria) | Educational settings<br>Inpatient settings<br>Forensic settings |
| Outcomes | To be included, studies were required to include self-reported, clinician-rated or parent/carer-rated outcomes for young people, for at least one of:<br>Measures of wait time, accessibility, or percentage of referrals accepted;<br>Anxiety or depression or general mental health symptom severity or diagnosis;<br>Social functioning, wellbeing, or quality of life outcomes; | Studies which only described an intervention or model of early intervention without reporting at least one outcome of interest.<br>Studies with acceptability or cost-effectiveness outcomes only. |
|  | Referrals to secondary mental health care or crisis/inpatient services;<br>Housing security, financial security, employment, education, contact with family, looked after status. |  |
| Research type | Quantitative, or mixed methods study designs which include a comparison. This can include comparisons with the same group before they received the intervention, or comparisons with a group that did not receive the intervention or received a different intervention.<br>We only included outcomes if they included a comparison time point or group, except in the case of measures of satisfaction with services.<br>Studies conducted in any country, language and with any publication date were eligible for inclusion. | Qualitative research<br>Narrative reviews, commentaries, editorials or book chapters which contained no primary research findings<br>Systematic or scoping reviews<br>Conference presentations or abstracts<br>Case studies<br>Theses/dissertations<br>Protocols<br>Study designs which described only outcomes for one group at one timepoint |

### Data extraction

A data extraction form was created in Microsoft Excel and piloted by three members of the research team (RA, CC, KT). Amendments were made to the form, and data was then extracted independently by two members of the research team (CC, PB, IM, UF, JY, JG, IE). All discrepancies were resolved through a third, senior member of the team.

### Quality appraisal

All papers were appraised using modified versions of the CASP checklists (Critical Appraisal Skills Programme, 2023). Details of modifications are available in Appendix 2. Quality assessment was conducted independently by two members of the research team (PB, RA, JY, JG, CC, IE), with randomised controlled trials (RCTs), cohort and other single group designs, and case-control studies assessed separately. Any disagreements were resolved with a third, senior researcher. Studies where multiple publications existed were appraised together where methods were the same (e.g. addition of extra follow up outcomes) and separately where alternative analysis and cohort selection methods may have been used (e.g. secondary analyses). Scores for the individual question items were combined to give a total overall score for each study. Whilst we did not exclude papers based on their quality, this was taken into consideration during interpretation of the findings.

While it is important to give detail on the certainty that can be placed on outcomes reported in systematic reviews e.g. through frameworks such as GRADE (Guyatt et al., 2011), there are a number of difficulties associated with assigning certainty of effects when including complex interventions in reviews (Montgomery et al., 2019). Furthermore, support models such as those of interest to this review utilise multiple treatment protocols flexibly which makes assessment of deviation from the intended intervention challenging, particularly in instances where all services used by individual participants are insufficiently reported. We therefore chose to consider certainty in reports of effects according to a) study design, and b) consistency in direction of effects in relation to null.

### Evidence synthesis

We conducted a narrative synthesis of data from included studies for each of our research questions. To describe early intervention models, we differentiated between models of support with different main treatment goals (for example, those which were trialling specific elements to increase speed of access versus those with a focus on more general provision of integrated support). We considered all available models when exploring effectiveness outcomes, although where important, we specified which type of model outcomes related to. We focused primarily on evidence based on comparisons to a contemporaneous control, prioritising more robust study designs such as RCTs to support interpretation of the best available evidence. Although not originally specified in our protocol, we also decided to summarise information regarding factors associated with intervention outcomes reported in the papers, as this data has potential interest for future policy making in this area.

We use the term “intervention” or “model” to describe specific intervention protocols, and the term “study” to describe evaluations conducted with a single sample. The term “publication” is used to describe separate individual publications describing specific outcomes of studies. For studies where multiple publications describe outcomes from the same sample, we refer to the primary (original) publication describing this intervention or model and the publication reporting specific outcomes.

## Results

### Study selection

The initial database search returned 9,572 studies of which 457 were deemed potentially eligible and assessed at full text. A further 149 studies were screened from citation and website searches. The update search returned 1067 studies of which 41 were assessed at full text. In total, 43 publications covering 38 studies met inclusion criteria and were included in the review. The full search and screening process is depicted in Figure 1, while reasons for excluding studies at full text are detailed in Appendix 3.

**Figure 1:**
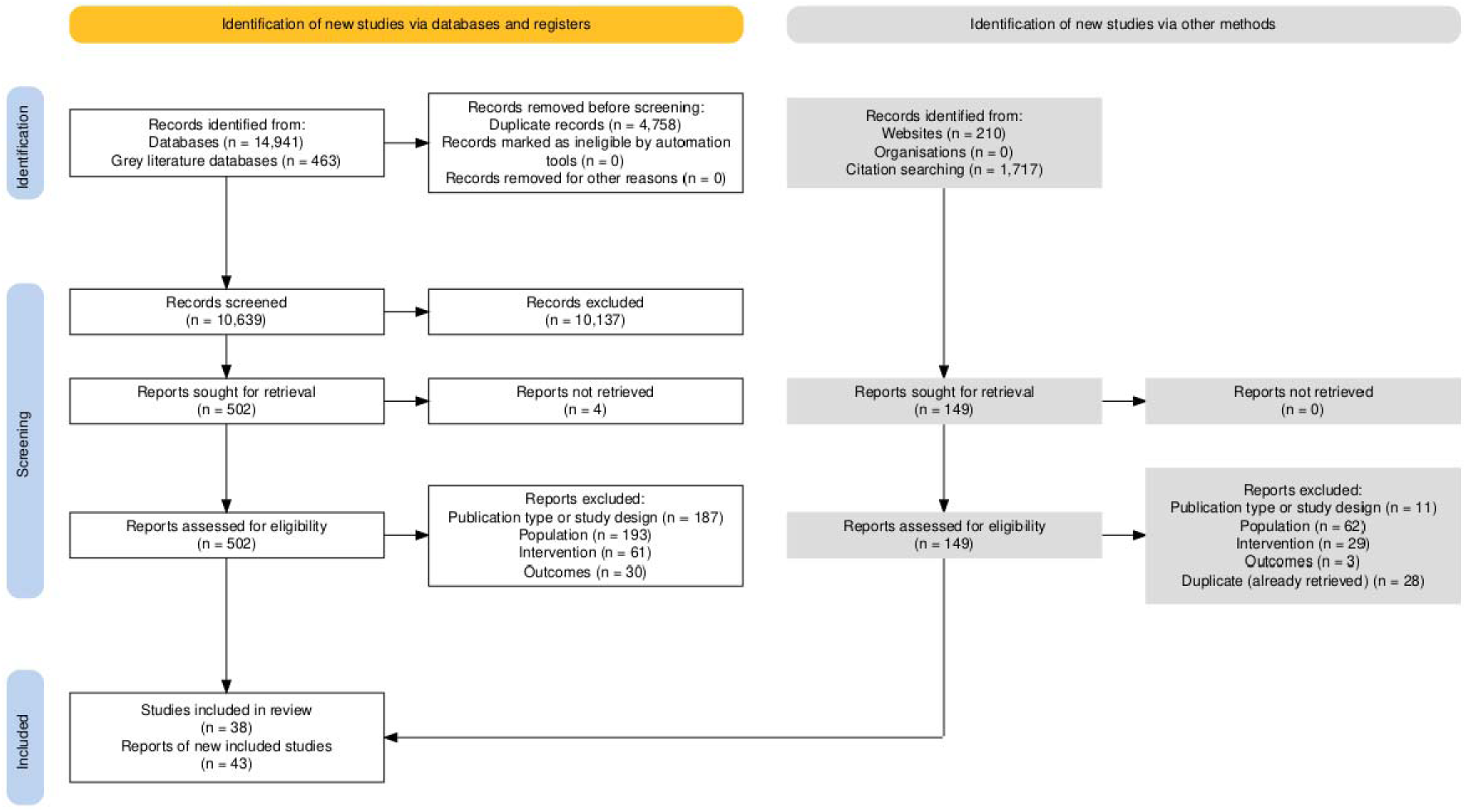
PRISMA flow diagram.

Studies were published in the USA (n=12), Australia (n=10), Canada (n=5), Ireland (n=4), the UK (n=2), the Netherlands (n=2), Chile (n=1), Japan (n=1), and Honduras (n=1). There were six RCTs (Asarnow et al., 2005; Kidd et al., 2020; Martínez et al., 2018; Richardson et al., 2014; Sheidow et al., 2021; Smith et al., 2012), as well as one cluster RCT(Sterling et al., 2018), and one study using a subset of data from this cluster RCT (Parthasarathy et al., 2021). We also included two trials which were non-randomised or quasi experimental (Saxe et al., 2012; Waters et al., 2022), four observational studies with a comparison group (Anderson et al., 2019; Osuch et al., 2019; Osuch et al., 2015; Telford et al., 2024), one study comparing cohorts seen before and after implementation of the model (Schley et al., 2019), and 22 studies reporting changes in outcomes in one group pre- and post-receipt of an intervention or service (Alvarez-Jimenez et al., 2020; Bond & Power, 2021; Boonstra et al., 2024; Chang et al., 2023; Corley et al., 2023; Dowell et al., 2021; Duncan et al., 2020; Hides et al., 2010; Hopkins et al., 2017; Kachor & Brothwell, 2020; Khazanov et al., 2024; Ng et al., 2023; O’Dell et al., 2020; O’Reilly et al., 2022; O’Keeffe et al., 2015; Parry et al., 2023; Peters et al., 2018; Rice et al., 2020; Schley et al., 2008; Souza et al., 2011; van Doorn et al., 2023; Waters et al., 2024). There was also one case-control study, but outcomes of interest were of a pre-post nature (Uchino et al., 2022).

The mean age of samples was between 13.12 and 21.75 years, and gender mix ranged from 100% females to 100% males. Ethnicity was often not reported, although where included, the majority of the samples were from white backgrounds. Regarding presenting problems, in those studies which reported symptoms, the majority reported only anxiety or depressive symptoms, although some also reported small proportions of their sample experiencing suicidal ideation or self-harm (n=5), social phobia, panic disorder or obsessive-compulsive disorder (n=4), personality disorder (n=2), or eating disorders (n=2). Two studies also reported that a small proportion of their sample (<10%) presented with symptoms of psychotic disorders. Appendix 4 provides further study characteristics and information on the individual results and publications reported within each study.

### Quality of included studies

Overall, the quality of the RCTs was high, with 5/11 scoring at least 75% of the maximum score and the remainder scoring at least 50%. Main reasons for lower scores centred around the quality of the reporting of results, although in most cases studies still partially met criteria. For example, often there was not a clear and specific description of the impact of the intervention on measured outcomes or clear articulation of the clinical implications of findings. RCTs with highest scores included Richardson et al. (2014) and Arsarnow et al., (2005; 2009), although secondary analyses of this latter trial were rated as lower quality due to differences in analysis methods (for example, subgroups which had more pronounced baseline differences between intervention and control, narrower focus on outcomes).

The quality of cohort studies was significantly more variable, with only two studies (Anderson et al., 2019; Souza et al., 2011) scoring at least 75% of the maximum score available. Eight of the 29 studies scored less than 50%. Main reasons for lower scores were a lack of consideration of confounding variables in both design and analysis (nine studies scored 0 for both of these aspects), insufficient follow up of participants, both in accounting for participants who had dropped out and in providing meaningful length of follow up in reporting of outcomes (only 9/29 studies reported outcomes for at least 80% of their baseline participants and only 6/29 reported at least 6 months follow up). Many also did not have a contemporaneous control comparison or reported outcomes which could not be trusted due to other methodological limitations such as small sample sizes (24/29). One case control study was included (Uchino et al., 2022). This study was of very low quality, scoring only 4/22. Details of individual quality ratings are also available in Appendix 2.

Study quality varied significantly, and we therefore chose to place more weight on evidence most likely to demonstrate valid results, including RCT designs and controlled cohort studies rated as high quality.

### Study and intervention characteristics

Interventions or models of care tended to fall under one of three broad categories: 1) Efforts to make care more comprehensive and joined up, 2) Specific efforts to increase the speed or ease of access to support, or 3) Provide targeted support for specific needs in addition to anxiety and depression. Models in category 1) were mainly whole service models which took the approach of integrating different forms of mental health and social support in primary care and providing a ‘joined up approach’ to supporting young people. Category 2) models explored effectiveness of an additional service component aimed at providing easier or earlier access to support, or of a strategy intended to result in faster overall response time (rather than aiming to improve the quality of the service delivered). Category 3) models aimed to provide targeted support for specific needs beyond anxiety depression, such as substance misuse, employment or homelessness. A brief description of interventions falling under each of these three categories is provided below.

### Models aimed at making care more comprehensive and joined up

We included 15 studies (described in 20 publications) testing 11 intervention models which had a general aim of making care more comprehensive and joined up (Anderson et al., 2019; Asarnow et al., 2005; Asarnow et al., 2009; Bond & Power, 2021; Chang et al., 2023; Corley et al., 2023; Khazanov et al., 2024; Martínez et al., 2018; Ngo et al., 2009; O’Reilly et al., 2022; O’Keeffe et al., 2015; Osuch et al., 2019; Osuch et al., 2016; Osuch et al., 2015; Parry et al., 2023; Peters et al., 2018; Rapp et al., 2017; Richardson et al., 2014; Uchino et al., 2022; Wells et al., 2012). These interventions attempted to encourage better integration of resources and treatment pathways in the community, and improve retention through involvement of multidisciplinary teams, flexible and personalised treatment options, and collaborative care provision. Although methods differed somewhat, all these interventions had a particularly all-round approach to improving all aspects of care from intake, retention and discharge, and usually aimed to take collaborative care or integrated primary care models as their starting point. For example, seven of the eleven models of care described specific efforts to individualise care options (Anderson et al., 2019; Asarnow et al., 2005; Bond & Power, 2021; Chang et al., 2023; Corley et al., 2023; Khazanov et al., 2024; O’Reilly et al., 2022; O’Keeffe et al., 2015; Osuch et al., 2019; Osuch et al., 2015; Richardson et al., 2014), with nine mentioning the offer of multiple treatment options and/or multidisciplinary, collaborative team provision (Asarnow et al., 2005; Bond & Power, 2021; Chang et al., 2023; Corley et al., 2023; Khazanov et al., 2024; Martínez et al., 2018; O’Reilly et al., 2022; O’Keeffe et al., 2015; Parry et al., 2023; Peters et al., 2018; Richardson et al., 2014; Uchino et al., 2022). Efforts to facilitate early detection and referral were described as central to the model in six of the eleven models, for example through integration of self-referral (Osuch et al., 2015), primary care screening (Martínez et al., 2018), drop-in service provision (Parry et al., 2023) or electronic referrals (Peters et al., 2018). In four models, specific pathways for onward specialist referral were integrated (Corley et al., 2023; O’Reilly et al., 2022; O’Keeffe et al., 2015; Parry et al., 2023; Richardson et al., 2014), while in five models, outreach or specific care manager follow up was employed to encourage ongoing engagement (Asarnow et al., 2005; Corley et al., 2023; Peters et al., 2018; Richardson et al., 2014; Uchino et al., 2022). Table 2 provides further details on each of these interventions.

**Table 2:**
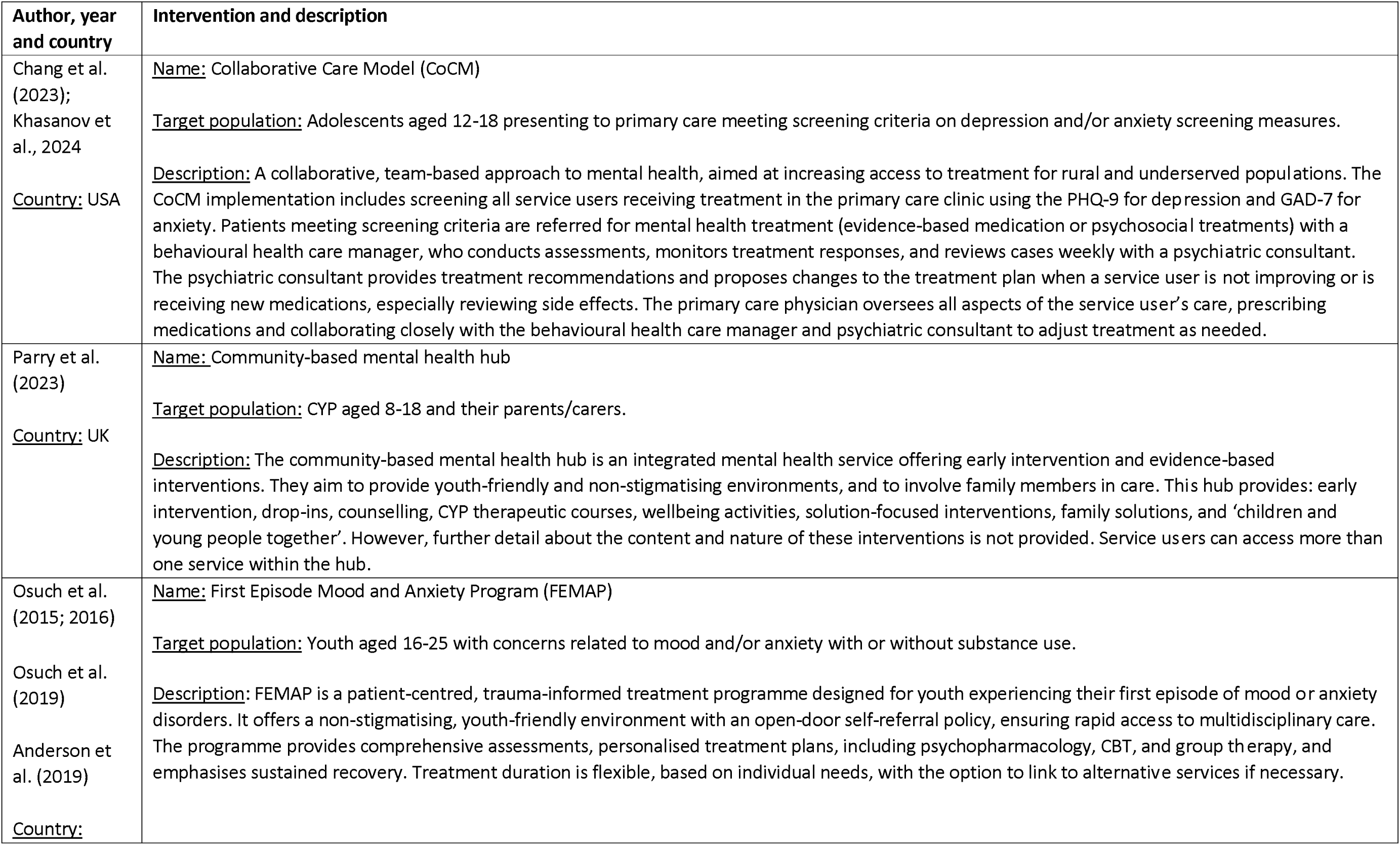
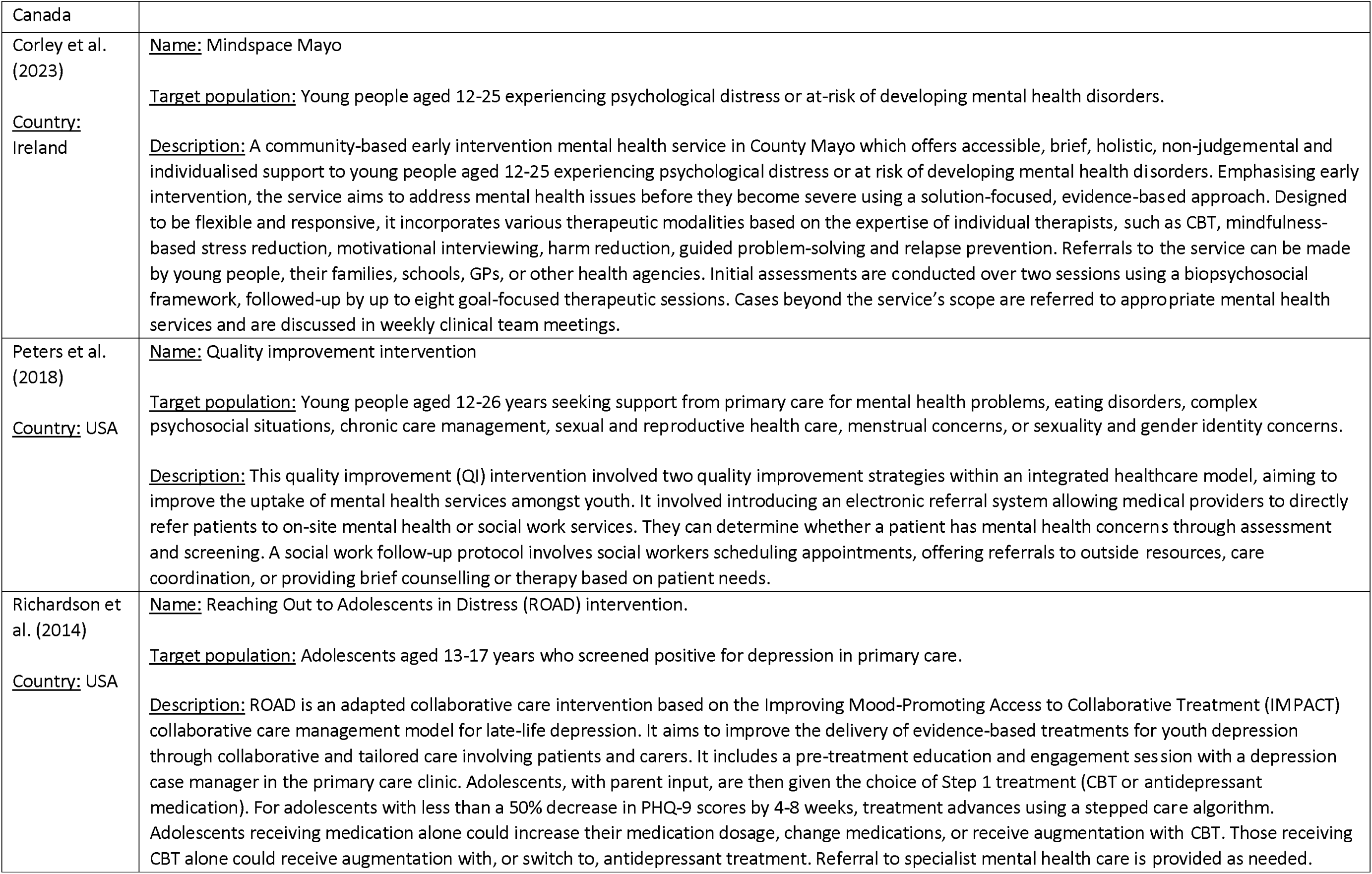
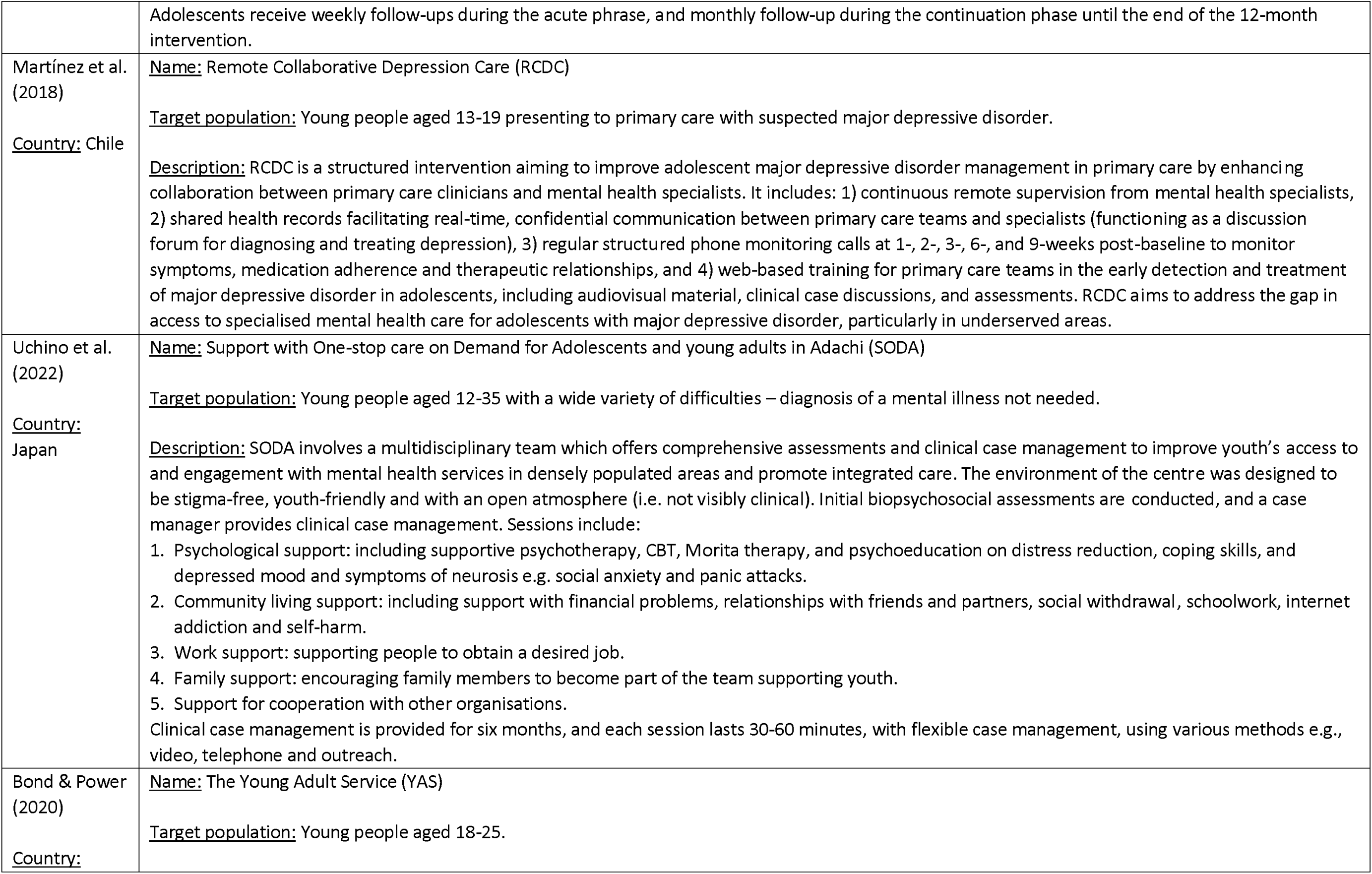
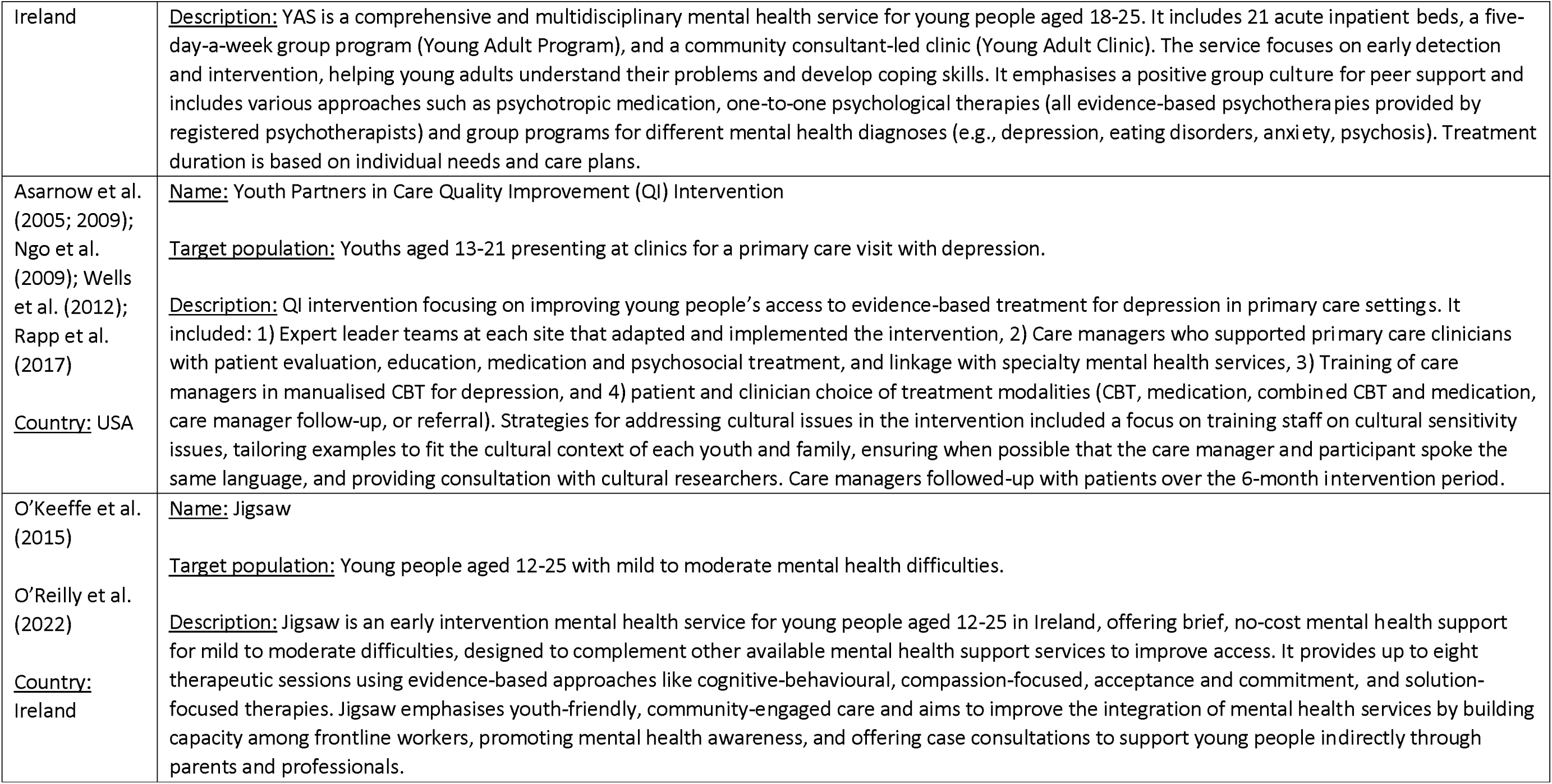
Outline of interventions with a main goal of making care more comprehensive and joined up.

### Models aimed at increasing speed or ease of access to support

We included 13 studies that, evaluated 9 intervention models which had a more specific target of increasing the speed and/or ease of accessing support for young people (Alvarez-Jimenez et al., 2020; Boonstra et al., 2024; Dowell et al., 2021; Duncan et al., 2020; Hopkins et al., 2017; Kachor & Brothwell, 2020; O’Dell et al., 2020; Rice et al., 2020; Schley et al., 2019; Schley et al., 2008; van Doorn et al., 2023; Waters et al., 2022, 2024). These models attempted to extend youth mental health services, which were usually also based on collaborative care models, and therefore aimed to establish the effectiveness of additional efforts to increase the speed or ease of access to available care through various means. For example, some aimed to improve access by integrating support into familiar and non-school based settings, such as primary care (O’Dell et al., 2020), community and voluntary sector services (Duncan et al., 2020), clients’ homes (Schley et al., 2008) or community sports programmes (Dowell et al., 2021; Waters et al., 2022, 2024), or by providing 24/7 access to support (Schley et al., 2008). Others utilised digital tools, such as social media-based platforms and digital resources, to provide young people with flexible and convenient support that they could access in a self-directed way (Alvarez-Jimenez et al., 2020; Dowell et al., 2021; Rice et al., 2020; van Doorn et al., 2023; Waters et al., 2022, 2024). Other models offered brief and targeted interventions to enable more timely access to support for those with lower complexity needs (Hopkins et al., 2017; Kachor & Brothwell, 2020; Schley et al., 2019). Some models targeted specific groups, including rural and underserved populations(Duncan et al., 2020) or males (Dowell et al., 2021; Rice et al., 2020; Waters et al., 2022, 2024). Table 3 provides further details on each of these interventions.

**Table 3.** Outline of interventions with a main goal of increasing speed or ease of access to care.

| Author, year and country | Intervention and description |
| --- | --- |
| Alvarez-Jimenez et al. (2020)<br><u>Country:</u> Australia | <u>Intervention:</u> Enhanced Moderated Online Social Therapy (MOST+)<br><u>Target population:</u> Young people aged 16 - 25 years who are help-seeking with concerns about their own mental health.<br><u>Description:</u> An accessible web-based youth mental health service embedded within a national web counselling service delivering immediate, short-term, flexible, and evidence-based support to help-seeking young people with mental ill-health. It offers 1) interactive user-directed web-based therapy, 2) peer-to-peer online social networking, 3) peer moderator support, 4) expert moderation, and 5) on-demand web chat with clinicians. All participants were provided with one week of access, with the option to extend enrolment on a weekly basis for a maximum of nine weeks. |
| Van Doorn et al., (2023)<br><u>Country:</u> Netherlands | <u>Name:</u> ENYOY-Platform<br><u>Description:</u> The ENYOY platform is based on the MOST platform (Alvarez-Jimenez 2020). It is a self-referral service provided online, involving biweekly coaching appointments and availability of the online platform 24/7 for 6 months. Therapy "journeys" are developed based on an algorithm and then can be tailored with a therapist according to preference. Peer workers also provide digital support. |
| O'Dell et al. (2020)<br><u>Country:</u> USA | <u>Name:</u> Acceptance and Commitment Therapy implemented within Integrated Primary Care (ACT-IPC)<br><u>Target population:</u> Youth aged 12-19 attending an integrated primary care clinic in rural USA.<br><u>Description:</u> ACT-IPC was designed specifically to address barriers to access to mental health care in ways that were feasible to implement in integrated primary care settings, for example through adaptations of content and home practices to reflect content that was relatable to participants, and incorporating techniques to encourage active participation, such as raffle entries for meaningful participation. Each session combined experiential and didactic exercises focused on ACT topics such as values, goals, mindfulness, defusion, creative hopelessness, self-compassion, willingness, self-as-context, and committed action. Participants were assigned weekly homework tasks. The intervention consists of nine 90-minute weekly sessions. |
| Rice et al. (2020)<br><u>Country:</u> Australia | <u>Name:</u> Entourage<br><u>Target population:</u> Young people aged 12–25 years inclusive with “probable” social phobia.<br><u>Description:</u> Entourage is a social media-based intervention which combines evidence-based therapeutic techniques for social anxiety with an engaging social media interface to ensure timely, ongoing access to support alongside usual care at local headspace early intervention support centres. Developed by a multidisciplinary team and young people with lived experience of social anxiety, it aims to improve social connection and address issues with past social anxiety interventions. It is used in a self-directed way over a 12-week intervention period. Alongside this, participants also receive continued |
|  | support at local headspace centres, addressing mental health, situational stressors, physical and sexual health, drug and alcohol counselling, and vocational support. |
| Schley et al.<br>(2019)<br><u>Country:</u><br>Australia | <u>Name:</u> Headspace Brief Intervention Clinic (BIC)<br><u>Target population:</u> Young people aged 12-25 presenting with a general mental health problem or mild to moderate levels of mental disorder, low level of risk to self/others and limited complexity (i.e. "simple" rather than multiple needs).<br><u>Description:</u> The BIC treatment model comprises eight skills-building and behavioural intervention models that young people can choose from, delivered over a maximum of six sessions by supervised graduate students. Developed to increase young people's access to mental health care, BIC addresses the challenge of timely access for those with lower complexity needs amidst increasing referrals to Headspace centres. Eligible young people, identified through the standard Headspace intake process, receive a biopsychosocial assessment and, if suitable, are enrolled in the BIC for weekly sessions. If BIC vacancies are unavailable, they follow the usual care pathway. The eight BIC modules cover common issues such as self-esteem, anger management, communication, exercise, problem-solving, mindfulness, mood and anxiety, and sleep. The BIC was refined with input from Youth Advisory Groups. |
| Dowell et al.<br>(2020)<br>Waters et al.<br>(2022)<br>Waters et al.<br>(2024)<br><u>Country:</u><br>Australia | <u>Name:</u> RISE Rugby League Development Programme<br><u>Target population:</u> Male rugby league players aged 12-15 years old.<br><u>Description:</u> The RISE program is a multi-component initiative that provides access to mental health support through integrating rugby league skills, strength and conditioning, with mental health and wellbeing support to address rising mental health issues in youth. It uses community-based participatory research as a conceptual framework, to involve all partners and leverage sports as an easy access point for early intervention. The program employs the integrated, three-step Life-Fit-Learning System, which includes:<br>1) Assess: assess the mental health and wellbeing of young people.<br>2) Reflect: provide immediate assessment summary reports at the individual level for young people, and organisational personnel at the aggregated cohort level.<br>3) Connect: connect young people to care via a multi-component intervention system including digital psychoeducational resources, group-based, modularised interventions, and tailored care for individuals at greatest risk.<br>Workshops are 30-40 minutes in duration and conducted in rotation with the tactical skills and strength and conditioning sessions at each RISE session. These workshops cover healthy lifestyle habits, grit and optimism, relaxation and mindfulness, and psychoeducation to explore activities focused on kindness and gratitude. |
| Hopkins et al.<br>(2016)<br><u>Country:</u> | <u>Name:</u> Single-session family therapy programme (SST)<br><u>Target population:</u> Young people presenting to a mental health service and their families or caregivers. |
| Australia | <u>Description:</u> SST, developed as an add-on at a Headspace centre, is a brief intervention aiming to meet the needs of young people in conjunction with their families and carers who may only attend a service once or twice, thereby reducing waiting times for the rest of the service. It aims to provide clinical care for young people, support for families and carers to understand their experiences, and enables families and carers to provide ongoing support at home to the young person. A pre-session questionnaire is mailed to families to help identify issues, and a session summary with solutions is provided afterward. Follow-up calls occur 4-5 weeks later to assess the need for further support. |
| Kachor et al.<br>(2020)<br><u>Country:</u><br>Canada | <u>Name:</u> Single-session therapy (SST)<br><u>Target population:</u> Youth aged 12-18 presenting with mild to moderate anxiety or depression, and their families.<br><u>Description:</u> SST is a future-focused, goal-directed and strengths-based approach to single-session therapy. It aims to reduce wait times and improves access to resources for those who may otherwise be required to wait for extended periods for multi-session therapy due to assessment of less severe presenting problems. On the SST appointment day, parents and youth complete a pre-session form to outline goals and assess distress levels. The youth and families were informed that the clinician has the intake information, meaning they would not have to repeat their story. The 90-minute SST session utilises solution-focused brief therapy and a client-centred, strengths-based approach. Families were also offered a telephone follow-up call one month later, to address any new or outstanding issues. |
| Schley et al.<br>(2008)<br><u>Country:</u><br>Australia | <u>Name:</u> The Intensive Mobile Youth Outreach Service (IMYOS) team (a sub programme of ORYGEN Youth Health (OYH))<br><u>Target population:</u> Young people aged 15-24 who display signs of mental illness, are considered at 'high-risk' (e.g. of suicide) and have a history of poor engagement with clinic-based services.<br><u>Description:</u> The IMYOS is a specialist mental health service for adolescents and young adults (ages 15-24), targeting youth experiencing signs of mental illness and considered at 'high-risk' (e.g. of suicide) and who have a history of poor engagement with clinic-based services. It employs a flexible outreach approach, offering support in natural settings, typically clients' homes, rather than requiring office visits. IMYOS operates during normal business hours, Monday to Friday. At other times, clients can contact the Youth Access Team, providing 24-hour, 7-days-per-week mobile assessment and acute community treatment. IMYOS is staffed by a multidisciplinary team, where each clinician has a caseload of 8-9 clients and provides around two home visits per week. Interventions include practical support and structured therapy, incorporating the individual's family and support system within a case management framework. |
| Duncan et al.<br>(2020)<br><u>Country:</u> UK | <u>Name:</u> Voluntary and community sector counselling services for CYP (Youth Information Advice and Counselling Services [YIACS]).<br><u>Target population:</u> Young people aged 11-25.<br><u>Description:</u> YIACS provide non-statutory counselling outside of the school environment. YIACS counsellors practice various therapeutic orientations, primarily person-centred and humanistic, with some offering cognitive or psychodynamic therapies. They aim to improve access to evidence-based practice and engagement for hard-to-reach and marginalised young people. The majority of the support offered is counselling, however, they also offer |
|  | information, emotional support, access to health clinics, and advice. Counsellors typically offer weekly sessions to clients. |
| Boonstra et al., (2024)<br><u>Country:</u><br>Netherlands | <u>Name:</u> @ease<br><u>Description:</u> @ease provides a safe and easily accessible space for young people aged 12–25 years to talk about their mental health problems or related concerns with trained peer counsellors in a youth-friendly environment. A healthcare professional is available on site and a psychiatrist is available on call to advise peer counsellors. Sessions involve active listening, solution-focused strategies, motivational interviewing, suicide prevention, noticing when other forms of care might be required and informing the young person how to access other services. |

### Models aiming to provide targeted support for specific needs in addition to anxiety or depression

We included ten studies which had the aim of enhancing existing healthcare through providing targeted support for specific needs in addition to anxiety or depression (Hides et al., 2010; Kidd et al., 2020; Ng et al., 2023; Parthasarathy et al., 2021; Saxe et al., 2012; Sheidow et al., 2021; Smith et al., 2012; Souza et al., 2011; Sterling et al., 2018; Telford et al., 2024). Some studies did this by combining mental health care with social support for other problems faced by their target population e.g. housing or education and employment (Kidd et al., 2020; Souza et al., 2011; Telford et al., 2024). Some interventions also aimed to support young people with substance misuse (Hides et al., 2010; Sheidow et al., 2021; Sterling et al., 2018) or other comorbidities (Parthasarathy et al., 2021). One example of this approach involved specifically screening for substance misuse problems and providing brief interventions in a primary care setting, with a protocol for referring onto specialist support if needed. Three studies aimed to support young people with trauma or post-traumatic stress disorder (PTSD) (Ng et al., 2023; Saxe et al., 2012; Smith et al., 2012). Three studies mentioned components of the intervention aimed at involving parents or families in the young person’s mental health support (Saxe et al., 2012; Sheidow et al., 2021; Smith et al., 2012). Table 4 provides further details on each of these interventions.

**Table 4:** Outline of interventions with a main goal of providing targeted support for specific needs in addition to anxiety or depression.

| Author, year and country | Intervention and description |
| --- | --- |
| Hides et al. (2010)<br><u>Country:</u><br>Australia | <u>Name:</u> Self-Help for Alcohol/other Drug use and Depression for Young People (SHADEY)<br><u>Target population:</u> Young people aged 15-25 with concurrent major depression and substance misuse.<br><u>Description:</u> SHADEY is an individual, integrated CBT intervention incorporating motivational interviewing, cognitive behavioural, and mindfulness skills within a harm minimisation framework. It provides multifaceted support for comorbidities, and holistic care to target social problems. Participants receive 10 sessions of individual targeted CBT treatment delivered with CBT over a 20-week period. |
| Kidd et al. (2020)<br><u>Country:</u><br>Canada | <u>Name:</u> Housing Outreach Program-Collaboration (HOP-C)<br><u>Target population:</u> Young people aged 16-26, residing within city limits, who had experienced at least 6 months (not necessarily consecutive) of homelessness, and had been housed in a stable arrangement (i.e., not a crisis shelter, not couch surfing) between 1 day and 1 year since their last homeless episode.<br><u>Description:</u> HOP-C is a multicomponent, multidisciplinary team-based critical time intervention (lasting 6 months) providing mental health and peer support, along with transitional case management, for youth who have transitioned from homelessness to stable housing. The multifaceted support includes transitional case management to support with navigating systems like housing, education, employment, justice and health alongside peer support and group mental health support interventions. The peer workers have their own personal experience of homelessness and marginality, and successful transitioning into housing, employment and education. This support was delivered through collaboration between multiservice organisations serving youth who have experienced homelessness (providing the case managers), an arts organisation (providing access to “feeling” spaces for programming and peer support), and a tertiary psychiatric hospital (providing the psychologist and connection to crisis services or consultation). |
| Ng et al. (2023)<br><u>Country:</u> USA | <u>Name:</u> The Primary Care Intervention for Post-traumatic stress disorder (PCIP)<br><u>Target population:</u> Young people aged 12-26 presenting to primary care suspected of having PTSD symptoms.<br><u>Description:</u> PCIP is a three-session, low-intensity, evidence-based PTSD intervention for young people designed to increase access to timely treatment and address treatment disparities for low-income and youth from ethnic minorities. The three sessions cover: 1) psychoeducation to enhance knowledge of PTSD symptoms, self-recognition, and motivation for treatment, reducing trauma-related cognitions, 2) breathing retraining to increase vagal tone, normalise sympathetic nervous system overactivation, and reduce physiological arousal in response to trauma reminders, and 3) positive coping strategies to facilitate effective management of stress and comorbid anxiety and depression symptoms. Homework tasks reinforce skills in real-world settings, promoting generalisation, proficiency and confidence. Each session lasts 30-50 minutes, delivered over 3 weeks to 3 months. |
| Sterling et al. (2018)<br>Parthasarathy et al. (2021)<br><u>Country:</u> USA | <u>Name:</u> Screening, brief intervention, and referral to treatment (SBIRT)<br><u>Target population:</u> Adolescents (aged 12-18) presenting to primary care with comorbid mental health and substance use difficulties.<br><u>Description:</u> SBIRT in primary care services aims to prevent and provide early intervention for substance use problems in youth. It involves systematic screening using evidence-based treatments, patient-centred brief interventions (typically informed by motivational interviewing), and protocols for referring severe patients to behavioural health treatment. There is no set treatment duration. |
| Saxe et al. (2012)<br><u>Country:</u> USA | <u>Name:</u> Trauma Systems Therapy (TST)<br><u>Target population:</u> Young people with trauma histories and prominent symptoms of PTSD, and their families.<br><u>Description:</u> The TST approach was designed to address the socio-ecological needs of children with trauma histories through a coordinated and integrated treatment plan. It aims to improve engagement and retention of children and families in treatment, particularly in marginalised populations, by embedding strategies for outreach and engagement at both the organisational and individual level. Delivered by a multidisciplinary team, the TST program offers four service modules: skills-based psychotherapy, psychopharmacology, home- and community-based care, and systems advocacy. Treatment begins with the “Ready-Set-Go!” module, which focuses on forming a therapeutic alliance, addressing barriers, and providing psychoeducation on traumatic stress. Families are offered various configurations of these service models based on their needs. |
| Sheidow et al. (2021)<br><u>Country:</u> USA | <u>Name:</u> Integrated family-based outpatient treatment for adolescents (OPT-A).<br><u>Target population:</u> Young people aged 10-17 requiring treatment for current comorbid substance use and internalising (mood or anxiety) disorders.<br><u>Description:</u> OPT-A is a novel clinic-based treatment approach that draws on multisystemic therapy. It provides comprehensive conceptualisation and treatment of multiple problems simultaneously rather than sequentially. It integrates multiple interventions simultaneously, depending on service users’ needs, and guides therapists to integrate specific strategies from evidence-based interventions. These include CBT techniques, behaviour medication targeting substance use and externalising behaviours, parents and communication training. Sessions occur weekly and treatment length is criterion-based, averaging 9.7 months. This approach ensures comprehensive and flexible support for families and adolescents. |
| Smith et al. (2012)<br><u>Country:</u> USA | <u>Name:</u> Multidimensional Treatment Foster Care adapted for girls with co-occurring trauma and delinquency (MTFC+T)<br><u>Target population:</u> Females, ages 12–17, at least one arrest in the year prior to referral, court mandated for out-of-home placement, at least one traumatic experience, and not currently pregnant.<br><u>Description:</u> This is a community-based fostering intervention for the treatment of ‘chronically delinquent adolescents and their families’, adapted to include more trauma-focused care. It aims to simultaneously treat trauma and delinquency. The model includes daily phone contact with parents, weekly parental group supervision, individual therapy, skills building practice, family therapy, case management and psychiatric consultation where |
|  | needed. The MTFC+T intervention had two phases for youth: Phase I focused on emotion-regulation, coping, and problem-solving skills, while Phase II focused on trauma-focused education, skill-building and processing. |
| Souza et al. (2011)<br><u>Country:</u><br>Honduras | <u>Name:</u> Multidisciplinary care provided in a drop-in centre<br><u>Target population:</u> Homeless youth under 25 years old<br><u>Description:</u> The intervention adapts the community reinforcement approach to address the clinical needs of homeless individuals, including substance use, homelessness and mental health problems, using an operant perspective. It assumes that environmental contingencies play a powerful role in behaviour change. It aims to improve access to medical or psychological support, laundry, showers, recreation and a place to rest for street youth. It involves four components:<br>1. Mental health strategy: utilising the community reinforcement approach for substance use, homelessness and mental health issues.<br>2. Medical strategy: Provides health education sessions on the most common diseases amongst street children.<br>3. Social care strategy: Developed a complete mapping of services available for street children, depending on their interests and needs.<br>4. Education strategy: Provides coordinated street outreach activities, recreational activities, and tracking children who did not attend the centre as planned. |
| Telford et al. (2024)<br><u>Country:</u><br>Australia | <u>Name:</u> Individual Placement and Support (IPS) added to HEADSPACE<br><u>Description:</u> Headspace centres are a network of community based, accessible and integrated support for youth. Fifty headspace centres offer the standardized IPS model as a vocational stream within their service offering. This includes time unlimited vocational services, integration of vocational and mental health support, and zero exclusion. The IPS work and study specialists work with young people to explore their individual preferences, interests and strengths and find suitable and competitive employment and/or education opportunities. The support includes resume writing and interview preparation. They also provide follow-up support once employment or education has been obtained. In addition to working directly with young people, the IPS work and study specialists visit local employers to build relationships in the community. |

### Effects of interventions

Despite differences in the overall goal of models of support, the three categories of model had similar patterns of outcomes. Results are therefore summarised together to give an overview of the impact of early intervention approaches to supporting young people overall. Further information on results of individual studies is available in Appendix 4.

### Outcomes relating to access and waiting times for mental health support

Comparisons with contemporaneous control groups were reported for outcomes relating to access and waiting times for mental health support for two RCTs (Asarnow et al., 2005; Richardson et al., 2014) and one high-quality observational study (Anderson et al., 2019). All these studies examined models aiming to make care more comprehensive and joined up. Results suggest that there were significantly greater odds of receiving mental health appointments (e.g. psychotherapy) within the first six months of the QI intervention being implemented for the innovative model than for usual care (Asarnow et al., 2005; Wells et al., 2012). In one RCT, participants in the intervention group also had more rapid access to a psychiatrist and lower rates of emergency department use for mental health reasons in the first six months (Anderson et al., 2019). In another, a higher proportion of participants in the intervention group received high quality care (defined by study authors as receipt of sufficient counselling and medication; 86% vs 27% TAU; Richardson et al. (2014)). However, at longer follow-up points (12-18 months) receipt of antidepressants and use of speciality mental healthcare were similar to control groups.

Changes in service access between different cohorts seen before and after implementation of the intervention were reported in three studies of models that aimed to make care more comprehensive and joined up (Osuch et al., 2015; Parry et al., 2023; Uchino et al., 2022), and two of models that focused specifically on increasing speed or ease of access (Kachor & Brothwell, 2020; Schley et al., 2019). Studies of models that focused on increasing speed or ease of access to services (both using brief interventions) reported improvements in waiting times after implementation of the model (Kachor & Brothwell, 2020; Schley et al., 2019), although no significance testing was conducted and the study by Kachor and Brothwell (2020) evaluated a pilot. An increase in demand for services was reported for two of the studies of models focused on making care more comprehensive and joined up (Osuch et al., 2015; Uchino et al., 2022), and this increased demand for resources was reflected in mixed results regarding improvements in wait times (Osuch et al., 2015; Parry et al., 2023; Uchino et al., 2022).

### Outcomes relating to mental health and wellbeing

Comparisons for mental health and wellbeing outcomes between an intervention and a contemporaneous control group were reported in 11 studies, of which four investigated models aiming to make care more comprehensive and joined up (RCTs: Asarnow et al. (2005); Martínez et al. (2018); Richardson et al. (2014); observational study: Osuch et al. (2019), one focused on a model which aimed to improve ease of access through mental health provision within a sports league (Waters et al., 2022), and six reported RCTs of models aiming to provide targeted support for specific needs in addition to anxiety or depression (RCT: Kidd et al. (2020); Parthasarathy et al. (2021); Sheidow et al. (2021); Sterling et al. (2018), Pilot RCT: Smith et al. (2012), observational with matched control: Telford et al. (2024)). Results suggest that interventions aimed at making care more comprehensive and joined up were effective in improving mental health outcomes, with both the Reaching Out to Adolescents in Distress (Richardson et al., 2014) and the Youth Partners in Care QI intervention (Asarnow et al., 2005; Asarnow et al., 2009) RCTs demonstrating significantly larger improvements in depression severity at 6 months favouring innovative models over controls. However, at 12 months, differences only remained significant in the RCT conducted by Richardson et al. (2014).. Another RCT investigated a remote format of telephone-based collaborative care (Martínez et al., 2018), but did not report any significant benefits of the intervention compared to usual care at 12 week follow up. There were no significant differences from usual care in self-harm presentations in the QI intervention (Asarnow et al., 2005). Wellbeing outcomes, reported in the RCTs conducted by Asarnow et al. (2005) and Martínez et al. (2018), were only reported to be significantly improved compared to controls in the QI intervention (Asarnow et al., 2005), however this was only present at 6-months follow up, and differences disappeared at 18 months.

The non-randomised trial that aimed to improve access to mental health support through a rugby league development programme (RISE) reported that anxiety, depression and behavioural symptoms reduced significantly more than in the waitlist control group, although only amongst those identified as ‘high risk’ of mental health problems (Waters et al., 2022). All but one intervention that aimed to provide targeted support for specific needs in addition to anxiety or depression (Kidd et al., 2020) reported a significant effect of the intervention on mental health or wellbeing outcomes compared to the control, including for emergency department use for mental health reasons at longer (3 year) follow-ups (Parthasarathy et al., 2021), depression (Parthasarathy et al., 2021; Sterling et al., 2018), substance misuse (Parthasarathy et al., 2021; Sheidow et al., 2021), quality of life (Telford et al., 2024), and trauma-related mental health symptoms (Smith et al., 2012). Both studies which reported substance misuse outcomes identified significant differences between the intervention and control groups at their longest follow-up time points (3 years and 18-months respectively), despite non-significant differences at earlier follow-ups (Parthasarathy et al., 2021; Sheidow et al., 2021). Comparisons of individuals before and after receipt of an intervention also tended to demonstrate improvements across a range of mental health and wellbeing outcomes over time (see Appendix 4). No studies identified any adverse effects of the interventions.

### Outcomes related to social functioning or social needs

There were three RCTs and one matched cohort study with a specific aim of providing support to improve social outcomes. One RCT which aimed to support youth who had recently transitioned out of homelessness with their mental health (Kidd et al., 2020) found that there was no difference in the likelihood of gaining or sustaining either housing or education/employment between participants who received the intervention and participants in the usual care control group at six months follow-up (Kidd et al., 2020). There was also no statistical difference in education outcomes (e.g. school enrolment) reported in an RCT comparing an integrated family-based outpatient treatment programme designed to support multiple problems simultaneously with control (Sheidow et al., 2021). However, one pilot RCT (Smith et al., 2012) of a community fostering intervention for girls with involvement in crime reported that the girls who took part in the intervention had significantly lower levels of law-breaking compared to treatment as usual at 12-months post-baseline. Telford et al. (2024) reported that addition of employment support to early intervention (HEADSPACE services) supported 65% of participants to obtain a job or commence study, compared to 12% who did not receive integrated employment support.

Of the studies of models with a more general aim of making care more comprehensive and joined up, one (Osuch et al., 2019) conducted post-hoc comparisons of improvements over time compared with a waiting list group and reported that functioning improved more in a group receiving a QI intervention (Youth Partners in Care) than in the waiting list group.

A range of other improvements were also reported for social outcomes, including loneliness and social support, goal-based outcome scores and violence (see Appendix 4) in comparisons between baseline and follow-up within cohorts without a comparison group.

### Outcomes relating to cost-effectiveness

One process evaluation study reported information relating to cost effectiveness (Osuch et al., 2015). This study reported that four months of treatment using a patient-centred, trauma-informed multidisciplinary model of care (FEMAP) would cost $1634 (CAD), however the study estimated that without this early intervention, there would be a cost of $2188 for psychiatric evaluation under the mental health act in Ontario and an additional $4392 for disability support for four months, in youth experiencing their first episode of anxiety or depression. Estimates, therefore, suggest that early outpatient treatment via FEMAP was more cost effective.

### Outcomes relating to acceptability

Acceptability outcomes generally suggested that participants across all early support interventions, where this was compared with a control (n=6), were more satisfied with care in intervention groups. In two RCTs (Asarnow et al., 2009; Richardson et al., 2014) and an observational study with waitlist comparisons (Osuch et al., 2019), satisfaction was significantly higher compared to controls. There were also significantly higher levels of satisfaction compared to usual care in an RCT of integrated family-based outpatient treatment specifically aiming to improve multiple presenting problems (Sheidow et al., 2021). Furthermore, although satisfaction with care did not improve significantly over time in those participating in the RISE rugby league intervention, it decreased in the control group (Waters et al., 2022). However, differences in satisfaction compared to control were only seen in the short term (6 months) during the Quality Improvement Intervention (Asarnow et al., 2009), and only in relation to psychological care (rather than facilities, medical care and non-professional staff treatment) in a collaborative care intervention delivered remotely (Martínez et al., 2018). Other descriptive reports of acceptability without a control (n=8) also tended to suggest that participants found interventions acceptable (see Appendix 4).

Results relating to drop-out were mixed. Although access to support was greater in the intervention group, Richardson et al. (2014) also reported that more participants dropped out of the collaborative care intervention than the usual care group. There was also no significant difference in continuation of case management contacts in previously homeless youth who received a Critical Time Intervention compared to control participants (Kidd et al., 2020). However, in a quasi-experimental study of trauma systems therapy, at the 3-month reassessment point, 90% of patients receiving the intervention were still enrolled in treatment compared with only 10% of patients in the treatment as usual condition, although the sample size for this study was only twenty (Saxe et al., 2012).

### Factors associated with outcomes

Two studies suggested that some age-related and ethnic inequalities in access to care and mental health outcomes (Ngo et al., 2009; Rapp et al., 2017) could be reduced with efforts to improve care-coordination and access. One of these also reported that not having English as a primary language was associated with poor access (Rapp et al., 2017). Furthermore, although two studies reported that baseline symptoms and diagnosis were not clearly associated with mental health outcomes (Parry et al., 2023; Smith et al., 2012), and one reported that higher baseline severity predicted greater improvements in distress and psychosocial functioning (Van Doorn 2023), it was reported in one study that older adolescents (aged 17+) with significant complexities (presenting with multiple problems) may have less positive outcomes than younger adolescents (Corley et al., 2023), making the impact of severity unclear across all early interventions. Models which provided personalised intervention options based on diagnosis or preferred format were found to be associated with improvements in mental health and satisfaction in two studies (Bond & Power, 2021; Waters et al., 2022). Although time spent in receipt of support was predictive of improvements in one study (Khazanov et al., 2024), in general, characteristics of treatment such as the number of sessions or specific interventions given were not reported to be associated with outcomes (Corley et al., 2023; Parry et al., 2023; Schley et al., 2019). Although, one study reported that the small proportion of their sample who had been referred from previous youth mental health services were more likely to drop out (Bond & Power, 2021), and another found that youth referred by schools were significantly more likely to experience deterioration in goal-based outcomes compared to those referred by other sources.

Further information on reported factors associated with outcomes is available by variable and outcome in Appendices 5a and 5b, respectively.

## Discussion

This systematic review aimed to summarise the evidence base for the effectiveness of early intervention models aimed at the first onset of anxiety and depression or other common mental health problems in young people. We found that generally, evidence thus far has focused on i) the impact of efforts to make care for young people experiencing symptoms of depression or anxiety more comprehensive and joined up, ii) the impact of interventions to make access to the support needed faster or easier, and iii) targeted efforts to evaluate the impact of more specific interventions to support young people with specific comorbid or additional needs.

Although a large body of the current evidence base consists of single group pre-post comparisons, there was some controlled evidence that early intervention approaches aiming to better serve young people experiencing early signs of anxiety or depression may improve outcomes. These include both access to mental health treatment and subsequent mental health outcomes in the short-term, including for minority ethnic groups. There was less evidence that these improvements were maintained at longer term follow-ups, mirroring findings of early intervention in psychosis studies (Bertelsen et al., 2008; Gafoor et al., 2010). This suggests that sustained efforts to improve quality of care may be needed to provide ongoing support to improve the longevity of service improvements.

There was also some evidence that targeted efforts to adapt such support to further improve speed of access can reduce wait times for treatment, though evidence on this was limited, and some studies did not test the statistical significance of these changes. As faster access to treatment has been found to increase rates of recovery and improvement in adults (Clark et al., 2018), targeted efforts such as these should be the focus of future research in this area, although it is also noteworthy that some models without this explicit aim also reported improved access. Intervening early to support youth with their mental health is important, as this can reduce the risk of presentations becoming more severe and requiring more intensive support such as inpatient CAMHS, or facilitate transitions to these services where needed (McCrone et al., 2013). However, in line with findings from a review of health access in adults (Twersky et al., 2024), improvements to access were found only for English-speaking participants in one study which explored this (Rapp et al., 2017), suggesting that further outreach work with, and involvement in research of groups typically underrepresented in mental health care may be required.

While evidence from one study indicated that an early intervention model providing support for people with general mental health symptoms did not improve substance use (Schley et al., 2008), there was some evidence that interventions providing concurrent support for both mental health and substance use, for example through provision of separate screening and brief intervention, led to significant and long-term improvements in substance use outcomes. As previous research has found that young people reporting early mental health symptoms report more substance use at ages 17-19 (Henderson et al., 2021), ensuring that early intervention efforts also support, as a minimum, screening for and signposting for substance use, may be an important aspect of effective early support for young people. Although there was some evidence to suggest that early intervention models can facilitate an improvement in measures of functioning, social and functioning outcomes were overall not well reported. Limited RCT evidence reported no difference compared to usual care in improvement in housing, or education and employment through interventions with specific population targets of either previously homeless youth or youth presenting with multiple difficulties. However, there was evidence from another RCT that community fostering efforts may be more effective than usual care in supporting reductions in offending.

Almost all the available evidence suggested that early intervention models of support are acceptable to youth and more so compared to usual care – an important finding given emphasis on patient and family experience of care (NHS England, 2016). Youth have reported negative experiences of mental health care and transitions in the past (Appleton et al., 2021), highlighting the importance of provision of acceptable care that could limit negative experiences that may result from inpatient treatment or emergency department presentations (Anderson et al., 2019; Clark & MacLennan, 2023). This finding supports the argument that providing early support may contribute to reducing stigma and negative experiences surrounding mental health intervention.

### Strengths and limitations

We utilised a comprehensive search strategy covering multiple academic and grey literature databases alongside rigorous review methods, including independent dual screening and extraction. We also integrated lived experience and topic expert involvement throughout these methods and considered a diverse range of study designs and conceptualisations of early intervention approaches to ensure the breadth of the literature was covered. Similarly, we considered a broad range of outcomes, meaning this review captures evidence on the extensive impact of these models.

However, the following limitations should be considered alongside interpretation of results. As is the case for all reviews of the literature, the strength of conclusions is constrained by the robustness of included literature, which in this instance was predominantly studies with only pre and post intervention measures. Our conclusions are also limited by a lack of evidence for some of our review questions, for example regarding cost effectiveness and social outcomes, variability in measurement (outcome measures used and length of follow up) and target populations. Included studies also often had limited detail on how (often highly complex) interventions were implemented, fidelity to established intervention protocols, and the extent of participants’ contact with individual components. Relatedly, our focus was on early intervention provided within a complex care model to establish what may be additionally beneficial compared to current service provision, however, this meant that some potentially effective stand-alone interventions for preventing exacerbation of early symptoms of depression or anxiety were not considered (e.g. self-help apps). There may be benefit in comparing outcomes for standalone interventions to those involving complex care. We also acknowledge that the effectiveness of early intervention services will ultimately depend on what types of support they offer, not just how well coordinated they are and how well the facilitate access, and we were limited in how far we were able to describe the extent of provision of specific evidence-based interventions within these complex early intervention services. We categorised interventions according to the main goal of the research describing them to inform understanding of the different approaches and efforts to improve early intervention. However, there is likely to be significant overlap between these categories and we did not use an established typology or test the reliability and validity of this categorisation. We also did not include qualitative accounts in this review, which could have provided further insight into patient, carer and staff experiences of early support, and what works well for whom, in what circumstances, and why.

### Implications for research and practice

This review indicates preliminary effectiveness of several models of early intervention in youth mental health, warranting further research and implementation. We are unable to draw firm conclusions on which models are most effective based on the evidence in this review alone, therefore any future implementation of these models should also feature embedded evaluation of outcomes. Over 60% of the studies in this review consisted of evaluations with only single group, pre-post reports of effects. As measures of mental health and wellbeing in particular are subject to both the effects of time (Vergunst et al., 2013) and simply having the opportunity to voice one’s problems (Oei & Shuttlewood, 1996), comparisons to controls in robust research designs such as RCTs with long-term follow-ups are necessary to establish whether particular approaches can improve outcomes for young people, and the longevity of these effects. It is also not clear whether young people may have a preference regarding which interventions should be offered. For example, while one study reported that attendance at interventions targeting specific symptoms identified could predict better outcomes (Bond & Power, 2021), this was not widely explored.

Many of the models of support described in this review aimed to make care more comprehensive and joined up, with some studies exploring additional ways to improve outcomes such as through methods to improve ease of access or provide targeted support for young people with specific concurrent difficulties. Only some integrated evidence-informed mental health support and intervention alongside additional holistic services to suit need, in line with research establishing common principles of “integrated community based youth service hubs” (Settipani et al., 2019). This heterogeneity in precise protocols of care and the intensity of mental health intervention provided reflects a key component of early support models for young people, as support can be based on need. For example, we found some evidence that the number of treatment sessions and intervention type received within services was not associated with effectiveness outcomes, although in one study provision of treatment individualised to presenting problems was found to predict positive outcomes (Bond & Power, 2021). This suggests that flexible, individualised support options are an important element of provision of early intervention and adapting support to suit need may be one way to facilitate ongoing engagement and satisfaction while maintaining positive impacts on mental health. However, this key strength presents a limitation for establishing effectiveness of care, as the variation in support provided prevents roll-out of replicable models with demonstrable effectiveness. Therefore, detailed information regarding the support received by individuals should be a key element of routine outcomes monitoring in services to allow exploration of the effectiveness of support options offered on a case-by-case basis, alongside co-produced approaches to understanding critical ingredients to effective care. It should be noted that the models reported on in our review tend to include professional staff and evidence-based clinical interventions (psychological and sometimes pharmacological). We cannot, therefore, draw conclusions as to whether youth support services in which specific mental health interventions may not be offered are effective (such as the young people’s hubs increasingly found in England (CYPMHC, 2024).

Finally, scale-up of early intervention for young people has the potential to save money through mitigating the progression of mental health problems, however there is at present almost no evidence of cost effectiveness of these models. Cost-effectiveness analyses are needed to assess the economic and social value of early interventions at the individual and societal level. By quantifying these benefits relative to the potentially large upfront cost of scaling up support offerings for large portions of the population, this research is very important in informing future funding decisions which are evidence-based and maximise use of public resources.

## Conclusions

There is evidence to suggest that models of early intervention for young people experiencing symptoms of comment mental health problems can reduce wait times and result in improved mental health and wellbeing, whilst being acceptable to young people. However, there are some limitations of the quality of existing evidence, in part due to the high heterogeneity of interventions and variability in outcomes measured. Around a quarter of studies had small sample sizes, and there was a lack of studies with a contemporaneous control group. Therefore, it was not possible to draw firm conclusions regarding impacts on certain outcomes of interest, including cost-effectiveness of these models and their impact on social outcomes such as housing or employment. Future research in this area should focus on conducting more robust evaluations, including collecting more comprehensive information about the exact support received by young people as part of the intervention, to inform what an optimum model of early intervention support for young people should look like.

## Lived Experience Commentary

### Written by two members of our working group with lived experience: Nima and SB

Our insights reflect our lived experience of mental health challenges, including the use of CAMHS and alternative (non-NHS) services that support young people.

This review highlights the benefits of collaborative care models, which effectively reduce wait times and incorporate family and community resources. However, it lacks emphasis on how mental health professionals can better understand the unique sociocultural and psychological complexities faced by children and young people (CYP) and the resulting impact on their mental health. For instance, discussions about academic pressures and cultural expectations experienced by CYP often skimmed the surface, neglecting deeper issues such as anxiety about future prospects and communal pressure that creates internal conflict and feelings of inadequacy. Enhanced training involving workshops and role-playing scenarios for those who work in early intervention services would ensure effective engagement with this demographic.

While the review rightly suggests further outreach to underrepresented groups, it should also dissect how cultural and demographic factors influence access to care and its outcomes. For example, younger individuals may fear being dismissed or misunderstood by older professionals, deterring them from seeking help. Similarly, those with disabilities and/or cooccurring health needs alongside their mental ill-health may feel perceived as ‘too complex’ for early intervention services. Additionally, in cultures where mental health challenges are commonly perceived as a weakness, there is a fear of judgement that can further complicate one’s willingness to seek support. This hesitation also hinders individual recovery by perpetuating a cycle of silence and misunderstanding surrounding mental health issues. Collaborative efforts are crucial; co-production with underrepresented groups would ensure these models better serve diverse communities and improve mental health outcomes.

A diverse range of innovations in early intervention for CYP were included in this review, creating a backdrop from which the breadth of approaches employed by early intervention services can be understood. Whilst some services proclaim to offer ‘non-stigmatising’ or trauma-informed treatments, none appeared to implement approaches which divert from a medical model way of understanding and responding to distress. As the call for non-pathologising services grows, particularly in lived experience and survivor circles, research that measures the effectiveness of non-medical model interventions becomes increasingly necessary to ensure that all approaches to mental ill-health and distress are appropriately understood, funded and promoted. This enables CYP to make informed choices about the type of care they wish to receive and helps to prevent institutionalisation or iatrogenic harm.

A key finding reported by this review relates to acceptability of early intervention services. In general, acceptability was high, and CYP were satisfied with the treatment they had received; considering the low levels of satisfaction with CAMH services identified in a recent national survey (Care Quality Commission, 2023), this finding feels particularly relevant. Understanding why user satisfaction is a key feature of early intervention models such as the ones explored in this review would provide a basis for improving CAMH services, thus transforming CYP’s experiences of CAMHS care and increasing acceptability rates.

This review provides a solid foundation for future research, which should incorporate the use of co-production. More robust research involving the use of a control group is needed to understand the possible association between early intervention services and high service user satisfaction. Future research should address how successful models of care might be applied in diverse communities, exploring the impact of cultural and demographic factors on access and outcomes.

## Data Availability

N/A - systematic review

