## Appendix 1 for "Approaches to early intervention for common mental health problems in young people: a systematic review"

### Appendix 1: Search terms and overview of search results

Overview of search results

| Published literature databases |  |
| --- | --- |
| PsycINFO (via OVID) 1806 to January 30 2024 | 3938 |
| MEDLINE (via OVID) 1946 to January 30, 2024 | 2028 |
| Web of Science 1900 – 30/01/2024 | 2520 |
| Embase (via OVID) 1974 to 2024 January 30 | 2867 |
| CINAHL (via Ebsco) 30/01/2024 | 2168 |
| Total: | 13,521 |
| Deduplicated: | 9154 |
| Grey literature databases: |  |
| PsycEXTRA | 239 |
| HMIC | 217 |
| Total grey literature from databases: | 456 |
| Deduplicated: | 433 |
| Update search |  |
| PsycINFO (via OVID) January 2024-4^th^ December 2024 | 267 |
| MEDLINE (via OVID) January 2024-4^th^ December 2024 | 157 |
| Web of Science 1900 – January 2024-4^th^ December 2024 | 312 |
| Embase (via OVID) January 2024-4^th^ December 2024 | 349 |
| CINAHL (via Ebsco) January 2024-4^th^ December 2024 | 328 |
| PsycEXTRA January 2024-4^th^ December 2024 | 3 |
| HMIC January 2024-4^th^ December 2024 | 4 |
| Total: | 1420 |
| Deduplicated: | 1067 |

Search terms

**APA PsycInfo <1806 to January 30 2024>**

1	(child* or boy* or girl* or kids or minors or adolesc* or teen* or (young adj (people or patient* or male* or female* or men or wom#n or survivor* or minorit* or adult*)) or youth*).ti,id.	735596

2	("200" or "300" or "320").ag. (*wanted age groups)*	2553127

3	adolescent development/ or adolescent health/ or early adolescence/ or late adolescence/ or emerging adulthood/	68217

4	infant development/ or early childhood development/ or neonatal development/	40568

5	(neonat* or baby or babies or infant or infants or toddler* or nursery or preschool* or pre-school* or kindergarten*).ti,id,hw.	123567

6	nursery schools/ or nursery school students/ or preschool students/ or kindergartens/ or kindergarten students/	23653

7	(("100" or "120" or "140" or "160" or "180" or "340" or "360" or "380" or "390") not ("200" or "300" or "320")).ag.	(*unwanted age groups*) 310596

8	(parents of children with or parents of adolescents with).ti.	2114

9	4 or 5 or 6 or 7 or 8	378227

10	1 or 2 or 3	2882746

11	10 not (9 not (10 and 9))	2882746

12	affective disorders/ or "depression (emotion)"/ or endogenous depression/ or recurrent depression/ or treatment resistant depression/ or atypical depression/ or anxiety disorders/ or generalized anxiety disorder/ or obsessive compulsive disorder/ or panic disorder/ or postraumatic stress disorder/ or phobias/ or social phobia/ or emotional trauma/ or anxiety/ or panic attack/	184515

13	mental disorders/	98422

14	mental health/	94063

15	(mood disorder* or affective disorder* or depression or depressive or dysthymi* or anxiety disorder* or agoraphobia or obsess* or compulsi* or panic or phobi* or ptsd or posttrauma* or post trauma* or affective symptoms or ((mental* or psychologic*) adj (health or well*))).ti,id.	420031

16	12 or 13 or 14 or 15	573493

17	child psychiatry/ or adolescent psychiatry/ or Child Psychotherapy/ or adolescent psychotherapy/ or Child Psychology/ or Child Psychopathology/ or youth mental health/	29146

18	11 and 16	363757

19	17 or 18	382730

20	((pathway* adj3 (care or mental or psyc* or service* or model or referral* or help or contact*)) or ((helpseek* or help-seek* or help seek*) adj3 (contact* or experienc* or step* or delay or duration)) or ((Referral* or treatment or "mental health service*") adj2 (pattern* or delay)) or (contact adj3 (service* or professional*)) or "Journey of care" or "point* of entry" or (entry adj2 (care or service* or treatment)) or "early intervention" or ((integrat* or multidisciplinary or multi-disciplinary or coordinat* or co-ordinat*) adj2 (care or treatment or therapy or team* or services)) or "early support" or "improv* access" or ((fast or faster or early) adj2 (access or referral* or signpost* or support or "mental health" or treatment)) or "support hub*").ti,id.	21396

21	((pathway* adj3 (care or mental or psyc* or service* or model or referral* or help or contact*)) or ((helpseek* or help-seek* or help seek*) adj3 (contact* or experienc* or step* or delay or duration)) or ((Referral* or treatment or "mental health service*") adj2 (pattern* or delay)) or (contact adj3 (service* or professional*)) or "Journey of care" or "point* of entry" or (entry adj2 (care or service* or treatment)) or "early intervention" or ((integrat* or multidisciplinary or multi-disciplinary or coordinat* or co-ordinat*) adj2 (care or treatment or therapy or team* or services)) or "early support" or "improv* access" or ((fast or faster or early) adj2 (access or referral* or signpost* or support or "mental health" or treatment)) or "support hub*").ab. /freq=2	11758

22	early intervention/ or preventative mental health services/	12886

23	forensic psychology/ or forensic psychiatry/ or hospitalized patients/ or psychiatric hospitalization/	32470

24	(forensic or school* or inpatient* or compulsor* detention or compulsor* detained or prison* or jail*).ti,id.	289730

25	23 or 24	305917

26	20 or 21 or 22	33076

27	26 not 25	30717

28	19 and 27	4603

29	16 or 17	592195

30	("early intervention" or "early detection" or "early support").ti.	4684

31	30 not 25	4421

32	29 and 31	623

33	28 or 32	4865

34	("2400" or "2500" or "2600" or "2800" or "2900" or "3000" or "3200" or "3400" or "3500" or "2600" or "4200" or "4000" or "4600" or "5000" or "4800" or "3800").dt.	(*remove editorials, commentaries, dissertations, book chapters)*	1359680

35	33 not 34	3938

**Ovid MEDLINE(R) ALL <1946 to January 29, 2024>**

1	(child* or boy* or girl* or kids or minors or adolesc* or teen* or (young adj (people or patient* or male* or female* or men or wom#n or survivor* or minorit* or adult*)) or youth*).ti,kf.	1310133

2	(child* or adolescen* or p?ediatric*).jw.	760029

3	Child/ or minors/ or adolescent development/ or adolescent health/ or adolescent/	3183710

4	Infant/ or infant, newborn/	1263222

5	(neonat* or baby or babies or infant or infants or toddler* or nursery or preschool* or pre-school* or kindergarten*).ti,kf,hw.	1839711

6	Schools, nursery/ or child, preschool/	996265

7	("parents of children with" or "parents of adolescents with").ti.	2328

8	4 or 5 or 6 or 7	1841345

9	1 or 2 or 3	3851161

10	9 not (8 not (9 and 8))	3851161

11	Mood disorders/ or depressive disorder/ or depressive disorder, major/ or depressive disorder, treatment resistant/ or anxiety disorders/ or obsessive-compulsive disorder/ or panic disorder/ or phobic disorders/ or phobia, social/ or Stress Disorders, Post-Traumatic/	222042

12	mental disorders/	179581

13	mental health/	65186

14	(mood disorder* or affective disorder* or depression or depressive or dysthymi* or anxiety disorder* or agoraphobia or obsess* or compulsi* or panic or phobi* or ptsd or posttrauma* or post trauma* or affective symptoms or ((mental* or psychologic*) adj (health or well*))).ti,kf.	413181

15	11 or 12 or 13 or 14	661695

16	child psychiatry/ or adolescent psychiatry/ or Psychology, Child/	20402

17	10 and 15	156979

18	16 or 17	173788

19	((pathway* adj3 (care or mental or psyc* or service* or model or referral* or help or contact*)) or ((helpseek* or help-seek* or help seek*) adj3 (contact* or experienc* or step* or delay or duration)) or ((Referral* or treatment or "mental health service*") adj2 (pattern* or delay)) or (contact adj3 (service* or professional*)) or "Journey of care" or "point* of entry" or (entry adj2 (care or service* or treatment)) or "early intervention" or ((integrat* or multidisciplinary or multi-disciplinary or coordinat* or co-ordinat*) adj2 (care or treatment or therapy or team* or services)) or "early support" or "improv* access" or ((fast or faster or early) adj2 (access or referral* or signpost* or support or "mental health" or treatment)) or "support hub*").ti,kw.	41411

20	((pathway* adj3 (care or mental or psyc* or service* or model or referral* or help or contact*)) or ((helpseek* or help-seek* or help seek*) adj3 (contact* or experienc* or step* or delay or duration)) or ((Referral* or treatment or "mental health service*") adj2 (pattern* or delay)) or (contact adj3 (service* or professional*)) or "Journey of care" or "point* of entry" or (entry adj2 (care or service* or treatment)) or "early intervention" or ((integrat* or multidisciplinary or multi-disciplinary or coordinat* or co-ordinat*) adj2 (care or treatment or therapy or team* or services)) or "early support" or "improv* access" or ((fast or faster or early) adj2 (access or referral* or signpost* or support or "mental health" or treatment)) or "support hub*").ab. /freq=2	32687

21	forensic psychology/ or forensic psychiatry/ or hospitalization/	147465

22	(forensic or school* or inpatient* or compulsor* detention or compulsor* detained or prison* or jail*).ti,kf.	218056

23	21 or 22	354666

24	19 or 20	64595

25	24 not 23	62360

26	18 and 25	1757

27	15 or 16	678380

28	("early intervention" or "early detection" or "early support").ti.	16511

29	28 not 23	16259

30	27 and 29	605

31	26 or 30	2116

32	academic dissertation/ or anecdotes/ or "book review"/ or comment/ or congress/ or editorial/ or encyclopedia/ or letter/ or meeting abstract/	2288076

33	31 not 32	2028

**Web of Science 1900 – 29/01/2024**

\# Searches:

1: (TI=((child* OR boy* OR girl* OR kids OR minors OR adolesc* OR teen* OR (young NEAR/0 (people OR patient* OR male* OR female* OR men OR wom?n OR survivor* OR minorit* OR adult* )) OR youth* ))) OR TS=((child* OR boy* OR girl* OR kids OR minors OR adolesc* OR teen* OR (young NEAR/0 (people OR patient* OR male* OR female* OR men OR wom?n OR survivor* OR minorit* OR adult* )) OR youth* ))	Date Run: Mon Jan 29 2024 11:53:39 GMT+0000 (Greenwich Mean Time)	Results: 3748541

2: TI=(neonat* or baby or babies or infant or infants or toddler* or nursery or preschool* or pre-school* or kindergarten*) or TS=(neonat* or baby or babies or infant or infants or toddler* or nursery or preschool* or pre-school* or kindergarten*)	Date Run: Mon Jan 29 2024 11:56:08 GMT+0000 (Greenwich Mean Time)	Results: 941401

3: TI=("parents of children with" or "parents of adolescents with")	Date Run: Mon Jan 29 2024 11:57:12 GMT+0000 (Greenwich Mean Time)	Results: 3358

4: (TI=( (mood disorder* or affective disorder* or depression or depressive or dysthymi* or anxiety disorder* or agoraphobia or obsess* or compulsi* or panic or phobi* or ptsd or posttrauma* or post trauma* or affective symptoms or ((mental* or psychologic*) adj (health or well*))))) OR TS=( (mood disorder* or affective disorder* or depression or depressive or dysthymi* or anxiety disorder* or agoraphobia or obsess* or compulsi* or panic or phobi* or ptsd or posttrauma* or post trauma* or affective symptoms or ((mental* or psychologic*) adj (health or well*))))	Date Run: Mon Jan 29 2024 11:57:43 GMT+0000 (Greenwich Mean Time)	Results: 1462049

5: TI=((pathway* NEAR/3 (care OR mental OR psyc* OR service* OR model OR referral* OR help OR contact* )) OR ((helpseek* OR help-seek* OR "help seek*") NEAR/3 (contact* OR experienc* OR step* OR delay OR duration )) OR ((Referral* OR treatment OR "mental health service*" ) NEAR/2 (pattern* OR delay )) OR (contact NEAR/3 (service* OR professional* )) OR "Journey of care" OR "point* of entry" OR (entry NEAR/2 (care OR service* OR treatment)) OR "early intervention" OR ((integrat* OR multidisciplinary OR multi-disciplinary OR coordinat* OR co-ordinat* ) NEAR/2 (care OR treatment OR therapy OR team* OR services )) OR "early support" OR "improv* access" OR ((fast OR faster OR early ) NEAR/2 (access OR referral* OR signpost* OR support OR "mental health" OR treatment )) OR "support hub*")	Date Run: Mon Jan 29 2024 12:03:21 GMT+0000 (Greenwich Mean Time)	Results: 82312

6: TI=(forensic or school* or inpatient* or compulsor* detention or compulsor* detained or prison* or jail*) OR TS=(forensic or school* or inpatient* or compulsor* detention or compulsor* detained or prison* or jail*)	Date Run: Mon Jan 29 2024 12:10:09 GMT+0000 (Greenwich Mean Time)	Results: 1231020

7: TI=("early intervention" or "early detection" or "early support")	Date Run: Mon Jan 29 2024 12:10:29 GMT+0000 (Greenwich Mean Time)	Results: 25045

8: #1 not ((#2 or #3) not ((#2 or #3) and #1))	Date Run: Mon Jan 29 2024 13:03:24 GMT+0000 (Greenwich Mean Time)	Results: 3748541

9: #5 not (#6 not (#5 and #6))	Date Run: Mon Jan 29 2024 13:04:01 GMT+0000 (Greenwich Mean Time)	Results: 82312

10: (#4 and #8 and #9) or (#4 and #7)	Date Run: Mon Jan 29 2024 13:05:05 GMT+0000 (Greenwich Mean Time)	Results: 3312

11: Limit #10 to Document Type: Article	Results: 2,520

**Embase <1974 to 2024 January 29>**

1	(child* or boy* or girl* or kids or minors or adolesc* or teen* or (young adj (people or patient* or male* or female* or men or wom#n or survivor* or minorit* or adult*)) or youth*).ti,kf.	1521360

2	(child* or adolescen* or p?ediatric*).jn.	359918

3	child/ or adolescent/ or young adult/	3453234

4	Infant/ or newborn/ or baby/	1155839

5	(neonat* or baby or babies or infant or infants or toddler* or nursery or preschool* or pre-school* or kindergarten*).ti,kf,hw.	1429159

6	nursery school/ or preschool child/	632969

7	("parents of children with" or "parents of adolescents with").ti.	3037

8	4 or 5 or 6 or 7	1700020

9	1 or 2 or 3	3950999

10	9 not (8 not (9 and 8))	3950999

11	mood disorder/ or depression/ or major affective disorder/ or depression/ or adolescent depression/ or endogenous depression/ or major depression/ or minor depression/ or "mixed anxiety and depression"/ or recurrent brief depression/ or treatment resistant depression/ or anxiety disorder/ or generalized anxiety disorder/ or panic/ or phobia/ or posttraumatic stress disorder/ or obsessive compulsive disorder/ or social anxiety/	725510

12	mental disease/	285051

13	mental health/ or psychological wellbeing/	243976

14	(mood disorder* or affective disorder* or depression or depressive or dysthymi* or anxiety disorder* or agoraphobia or obsess* or compulsi* or panic or phobi* or ptsd or posttrauma* or post trauma* or affective symptoms or ((mental* or psychologic*) adj (health or well*))).ti,kf.	517947

15	11 or 12 or 13 or 14	1210184

16	child psychiatry/ or child psychology/	38621

17	10 and 15	236553

18	16 or 17	263321

19	((pathway* adj3 (care or mental or psyc* or service* or model or referral* or help or contact*)) or ((helpseek* or help-seek* or help seek*) adj3 (contact* or experienc* or step* or delay or duration)) or ((Referral* or treatment or "mental health service*") adj2 (pattern* or delay)) or (contact adj3 (service* or professional*)) or "Journey of care" or "point* of entry" or (entry adj2 (care or service* or treatment)) or "early intervention" or ((integrat* or multidisciplinary or multi-disciplinary or coordinat* or co-ordinat*) adj2 (care or treatment or therapy or team* or services)) or "early support" or "improv* access" or ((fast or faster or early) adj2 (access or referral* or signpost* or support or "mental health" or treatment)) or "support hub*").ti,kw.	60459

20	((pathway* adj3 (care or mental or psyc* or service* or model or referral* or help or contact*)) or ((helpseek* or help-seek* or help seek*) adj3 (contact* or experienc* or step* or delay or duration)) or ((Referral* or treatment or "mental health service*") adj2 (pattern* or delay)) or (contact adj3 (service* or professional*)) or "Journey of care" or "point* of entry" or (entry adj2 (care or service* or treatment)) or "early intervention" or ((integrat* or multidisciplinary or multi-disciplinary or coordinat* or co-ordinat*) adj2 (care or treatment or therapy or team* or services)) or "early support" or "improv* access" or ((fast or faster or early) adj2 (access or referral* or signpost* or support or "mental health" or treatment)) or "support hub*").ab. /freq=2	55294

21	forensic psychiatry/ or forensic psychology/ or hospital patient/ or hospitalized adolescent/ or hospitalized child/ or hospitalized infant/	258414

22	(forensic or school* or inpatient* or compulsor* detention or compulsor* detained or prison* or jail*).ti,kf.	252702

23	21 or 22	471396

24	19 or 20	100334

25	24 not 23	95639

26	18 and 25	3259

27	15 or 16	1235725

28	("early intervention" or "early detection" or "early support").ti.	22652

29	28 not 23	22263

30	27 and 29	1395

31	26 or 30	4137

32	("Books" or "Chapter" or "conference abstract" or "Conference Review" or "editorial" or "letter").pt.	7213644

33	31 not 32	2867

**CINAHL Searched 30/01/2024**

| S32 | S31 | Limiters - **Exclude MEDLINE records**  Expanders - Apply equivalent subjects  Search modes - Boolean/Phrase | Interface - EBSCOhost Research Databases  Search Screen - Advanced Search  Database - CINAHL Plus | 2,168 |
| --- | --- | --- | --- | --- |
| S31 | S29 NOT S30 | Expanders - Apply equivalent subjects  Search modes - Boolean/Phrase | Interface - EBSCOhost Research Databases  Search Screen - Advanced Search  Database - CINAHL Plus | 4,030 |
| S30 | s29 | **Limiters - Publication Type: Abstract, Book, Book Chapter, Book Review, Commentary, Doctoral Dissertation, Editorial, Interview, Letter, Masters Thesis**  Expanders - Apply equivalent subjects  Search modes - Boolean/Phrase | Interface - EBSCOhost Research Databases  Search Screen - Advanced Search  Database - CINAHL Plus | 209 |
| S29 | S24 OR S28 | Expanders - Apply equivalent subjects  Search modes - Boolean/Phrase | Interface - EBSCOhost Research Databases  Search Screen - Advanced Search  Database - CINAHL Plus | 4,239 |
| S28 | S25 AND S27 | Expanders - Apply equivalent subjects  Search modes - Boolean/Phrase | Interface - EBSCOhost Research Databases  Search Screen - Advanced Search  Database - CINAHL Plus | Display |
| S27 | S26 NOT S21 | Expanders - Apply equivalent subjects  Search modes - Boolean/Phrase | Interface - EBSCOhost Research Databases  Search Screen - Advanced Search  Database - CINAHL Plus | Display |
| S26 | TI ("early intervention" or "early detection" or "early support") | Expanders - Apply equivalent subjects  Search modes - Boolean/Phrase | Interface - EBSCOhost Research Databases  Search Screen - Advanced Search  Database - CINAHL Plus | Display |
| S25 | S14 OR S15 | Expanders - Apply equivalent subjects  Search modes - Boolean/Phrase | Interface - EBSCOhost Research Databases  Search Screen - Advanced Search  Database - CINAHL Plus | Display |
| S24 | S22 AND S23 | Expanders - Apply equivalent subjects  Search modes - Boolean/Phrase | Interface - EBSCOhost Research Databases  Search Screen - Advanced Search  Database - CINAHL Plus | 4,067 |
| S23 | S15 OR S16 | Expanders - Apply equivalent subjects  Search modes - Boolean/Phrase | Interface - EBSCOhost Research Databases  Search Screen - Advanced Search  Database - CINAHL Plus | Display |
| S22 | S18 NOT S21 | Expanders - Apply equivalent subjects  Search modes - Boolean/Phrase | Interface - EBSCOhost Research Databases  Search Screen - Advanced Search  Database - CINAHL Plus | 189,801 |
| S21 | S19 OR S20 | Expanders - Apply equivalent subjects  Search modes - Boolean/Phrase | Interface - EBSCOhost Research Databases  Search Screen - Advanced Search  Database - CINAHL Plus | Display |
| S20 | TI (forensic or school* or inpatient* or compulsor* detention or compulsor* detained or prison* or jail*) | Expanders - Apply equivalent subjects  Search modes - Boolean/Phrase | Interface - EBSCOhost Research Databases  Search Screen - Advanced Search  Database - CINAHL Plus | Display |
| S19 | (MH "Forensic Psychiatry") OR (MH "Mentally Ill Offenders") OR (MH "Forensic Psychology") | Expanders - Apply equivalent subjects  Search modes - Boolean/Phrase | Interface - EBSCOhost Research Databases  Search Screen - Advanced Search  Database - CINAHL Plus | Display |
| S18 | TI ( ((pathway* N3 (care or mental or psyc* or service* or model or referral* or help or contact*)) or ((helpseek* or help-seek* or help seek*) N3 (contact* or experienc* or step* or delay or duration)) or ((Referral* or treatment or "mental health service*") N2 (pattern* or delay)) or (contact N3 (service* or professional*)) or "Journey of care" or "point* of entry" or (entry N2 (care or service* or treatment)) or "early intervention" or ((integrat* or multidisciplinary or multi-disciplinary or coordinat* or co-ordinat*) N2 (care or treatment or therapy or team* or services)) or "early support" or "improv* access" or ((fast or faster or early) N2 (access or referral* or signpost* or support or "mental health" or treatment)) or "support hub*") ) OR MW ( ((pathway* N3 (care or mental or psyc* or service* or model or referral* or help or contact*)) or ((helpseek* or help-seek* or help seek*) N3 (contact* or experienc* or step* or delay or duration)) or ((Referral* or treatment or "mental health service*") N2 (pattern* or delay)) or (contact N3 (service* or professional*)) or "Journey of care" or "point* of entry" or (entry N2 (care or service* or treatment)) or "early intervention" or ((integrat* or multidisciplinary or multi-disciplinary or coordinat* or co-ordinat*) N2 (care or treatment or therapy or team* or services)) or "early support" or "improv* access" or ((fast or faster or early) N2 (access or referral* or signpost* or support or "mental health" or treatment)) or "support hub*") ) or AB ( ((pathway* N3 (care or mental or psyc* or service* or model or referral* or help or contact*)) or ((helpseek* or help-seek* or help seek*) N3 (contact* or experienc* or step* or delay or duration)) or ((Referral* or treatment or "mental health service*") N2 (pattern* or delay)) or (contact N3 (service* or professional*)) or "Journey of care" or "point* of entry" or (entry N2 (care or service* or treatment)) or "early intervention" or ((integrat* or multidisciplinary or multi-disciplinary or coordinat* or co-ordinat*) N2 (care or treatment or therapy or team* or services)) or "early support" or "improv* access" or ((fast or faster or early) N2 (access or referral* or signpost* or support or "mental health" or treatment)) or "support hub*") ) | Expanders - Apply equivalent subjects  Search modes - Boolean/Phrase | Interface - EBSCOhost Research Databases  Search Screen - Advanced Search  Database - CINAHL Plus | 193,253 |
| S17 | S15 OR S16 | Expanders - Apply equivalent subjects  Search modes - Boolean/Phrase | Interface - EBSCOhost Research Databases  Search Screen - Advanced Search  Database - CINAHL Plus | Display |
| S16 | S9 AND S14 | Expanders - Apply equivalent subjects  Search modes - Boolean/Phrase | Interface - EBSCOhost Research Databases  Search Screen - Advanced Search  Database - CINAHL Plus | Display |
| S15 | (MH "Child Psychiatry") OR (MH "Adolescent Psychology") OR (MH "Child Psychology") | Expanders - Apply equivalent subjects  Search modes - Boolean/Phrase | Interface - EBSCOhost Research Databases  Search Screen - Advanced Search  Database - CINAHL Plus | Display |
| S14 | S10 OR S11 OR S12 OR S13 | Expanders - Apply equivalent subjects  Search modes - Boolean/Phrase | Interface - EBSCOhost Research Databases  Search Screen - Advanced Search  Database - CINAHL Plus | Display |
| S13 | TI ("mood disorder*" or "affective disorder*" or depression or depressive or dysthymi* or "anxiety disorder*" or agoraphobia or obsess* or compulsi* or panic or phobi* or ptsd or posttrauma* or "post trauma*" or "affective symptoms" or ((mental* or psychologic*) N (health or well*))) | Expanders - Apply equivalent subjects  Search modes - Boolean/Phrase | Interface - EBSCOhost Research Databases  Search Screen - Advanced Search  Database - CINAHL Plus | Display |
| S12 | (MH "Mental Health") | Expanders - Apply equivalent subjects  Search modes - Boolean/Phrase | Interface - EBSCOhost Research Databases  Search Screen - Advanced Search  Database - CINAHL Plus | Display |
| S11 | (MH "Mental Disorders") OR (MH "Mental Disorders, Chronic") OR (MH "Social Anxiety Disorders") | Expanders - Apply equivalent subjects  Search modes - Boolean/Phrase | Interface - EBSCOhost Research Databases  Search Screen - Advanced Search  Database - CINAHL Plus | Display |
| S10 | (MH "Affective Disorders") OR (MH "Depression") OR (MH "Anxiety Disorders") OR (MH "Generalized Anxiety Disorder") OR (MH "Obsessive-Compulsive Disorder") OR (MH "Panic Disorder") OR (MH "Phobic Disorders") OR (MH "Social Anxiety Disorders") OR (MH "Stress Disorders, Post-Traumatic") | Expanders - Apply equivalent subjects  Search modes - Boolean/Phrase | Interface - EBSCOhost Research Databases  Search Screen - Advanced Search  Database - CINAHL Plus | Display |
| S9 | S8 NOT S7 | Expanders - Apply equivalent subjects  Search modes - Boolean/Phrase | Interface - EBSCOhost Research Databases  Search Screen - Advanced Search  Database - CINAHL Plus | Display |
| S8 | S1 OR S2 | Expanders - Apply equivalent subjects  Search modes - Boolean/Phrase | Interface - EBSCOhost Research Databases  Search Screen - Advanced Search  Database - CINAHL Plus | Display |
| S7 | S3 OR S4 OR S5 OR S6 | Expanders - Apply equivalent subjects  Search modes - Boolean/Phrase | Interface - EBSCOhost Research Databases  Search Screen - Advanced Search  Database - CINAHL Plus | Display |
| S6 | TI ("parents of children with" or "parents of adolescents with") | Expanders - Apply equivalent subjects  Search modes - Boolean/Phrase | Interface - EBSCOhost Research Databases  Search Screen - Advanced Search  Database - CINAHL Plus | Display |
| S5 | (MH "Schools, Nursery") | Expanders - Apply equivalent subjects  Search modes - Boolean/Phrase | Interface - EBSCOhost Research Databases  Search Screen - Advanced Search  Database - CINAHL Plus | Display |
| S4 | TI (neonat* or baby or babies or infant or infants or toddler* or nursery or preschool* or pre-school* or kindergarten*) | Expanders - Apply equivalent subjects  Search modes - Boolean/Phrase | Interface - EBSCOhost Research Databases  Search Screen - Advanced Search  Database - CINAHL Plus | Display |
| S3 | (MH "Infant") OR (MH "Infant, Newborn") OR (MH "Child, Preschool") | Expanders - Apply equivalent subjects  Search modes - Boolean/Phrase | Interface - EBSCOhost Research Databases  Search Screen - Advanced Search  Database - CINAHL Plus | Display |
| S2 | (MH "Adolescence") OR (MH "Child") OR (MH "Minors (Legal)") OR (MH "Young Adult") | Expanders - Apply equivalent subjects  Search modes - Boolean/Phrase | Interface - EBSCOhost Research Databases  Search Screen - Advanced Search  Database - CINAHL Plus | Display |
| S1 | (child* or boy* or girl* or kids or minors or adolesc* or teen* or (young N1 (people or patient* or male* or female* or men or woman or women or survivor* or minorit* or adult*)) or youth*) | Expanders - Apply equivalent subjects  Search modes - Boolean/Phrase | Interface - EBSCOhost Research Databases  Search Screen - Advanced Search  Database - CINAHL Plus | Display |

**HMIC Health Management Information Consortium <1979 to November 2023>**

1	(child* or boy* or girl* or kids or minors or adolesc* or teen* or (young adj (people or patient* or male* or female* or men or wom#n or survivor* or minorit* or adult*)) or youth*).ti,hw.	35651

2	Early teenagers/ or late teenagers/ or young people/	11146

3	infants/ or neonates/ or toddlers/	1764

4	(neonat* or baby or babies or infant or infants or toddler* or nursery or preschool* or pre-school* or kindergarten*).ti,hw.	5030

5	nursery schools/ or nursery centres/ or day nurseries/ or nurseries/ or pre school education/	229

6	(parents of children with or parents of adolescents with).ti.	37

7	1 or 2	35651

8	3 or 4 or 5 or 6	5140

9	7 not (8 not (7 and 8))	35651

10	exp mental disorders/ or exp Mental health/ or exp Anxiety/ or exp Mental illness/ or exp Phobias/ or exp Depression/	28751

11	(mood disorder* or affective disorder* or depression or depressive or dysthymi* or anxiety disorder* or agoraphobia or obsess* or compulsi* or panic or phobi* or ptsd or posttrauma* or post trauma* or affective symptoms or ((mental* or psychologic*) adj (health or well*))).ti,hw.	24048

12	10 or 11	38887

13	child psychiatry/ or child mental health services/ or child psychotherapy/ or adolescent psychiatry/	234

14	9 and 12	4650

15	13 or 14	4710

16	((pathway* adj3 (care or mental or psyc* or service* or model or referral* or help or contact*)) or ((helpseek* or help-seek* or help seek*) adj3 (contact* or experienc* or step* or delay or duration)) or ((Referral* or treatment or "mental health service*") adj2 (pattern* or delay)) or (contact adj3 (service* or professional*)) or "Journey of care" or "point* of entry" or (entry adj2 (care or service* or treatment)) or "early intervention" or ((integrat* or multidisciplinary or multi-disciplinary or coordinat* or co-ordinat*) adj2 (care or treatment or therapy or team* or services)) or "early support" or "improv* access" or ((fast or faster or early) adj2 (access or referral* or signpost* or support or "mental health" or treatment)) or "support hub*").ti,hw.	6841

17	((pathway* adj3 (care or mental or psyc* or service* or model or referral* or help or contact*)) or ((helpseek* or help-seek* or help seek*) adj3 (contact* or experienc* or step* or delay or duration)) or ((Referral* or treatment or "mental health service*") adj2 (pattern* or delay)) or (contact adj3 (service* or professional*)) or "Journey of care" or "point* of entry" or (entry adj2 (care or service* or treatment)) or "early intervention" or ((integrat* or multidisciplinary or multi-disciplinary or coordinat* or co-ordinat*) adj2 (care or treatment or therapy or team* or services)) or "early support" or "improv* access" or ((fast or faster or early) adj2 (access or referral* or signpost* or support or "mental health" or treatment)) or "support hub*").ab. /freq=2	2059

18	forensic psychiatry/ or hospitalisation/ or patient admission/	2507

19	(forensic or school* or inpatient* or compulsor* detention or compulsor* detained or prison* or jail*).ti,hw.	7942

20	18 or 19	10172

21	16 or 17	7643

22	21 not 20	7424

23	15 and 22	142

24	12 or 13	38947

25	("early intervention" or "early detection" or "early support").ti.	236

26	25 not 20	224

27	24 and 26	101

28	23 or 27	217

**APA PsycExtra <1908 to March 11, 2024>**

1	(child* or boy* or girl* or kids or minors or adolesc* or teen* or (young adj (people or patient* or male* or female* or men or wom#n or survivor* or minorit* or adult*)) or youth*).ti,id.	40807

2	("200" or "300" or "320").ag.	102368

3	adolescent development/ or adolescent health/ or early adolescence/ or late adolescence/ or emerging adulthood/	3726

4	infant development/ or early childhood development/ or neonatal development/	1974

5	(neonat* or baby or babies or infant or infants or toddler* or nursery or preschool* or pre-school* or kindergarten*).ti,id,hw.	4172

6	nursery schools/ or nursery school students/ or preschool students/ or kindergartens/ or kindergarten students/	1091

7	(("100" or "120" or "140" or "160" or "180" or "340" or "360" or "380" or "390") not ("200" or "300" or "320")).ag.	13106

8	(parents of children with or parents of adolescents with).ti.	60

9	4 or 5 or 6 or 7 or 8	15237

10	1 or 2 or 3	123843

11	10 not (9 not (10 and 9))	123843

12	affective disorders/ or "depression (emotion)"/ or endogenous depression/ or recurrant depression/ or treatment resistant depression/ or atypical depression/ or anxiety disorders/ or generalized anxiety disorder/ or obsessive compulsive disorder/ or panic disorder/ or postraumatic stress disorder/ or phobias/ or social phobia/ or emotional trauma/ or anxiety/ or panic attack/	6159

13	mental disorders/	4165

14	mental health/	6824

15	(mood disorder* or affective disorder* or depression or depressive or dysthymi* or anxiety disorder* or agoraphobia or obsess* or compulsi* or panic or phobi* or ptsd or posttrauma* or post trauma* or affective symptoms or ((mental* or psychologic*) adj (health or well*))).ti,id.	25475

16	12 or 13 or 14 or 15	31473

17	child psychiatry/ or adolescent psychiatry/ or Child Psychotherapy/ or adolescent psychotherapy/ or Child Psychology/ or Child Psychopathology/ or youth mental health/	1661

18	11 and 16	13908

19	17 or 18	15017

20	((pathway* adj3 (care or mental or psyc* or service* or model or referral* or help or contact*)) or ((helpseek* or help-seek* or help seek*) adj3 (contact* or experienc* or step* or delay or duration)) or ((Referral* or treatment or "mental health service*") adj2 (pattern* or delay)) or (contact adj3 (service* or professional*)) or "Journey of care" or "point* of entry" or (entry adj2 (care or service* or treatment)) or "early intervention" or ((integrat* or multidisciplinary or multi-disciplinary or coordinat* or co-ordinat*) adj2 (care or treatment or therapy or team* or services)) or "early support" or "improv* access" or ((fast or faster or early) adj2 (access or referral* or signpost* or support or "mental health" or treatment)) or "support hub*").ti,id.	1524

21	((pathway* adj3 (care or mental or psyc* or service* or model or referral* or help or contact*)) or ((helpseek* or help-seek* or help seek*) adj3 (contact* or experienc* or step* or delay or duration)) or ((Referral* or treatment or "mental health service*") adj2 (pattern* or delay)) or (contact adj3 (service* or professional*)) or "Journey of care" or "point* of entry" or (entry adj2 (care or service* or treatment)) or "early intervention" or ((integrat* or multidisciplinary or multi-disciplinary or coordinat* or co-ordinat*) adj2 (care or treatment or therapy or team* or services)) or "early support" or "improv* access" or ((fast or faster or early) adj2 (access or referral* or signpost* or support or "mental health" or treatment)) or "support hub*").ab. /freq=2	481

22	early intervention/ or preventative mental health services/	726

23	forensic psychology/ or forensic psychiatry/ or hospitalized patients/ or psychiatric hospitalization/	1510

24	(forensic or school* or inpatient* or compulsor* detention or compulsor* detained or prison* or jail*).ti,id.	22710

25	23 or 24	23395

26	20 or 21 or 22	2044

27	26 not 25	1853

28	19 and 27	216

29	16 or 17	32571

30	("early intervention" or "early detection" or "early support").ti.	230

31	30 not 25	215

32	29 and 31	37

33	28 or 32	239
