## Appendix 2 for "Approaches to early intervention for common mental health problems in young people: a systematic review"

### Appendix 2a: adaptation of scoring system

| RCT | | |
| --- | --- | --- |
| 1 | Did the trial address a clearly focused issue? | 1=statement of aim 2=statement of aim which includes specific population, intervention, and outcomes of interest |
| 2 | Was the assignment of patients to treatments  randomised? | 1=state randomly assigned 2=provide clear description of method of randomisation |
| 3 | Were patients, health workers and study  personnel blinded? | 1= interviewers blinded but participants not 2= participants also blinded (note: this is unlikely) |
| 4 | Were the groups similar at the start of the trial? | 1= differences but unlikely to impact effect of intervention or outcomes 2=no differences |
| 5 | Aside from the experimental intervention,  were the groups treated equally? | 1=few differences, and those present unlikely to affect outcomes  2= nothing to suggest that other aspects of participant experience were different |
| 6 | Were all of the patients who entered  the trial properly accounted for at its conclusion? | 1=not all accounted for but similar numbers of drop out in each arm 2= ITT analysis and no crossover |
| 7 | How large was the treatment effect? | 1= clear reporting of outcomes but less clear on primary outcomes 2= clear statement of primary outcomes and these reported, clear reporting of the effect of the intervention |
| 8 | How precise was the estimate of the treatment effect? | 1: confidence intervals reported where relevant, range reported for n %) 2: confidence intervals or range suggest that there is little variation and sample size is sufficient - >200 people |
| 9 | Can the results be applied in your context? (or to the local population?) | 1= narrow group so would need to extrapolate to our target population, but would likely react to intervention in similar way 2= sample and intervention setting very similar to our target group |
| 10 | Were all clinically important outcomes considered? | 1= clinical outcomes relevant to problem 2= both clinical (relating to mental health problem of interest) and social/functional outcomes considered |
| 11 | Are the benefits worth the harms and costs? | 1= limited change in any measures of problem of interest, but no harms 2=no harms reported and improvement in an aspect |
| Cohort | | |
| 1 | Did the study address a clearly focused issue? | 1=statement of aim 2=statement of aim which includes specific population, intervention, and outcomes of interest |
| 2 | Was the cohort or sample recruited in an acceptable way? | 1= cohort is representative of the target population defined in the study, and wasn’t a more specific special sub-group 2= and recruitment process meant that people who should have been included were |
| 3 | Was the exposure/intervention/service receipt accurately measured to  minimise bias? | 1= describe the intervention well 2= report the extent that participants used the services |
| 4 | Was the outcome accurately measured to  minimise bias? | 2= validated outcome measures, or routine outcome monitoring/health record data mark down to 1 if reason to suspect aspects of the above are less accurate e.g. Subscales or if large number of non-validated outcomes also reported |
| 5a | Have the authors identified all important confounding factors? | 1= age, gender, ethnicity (where relevant) 2= additionally baseline severity of symptoms (or where relevant, other items you think are equally as relevant for the research question) |
| 5b | Have they taken account of the confounding factors in the design and/or analysis? | 1= include in some analyses 2= include in all analyses where relevant |
| 6a | Was the follow up of subjects complete  enough? (>=80%) | 1= <80% followed up but this is similar in both groups (when control present) or if pre-post, >60% followed up or authors discuss reasons for drop-out and it seems unrelated to intervention or outcomes 2= >= 80% followed up |
| 6b | Was the follow up of subjects long  enough? | 1= 3+ months 2=6+ months |
| 7 | What are the results of this study? | 1= some statement linking the impact of the intervention on outcomes 2= clear statement of the extent of impact of the intervention on outcomes |
| 8 | How precise are the results? | 1= confidence intervals reported where relevant, range reported for n %) 2= confidence intervals or range suggest that there is little variation and sample size is sufficient - >200 people |
| 9 | Do you believe the results? | 1= no contemporaneous control but reason to believe results are true 2= contemporaneous control + no other concerns regarding validity e.g. Low fidelity, not full reporting, no consideration of confounders |
| 10 | Can the results be applied to the local population? | 0= no information or a group likely to respond very differently to the support e.g. Significant comorbidities of specific living situations which cause additional risk 1= narrow group so would need to extrapolate to our target population, but would likely react to intervention in similar way or age range of population extends outside of our target group or some indication that some participants may have more enduring problems also 2= sample and intervention setting very similar to our target group |
| 12 | What are the implications of this study for practice? | 0= no mention of implications for clinical practice 1= mentioned but unfounded 2= mentioned and supported by the evidence found in the study |
| Case-control | | |
| 1 | Did the study address a clearly focused issue? | 1=statement of aim 2=statement of aim which includes specific population, intervention, and outcomes of interest |
| 2 | Did the authors use an appropriate method  to answer their question? | 1= case-control method is appropriate  2= clear rationale described |
| 3 | Were the cases recruited in an acceptable way? | 1= cases representative of the target population defined in the study, and wasn’t a more specific special sub-group 2= and recruitment process meant that people who should have been included were |
| 4 | Were the controls selected in an  acceptable way? | 2= controls recruited in the same way that cases were |
| 5 | Was the exposure accurately measured to minimise bias? | 1: describe the intervention well 2: report the extent that participants used the services |
| 6a | (a) what confounding factors have the  authors accounted for? | 1. age, gender, ethnicity (where relevant) 2. additionally, baseline severity of symptoms (or where relevant, other items you think are equally as relevant for the research question) |
| 6b | (b) have the authors taken account  of the potential confounding factors in the design and/or in their analysis? | 1. include in some analyses 2. include in all analyses where relevant |
| 7 | What are the results of this study? | 1= some statement linking the impact of the intervention on outcomes 2= clear statement of the extent of impact of the intervention on outcomes |
| 8 | How precise are the results? How precise is the estimate of risk? | 1= confidence intervals reported where relevant, range reported for n %) 2= confidence intervals or range suggest that there is little variation and sample size is sufficient - >200 people |
| 9 | Do you believe the results? | 1= no contemporaneous control but reason to believe results are true 2= contemporaneous control + no other concerns regarding validity e.g. Low fidelity, not full reporting, no consideration of confounders |
| 10 | Can the results be applied to the local  population? | 0= no information or a group likely to respond very differently to the support e.g. Significant comorbidities of specific living situations which cause additional risk 1= narrow group so would need to extrapolate to our target population, but would likely react to intervention in similar way or age range of population extends outside of our target group or some indication that some participants may have more enduring problems also 2= sample and intervention setting very similar to our target group |

### Appendix 2b : Quality appraisal of included studies

**Quality Appraisal of Randomised Controlled Trial (RCT) Studies**

| Paper | Q1 | Q2 | Q3 | Q4 | Q5 | Q6 | Q7 | Q8 | Q9 | Q10 | Q11 | Total Score (out of 22) |
| --- | --- | --- | --- | --- | --- | --- | --- | --- | --- | --- | --- | --- |
| Arsarnow (2005; 2009) | 2 | 2 | 1 | 2 | 2 | 2 | 2 | 2 | 2 | 1 | 1 | 19 |
| Rapp (2017) [secondary analysis of Arsarnow 2005; 2009] | 1 | 2 | 1 | 2 | 2 | 2 | 1 | 1 | 2 | 0 | 2 | 16 |
| Ngo (2010) [secondary analysis of Arsarnow 2005; 2009] | 2 | 2 | 1 | 0 | 2 | 2 | 1 | 1 | 1 | 1 | 2 | 15 |
| Wells (2012) [secondary analysis of Arsarnow 2005; 2009] | 2 | 2 | 0 | 0 | 2 | 2 | 0 | 2 | 1 | 0 | 2 | 13 |
| Richardson (2014) | 2 | 2 | 1 | 2 | 2 | 2 | 2 | 2 | 2 | 2 | 2 | 21 |
| Martinez (2018) | 2 | 2 | 1 | 1 | 2 | 2 | 2 | 2 | 1 | 1 | 1 | 17 |
| Smith (2012) | 2 | 1 | 1 | 2 | 2 | 0 | 1 | 1 | 0 | 2 | 1 | 13 |
| Sheidow (2021) | 1 | 2 | 1 | 2 | 2 | 2 | 1 | 1 | 1 | 2 | 2 | 17 |
| Kidd (2020) | 1 | 2 | 0 | 1 | 2 | 2 | 2 | 1 | 0 | 2 | 1 | 14 |
| Sterling (2018) | 1 | 1 | 0 | 1 | 1 | 1 | 2 | 0 | 1 | 1 | 2 | 11 |
| Parthasarathy (2021) [subsample of sterling 2018] | 2 | 2 | 0 | 2 | 1 | 2 | 2 | 2 | 1 | 1 | 2 | 17 |
| Note. Q1 = Did the trial address a clearly focused issue?^a^; Q2 = Was the assignment of patients to treatments randomised?^a^ ;Q3 = Were patients, health workers and study personnel blinded?^b^; Q4 = Were the groups similar at the start of the trial?^b^ ; Q5 = Aside from the experimental intervention, were the groups treated equally?^b^ ; Q6 = Were all of the patients who entered the trial properly accounted for at its conclusion?^a^ ; Q7 = How large was the treatment effect?^c^ ; Q8 = How precise was the estimate of the treatment effect?^c^ ;Q9 = Can the results be applied in your context? (or to the local population?)^d^ ; Q10 = Were all clinically important outcomes considered?^c^ ;Q11 = Are the benefits worth the harms and costs?^c^ | | | | | | | | | | | | |
| 2 = Criterion is completely met ; 1 = Criterion is partially met ; 0 = Criterion not applicable, not met, or not mentioned | | | | | | | | | | | | |

^a:^ Validity of the study design for randomised controlled trials

^b^ : Methodological soundness of the study

^c:^ Quality of the results

^d^ Extent that results will help locally

**Quality Appraisal of Cohort Studies**

| Paper | Q1 | Q2 | Q3 | Q4 | Q5a | Q5b | Q6a | Q6b | Q7 | Q8 | Q9 | Q10 | Q12 | Total Score (out of 26) |
| --- | --- | --- | --- | --- | --- | --- | --- | --- | --- | --- | --- | --- | --- | --- |
| Corley (2023) | 2 | 2 | 1 | 2 | 2 | 2 | 2 | 0 | 2 | 0 | 1 | 2 | 1 | 19 |
| Parry (2023) | 2 | 2 | 1 | 2 | 0 | 0 | 1 | 0 | 1 | 1 | 0 | 2 | 1 | 13 |
| Schley (2008) | 2 | 1 | 1 | 2 | 0 | 0 | 2 | 2 | 2 | 0 | 1 | 1 | 1 | 15 |
| O'Dell (2020) | 1 | 2 | 2 | 2 | 2 | 2 | 0 | 0 | 2 | 1 | 1 | 1 | 2 | 18 |
| Chang (2023) | 1 | 2 | 1 | 2 | 0 | 0 | 0 | 0 | 1 | 0 | 0 | 1 | 1 | 9 |
| Khazanov (2024) | 2 | 2 | 1 | 2 | 2 | 2 | 1 | 0 | 2 | 0 | 2 | 1 | 2 | 19 |
| O'Keeffe (2015) | 2 | 2 | 1 | 2 | 1 | 1 | 0 | 0 | 1 | 2 | 1 | 2 | 1 | 16 |
| Saxe (2012) | 2 | 1 | 1 | 2 | 1 | 0 | 2 | 1 | 1 | 0 | 1 | 0 | 2 | 14 |
| Kachor (2020) | 0 | 0 | 1 | 2 | 0 | 0 | 0 | 0 | 1 | 0 | 1 | 0 | 1 | 6 |
| Osuch (2015/2016) | 2 | 2 | 1 | 2 | 1 | 0 | 0 | 1 | 1 | 1 | 0 | 0 | 1 | 12 |
| Osuch (2019) | 1 | 2 | 1 | 2 | 1 | 0 | 0 | 1 | 2 | 1 | 2 | 1 | 1 | 15 |
| Hides (2010) | 2 | 0 | 1 | 2 | 1 | 0 | 2 | 2 | 2 | 2 | 1 | 1 | 1 | 17 |
| Bond & Power (2020) | 1 | 2 | 2 | 1 | 2 | 1 | 2 | 2 | 1 | 0 | 1 | 2 | 1 | 18 |
| Rice (2020) | 2 | 1 | 2 | 2 | 0 | 1 | 2 | 0 | 2 | 1 | 1 | 1 | 2 | 17 |
| Waters (2024) | 2 | 2 | 2 | 2 | 0 | 1 | 1 | 0 | 2 | 1 | 1 | 0 | 1 | 15 |
| Peters (2018) | 2 | 2 | 1 | 1 | 0 | 0 | 0 | 0 | 1 | 0 | 1 | 1 | 1 | 10 |
| Alvarez-Jimenez (2021) | 2 | 1 | 2 | 1 | 0 | 0 | 0 | 0 | 1 | 1 | 1 | 1 | 2 | 12 |
| Schley (2019) | 0 | 1 | 1 | 2 | 1 | 1 | 1 | 0 | 1 | 0 | 1 | 2 | 2 | 13 |
| Dowell (2021) | 2 | 1 | 1 | 2 | 0 | 0 | 0 | 0 | 1 | 1 | 0 | 0 | 1 | 9 |
| Hopkins (2016) | 1 | 0 | 1 | 1 | 0 | 0 | 0 | 0 | 1 | 0 | 0 | 0 | 0 | 4 |
| O'Reilly (2022) | 2 | 2 | 1 | 1 | 0 | 2 | 1 | 0 | 2 | 2 | 0 | 2 | 2 | 17 |
| Souza (2011) | 2 | 2 | 2 | 2 | 1 | 1 | 0 | 2 | 2 | 2 | 0 | 2 | 2 | 20 |
| Anderson (2019) | 2 | 2 | 1 | 2 | 2 | 2 | 2 | 2 | 2 | 2 | 2 | 2 | 2 | 25 |
| Ng (2023) | 2 | 2 | 2 | 2 | 2 | 0 | 0 | 0 | 1 | 1 | 0 | 0 | 0 | 12 |
| Waters (2022) | 2 | 1 | 1 | 2 | 0 | 0 | 2 | 0 | 2 | 0 | 2 | 1 | 1 | 14 |
| Duncan (2020) | 1 | 1 | 1 | 2 | 2 | 2 | 0 | 0 | 1 | 2 | 1 | 2 | 2 | 17 |
| Boonstra (2024) | 2 | 2 | 1 | 2 | 1 | 1 | 0 | 0 | 1 | 0 | 0 | 1 | 2 | 13 |
| Telford (2024) | 2 | 1 | 0 | 2 | 2 | 1 | 2 | 0 | 2 | 0 | 2 | 2 | 1 | 17 |
| Van Doorn (2023) | 2 | 0 | 1 | 2 | 2 | 2 | 0 | 2 | 2 | 0 | 0 | 1 | 2 | 16 |
| Note. Q1 = Did the trial address a clearly focused issue?^a^; Q2 = Was the cohort or sample recruited in an acceptable way?^a^ ;Q3 = Was the exposure/intervention/service receipt accurately measured to minimise bias?^a^; Q4 = Was the outcome accurately measured to minimise bias?^a^ ; Q5a= Have the authors identified all the important confounding factors?^a^ ; Q5b= Haver they taken account of the confounding factors in the design and/or analysis?^a^ Q6a = Was the follow up of subjects complete enough?^a^ ; Q6b= was the follow up of subjects long enough?^a^ Q7 = What are the results of the study?^b^ Q8 = How precise are the results?^b^ Q9 = Do you believe the results?^b^ ; Q10 = Can the results be applied in your context? (or to the local population?)^c^ ; Q12 = What are the implications of this study for practice?^c^ Note: Q11 (Do the results of this study fit with other available evidence?) Was removed from the appraisal because there is limited evidence in this area | | | | | | | | | | | | | | |
| 2 = Criterion is completely met ; 1 = Criterion is partially met ; 0 = Criterion not applicable, not met, or not mentioned | | | | | | | | | | | | | | |

^a:^ Validity of the study design

^b:^ Quality of the results

^c^ Extent that results will help locally

**Summary of quality of studies**

The quality of selected cohort studies varied substantially. Most studies clearly stated an aim and utilised objective, validated measures or routine outcomes, thereby minimising bias. The most common quality issue involved whether the authors identified important confounding factors and whether the follow-up periods for subjects were sufficient. Only some studies fully met this criterion with follow-ups of over six months (Anderson, 2019; Hides, 2010; Osuch, 2019; Schley, 2008, Van Doorn 2023). Furthermore, most of the included studies partially met the criteria for the applicability of results to the target population of the study. Examples of papers demonstrating very good applicability include Anderson (2019), Corley (2023), Bond & Power (2020), O'Reilly (2022), and Duncan (2020).

**Quality Appraisal of Case Control Studies**

| Paper | Q1 | Q2 | Q3 | Q4 | Q5 | Q6a | Q6b | Q7 | Q8 | Q9 | Q10 | Total Score (out of 22) |
| --- | --- | --- | --- | --- | --- | --- | --- | --- | --- | --- | --- | --- |
| Uchino (2021) | 1 | 0 | 1 | 1 | 1 | 0 | 0 | 0 | 0 | 0 | 0 | 4 |
| Note. Q1 = Did the trial address a clearly focused issue?^a^; Q2 = Did the authors use an appropriate method to answer their question?^a^ ;Q3 = Were the cases recruited in an acceptable way?^a^; Q4 = Were the controls selected in an acceptable way?^a^ ; Q5= Was the exposure accurately measured to minimise bias?^a^ ; Q6a = What confounding factors have the authors accounted for?^a^ ; Q6b= Have the authors taken account of the potential confounding factors in the design and/or in their analysis?^a^ Q7 = What are the results of the study?^b^ Q8 = How precise are the results?^b^ Q9 = Do you believe the results?^b^ ; Q10 = Can the results be applied in your context? (or to the local population?)^c^ Note: Q11 (Do the results of this study fit with other available evidence?) Was removed from the appraisal because there is limited evidence in this area | | | | | | | | | | | | |
| 2 = Criterion is completely met ; 1 = Criterion is partially met ; 0 = Criterion not applicable, not met, or not mentioned | | | | | | | | | | | | |

^a:^ Validity of the study design

^b:^ Quality of the results

^c^ Extent that results will help locally

**Summary of quality of studies**

In the quality appraisal of the included case-control study by Uchino (2021), the paper did not meet the criteria for credibility or effectively demonstrating the impact of the intervention on outcomes and context due to its small sample size and largely descriptive pre-post result scores. Additionally, the study did not address confounding factors. However, it did partially meet the criteria for having a clear aim and for an acceptable method of selection of cases and controls.
