## Appendix 3 for "Approaches to early intervention for common mental health problems in young people: a systematic review"

### Appendix 3: Studies excluded at full text with reasons

| **Screening source** | **Study Author** | **Title** | **Exclusion reason** |
| --- | --- | --- | --- |
| Academic literature database searches | Abas 2016 | 'Opening up the mind': Problem-solving therapy delivered by female lay health workers to improve access to evidence-based care for depression and other common mental disorders through the Friendship Bench Project in Zimbabwe | Study design/publication type |
| Academic literature database searches | Abrahams 2002 | An Evaluation of a Primary Careâ€Based Child Clinical Psychology Service | Population |
| Academic literature database searches | Adrian 2015 | Occupied bed days a redundant currency? An evaluation of the first 10 years of an integrated model of care for mentally ill adolescents | Population |
| Academic literature database searches | Ahmadi 2020 | TRAUMA-INFORMED EVIDENCE-BASED SCREENING PROCEDURE AND EARLY INTERVENTION FOR YOUTH WITH PTSD | Study design/publication type |
| Academic literature database searches | Aitken 2023 | Development, reach, acceptability and associated clinical changes of a group intervention to improve caregiver-adolescent relationships in the context of adolescent depression | Intervention |
| Academic literature database searches | Akiba 2023 | Champion and audit and feedback strategy fidelity and their relationship to depression intervention fidelity: A mixed method study | Intervention |
| Academic literature database searches | Akroyd 2015 | How can an early years setting support the mental health of young children and why is this important? | Study design/publication type |
| Academic literature database searches | Aldridge 2022 | Cost-effectiveness of psychological intervention within services for depression delivered by primary care workers in Nepal: Economic evaluation of a randomized control trial | Population |
| Academic literature database searches | Al-khatib 2015 | A Family Consultation Service: Single session intervention to build the mental health and wellbeing of children and their families | Study design/publication type |
| Academic literature database searches | Allgaier 2014 | [Early detection of child and adolescent depression exemplifying the role of screening tools in mental health care] | Intervention |
| Academic literature database searches | Anderson 2017 | Care Coordination Services: A Description of an Alternative Service Model for At-Risk Families | Population |
| Academic literature database searches | Andersson 2007 | Unmet mental health service needs among Norwegian children and adolescents | Study design/publication type |
| Academic literature database searches | Andres 2004 | A prevention and early detection program for the treatment of eating disorders: Experience from Spain. [Spanish] | Population |
| Academic literature database searches | Andrews 2015 | A pilot investigation of Quest Institute Cognitive Hypnotherapy services using Improving Access to Psychological Therapies as the benchmark | Population |
| Academic literature database searches | Annamalai 2018 | Establishing an integrated health care clinic in a community mental health center: Lessons learned | Study design/publication type |
| Academic literature database searches | Anonymous 1981 | Early intervention program: the continuing saga of a wonderfully worthwhile mental health program | Full text not available |
| Academic literature database searches | Anonymous 2002 | Significant achievement awards: the Theiss Child Development Center--an innovative provider of early childhood mental health services | Study design/publication type |
| Academic literature database searches | Anonymous 2010 | Early intervention in panic: Pragmatic randomised controlled trial (British Journal of Psychiatry 196, (326-331)) | Population |
| Academic literature database searches | Anonymous 2017 | Cognitive therapy effective early treatment for children with PTSD | Study design/publication type |
| Academic literature database searches | Armando 2014 | Early Intervention Services versus Generic Community Mental Health Services: A Paradigm Shift | Study design/publication type |
| Academic literature database searches | Aronen 2000 | Effects of early intervention on psychiatric symptoms of young adults in low-risk and high-risk families | Intervention |
| Academic literature database searches | Arrojo 2023 | Pediatric Primary-Care Integrated Behavioral Health A Framework for Reducing Inequities in Behavioral Health Care and Outcomes for Children | Study design/publication type |
| Academic literature database searches | Arthur 2005 | Allocating cases in a CAMHS team | Study design/publication type |
| Academic literature database searches | Asarnow 2015 | Integrating depression treatment within primary care improves outcomes in adolescents | Study design/publication type |
| Academic literature database searches | Ashby 2016 | Implications of Comprehensive Mental Health Services Embedded in an Adolescent Obstetric Medical Home | Intervention |
| Academic literature database searches | Astrup 1977 | Attempts at applying psychophysiological measures for early detection and treatment of mental illness | Study design/publication type |
| Academic literature database searches | Auclair 2012 | Mental health in Inuit youth from Nunavik: Clinical considerations on a transcultural, interdisciplinary, community-oriented approach | Study design/publication type |
| Academic literature database searches | Aupont 2013 | A Collaborative Care Model to Improve Access to Pediatric Mental Health Services | Population |
| Academic literature database searches | Austin 1999 | Description and evaluation of a domiciliary perinatal mental health service focussing on early intervention | Population |
| Academic literature database searches | Austin 2000 | The Eastern Sydney Perinatal Mental Health Service: A model for early intervention in depression | Study design/publication type |
| Academic literature database searches | Austin 2015 | Psychosocial Assessment and Integrated Perinatal Care | Study design/publication type |
| Academic literature database searches | Austin 2022 | The First Episode Rapid Early Intervention for Eating Disorders - Upscaled study: Clinical outcomes | Population |
| Academic literature database searches | Axford 2021 | The Effectiveness of a Community-Based Mentoring Program for Children Aged 5-11 Years: Results from a Randomized Controlled Trial | Population |
| Academic literature database searches | Ayano 2017 | Mental health training for primary health care workers and implication for success of integration of mental health into primary care: Evaluation of effect on knowledge, attitude and practices (KAP) | Population |
| Academic literature database searches | Ayoub 2014 | Early Head Start: mental health, parenting, and impacts on children | Study design/publication type |
| Academic literature database searches | Bachmann 2009 | Integrating children's services in England: national evaluation of children's trusts | Study design/publication type |
| Academic literature database searches | Bai 2018 | Reducing health risk behaviors and improving depression in adolescents: A randomized controlled trial in primary care clinics | Intervention |
| Academic literature database searches | Baker-Ericzen 2008 | Partnership for Women's Health: a new-age collaborative program for addressing maternal depression in the postpartum period | Population |
| Academic literature database searches | Balakrishna 2015 | Early intervention for increased antenatal anxiety associated with foetal development risk | Population |
| Academic literature database searches | Banasiewicz 2013 | Integrative care for adolescent mood problems: brief report from a Pediatric Second Opinion Clinic | Study design/publication type |
| Academic literature database searches (update search) | Barbic 2024 | Implementing foundry: A cohort study describing the regional and virtual expansion of a youth integrated service in british columbia, canada | Outcomes |
| Academic literature database searches | Barker 2020 | Values-Based Interventions in Patient Engagement for Those with Complex Needs | Population |
| Academic literature database searches | Barkham 2021 | Person-centred experiential therapy versus cognitive behavioural therapy delivered in the English Improving Access to Psychological Therapies service for the treatment of moderate or severe depression (PRaCTICED): A pragmatic, randomised, non-inferiority | Population |
| Academic literature database searches | Barnard 1988 | Prevention of parenting alterations for women with low social support.DP - Aug 1988 | Population |
| Academic literature database searches | Barnet 2007 | Home visiting for adolescent mothers: effects on parenting, maternal life course, and primary care linkage | Population |
| Academic literature database searches | Bassilios 2016 | Improving access to primary mental health care for Australian children | Population |
| Academic literature database searches | Baten 1995 | Occupational therapy in child and adult psychiatry | Full text not available |
| Academic literature database searches | Beatty 2019 | Referral pathways between the mental health services and Jigsaw | Study design/publication type |
| Academic literature database searches (update search) | Bechdolf 2024 | soulspace: Integrated youth mental health care in Berlin, Germany-An introduction to the program and a description of its users | Outcomes |
| Academic literature database searches | Bell 2008 | Should culture considerations influence early intervention? | Study design/publication type |
| Academic literature database searches | Bell 2014 | A multi-agency evaluation of the Leeds Dual Diagnosis care co-ordination protocol | Population |
| Academic literature database searches | Bendelius 2005 | Eating disorders: early detection can lead to improved outcomes | Population |
| Grey literature database searches | Berkowitz 2011 | A 4-session intervention shows promise for preventing PTSD symptoms: Reference to study ‘The Child and Family Traumatic Stress Intervention: Secondary Prevention for Youth at Risk Youth of Developing PTSD’ | Intervention |
| Academic literature database searches | Bernstein 2018 | The world was not built for us: Improving access to care for transgender youth | Study design/publication type |
| Grey literature database searches | Bevington 2013 | Innovations in practice : Adolescent mentalisation-based integrative therapy (AMBIT) - a new integrated approach to working with the most hard to reach adolescents with severe complex mental health needs | Study design/publication type |
| Academic literature database searches | Bhat 2018 | Delivering perinatal depression care in a rural obstetric setting: A mixed methods study of feasibility, acceptability and effectiveness | Population |
| Academic literature database searches | Bhatara 1981 | The comprehensive diagnosis of developmental behavioral disorders in primary care: an integrated approach | Study design/publication type |
| Academic literature database searches | Bhatia 2018 | Anxiety disorders in children and adolescents: Need for early detection | Study design/publication type |
| Academic literature database searches | Bhavsar 2021 | The association of migration and ethnicity with use of the Improving Access to Psychological Treatment (IAPT) programme: A general population cohort study | Outcome |
| Academic literature database searches | Birnbaum 2022 | Digital Strategies to Accelerate Help-Seeking in Youth With Psychiatric Concerns in New York State | Population |
| Academic literature database searches (update search) | Birnbaum 2022 | Digital strategies to accelerate help-seeking in youth with psychiatric concerns in New York State | Outcomes |
| Academic literature database searches | Birrane 2015 | Development and evaluation of an educational intervention in youth mental health for primary care practitioners | Study design/publication type |
| Academic literature database searches | Bittner 2014 | Early Intervention in Pregnant Women With Elevated Anxiety and Depressive Symptoms <i>Efficacy of a Cognitive</i>-<i>Behavioral Group Program</i> | Population |
| Academic literature database searches (update search) | Blais 2024 | Using Neuropsychological Profiling to Tailor Mental Health Care for Children and Youth: a Quality Improvement Project to Measure Feasibility | Outcomes |
| Academic literature database searches | Blakemore 2009 | Early intervention in childhood is the key to avoiding psychiatric illness in later life | Study design/publication type |
| Academic literature database searches | Bogucki 2021 | Cognitive behavioral therapy for depressive disorders: Outcomes from a multi-state, multi-site primary care practice | Population |
| Academic literature database searches | Bondar 2020 | Symptom clusters in adolescent depression and differential response to treatment: A secondary analysis of the Treatment for Adolescents with Depression Study randomised trial | Intervention |
| Academic literature database searches | Boobpamela 2022 | Effectiveness of an early depression prevention program on coping skills and depression among pregnant adolescents: a randomized controlled trial | Intervention |
| Academic literature database searches | Borghini 2014 | Effects of an early intervention on maternal post-traumatic stress symptoms and the quality of mother-infant interaction: The case of preterm birth | Population |
| Academic literature database searches | Bradley 2022 | Adolescent Suicide: Are There Warning Signs? | Study design/publication type |
| Academic literature database searches | Brakoulias 2018 | A call for prevention and early intervention in obsessive-compulsive disorder | Study design/publication type |
| Academic literature database searches | Brakoulias 2021 | Short communication: A report of the first twelve months of an early intervention service for obsessive-compulsive disorder (OCD) | Population |
| Academic literature database searches (update search) | Brakoulias 2021 | Short communication: A report of the first twelve months of an early intervention service for obsessive-compulsive disorder (OCD) | Population |
| Academic literature database searches | Brettschneider 2020 | Cost-effectiveness of guideline-based stepped and collaborative care versus treatment as usual for patients with depression-A cluster-randomized trial | Outcome |
| Academic literature database searches | Bridges 2015 | Diagnoses, Intervention Strategies, and Rates of Functional Improvement in Integrated Behavioral Health Care Patients | Population |
| Academic literature database searches | Broersen 2020 | Young Patients With Persistent and Complex Care Needs Require an Integrated Care Approach: Baseline Findings From the Multicenter Youth Flexible ACT Study | Outcome |
| Academic literature database searches (update search) | Broersen 2022 | Case study in youth flexible assertive community treatment: An illustration of the need for integrated care | Population |
| Academic literature database searches | Broersen 2023 | Effects of youth flexible assertive community treatment: Outcomes of an 18-month observational study.PS - First Posting | Population |
| Academic literature database searches | Broersen 2023 | Effects of Youth Flexible Assertive Community Treatment: outcomes of an 18-month observational study | Population |
| Academic literature database searches (update search) | Broersen 2024 | Effects of Youth Flexible Assertive Community Treatment: outcomes of an 18-month observational study | Population |
| Academic literature database searches | Brown 1999 | New community mental health team for acute psychiatric illness.DP - Mar 1999 | Population |
| Academic literature database searches | Brown 2014 | How equitable are psychological therapy services in South East London now? A comparison of referrals to a new psychological therapy service with participants in a psychiatric morbidity survey in the same London borough | Population |
| Academic literature database searches | Brown 2019 | Availability of integrated primary care services in community mental health care settings | Intervention |
| Academic literature database searches | Browne 2010 | Employment services as an early intervention for young people with mental illness | Intervention |
| Academic literature database searches | Buchanan 2021 | Integration of behavioral health services and adolescent depression screening in primary care | Outcome |
| Academic literature database searches | Buntrock 2017 | Preventing depression in adults with subthreshold depression: Health-economic evaluation alongside a pragmatic randomized controlled trial of a web-based intervention | Population |
| Academic literature database searches | Burman 2008 | Depression and anxiety outcomes at a free clinic in a rural state | Population |
| Academic literature database searches | Burnam 2014 | Evaluation of the California Mental Health Services Authority's Prevention and Early Intervention Initiatives: Progress and Preliminary Findings | Population |
| Academic literature database searches | Buus 2019 | The association between Open Dialogue to young Danes in acute psychiatric crisis and their use of health care and social services: A retrospective register-based cohort study | Outcome |
| Academic literature database searches | Buzi 2014 | Project Passport: An Integrated Group-Centered Approach Targeting Pregnant Teens and Their Partners | Study design/publication type |
| Academic literature database searches | Byrne 2014 | Early Intervention in the General Hospital | Study design/publication type |
| Academic literature database searches | Callaly 2010 | Mental health services for young people - the challenge of integrating services | Study design/publication type |
| Academic literature database searches | Callaly 2014 | Early Intervention for Young People with Mental Illness | Study design/publication type |
| Academic literature database searches | Calvano 2021 | Evaluation of an Early Intervention Model for Child and Adolescent Victims of Interpersonal Violence | Population |
| Academic literature database searches | Calveley 2009 | A patient-centred referral pathway for mild to moderate lifestyle and mental health problems: does this model work in practice? | Population |
| Academic literature database searches | Camilleri 2017 | Innovations in Practice: A case control and follow-up study of 'hard to reach' young people who suffered from multiple complex mental disorders | Population |
| Academic literature database searches | Camm 2005 | Early intervention and mental health | Study design/publication type |
| Academic literature database searches | Canady 2019 | Embedding child psychiatrist in PC practice could remove barriers to care | Study design/publication type |
| Academic literature database searches | Cano-Vindel 2022 | Improving access to psychological therapies in Spain: From IAPT to PsicAP | Intervention |
| Academic literature database searches | Carignan 2013 | [Gaining access in mental health: an experience in six stages] | Study design/publication type |
| Academic literature database searches | Carleton 2020 | Enhancing the scalability of the collaborative care model for depression using mobile technology | Population |
| Academic literature database searches | Carlsson 2010 | Late Mental Health Changes in Tortured Refugees in Multidisciplinary Treatment | Population |
| Academic literature database searches | Carroll 2009 | Integrated cognitive behaviour therapy for co-occurring substance misuse and major depression: lessons from a youth mental health service | Study design/publication type |
| Academic literature database searches (update search) | Carroll 2024 | Racial match: Black youth and pediatric integrated primary care | Population |
| Academic literature database searches | Chakawa 2022 | Bridging the gap: A pilot study of a lay health worker model to decrease child mental health stigma and promote parents' professional help-seeking for Black/African American children | Population |
| Academic literature database searches (update search) | Chakawa 2024 | Disparities in accessing specialty behavioral health services during the COVID-19 pandemic and why we need pediatric integrated primary care | Population |
| Academic literature database searches | Chowdhury 2004 | How to operationalise community mental health service at the primary care?: experience of IRMC model from Sundarban, India | Population |
| Academic literature database searches | Chugg 2009 | Managed Networks and Integrated Children's Services Case Study of Devon | Study design/publication type |
| Academic literature database searches | Clark 2018 | Improving Access to Child and Adolescent Mental Health Care: The Choice and Partnership Approach | Population |
| Academic literature database searches (update search) | Clarke 2023 | Integrating trauma-informed services in out-of-school time programs to mitigate the impact of community gun violence on youth mental health | Population |
| Academic literature database searches | Clemente 2006 | Evaluation of a Waiting List Initiative in a Child and Adolescent Mental Health Service | Population |
| Academic literature database searches | Clementi 2016 | Sleep-Related Outcomes Following Early Intervention for Childhood Anxiety | Population |
| Academic literature database searches | Coggins 2021 | The efficacy of child-teacher relationship training as an early childhood mental health intervention in Head Start programs | Population |
| Academic literature database searches (update search) | Colizzi 2024 | Lessons learnt from running a transition-age youth mental health outpatient clinic in italy: The precocity of intervention in adolescent medicine (prima) experience | Population |
| Academic literature database searches | Constant 1986 | The clinical picture of the psychopathologic evaluation in a multidisciplinary team | Study design/publication type |
| Academic literature database searches | Cooper 1999 | Demographic and social backdrop: How changes in society support the need for a services integration perspective to service delivery in schools and communities | Study design/publication type |
| Academic literature database searches | Cosgrave 2008 | Met and unmet need in youth mental health | Outcome |
| Academic literature database searches | Courtney 2021 | 11.9 The Caribou-1 Care Pathway for Adolescent Depression: A Pilot Controlled Clinical Trial | Study design/publication type |
| Academic literature database searches | Courtney 2022 | CARIBOU‐1: A pilot controlled trial of an Integrated Care Pathway for the treatment of depression in adolescents. | Population |
| Academic literature database searches | Cox 2010 | Wraparound retrospective: factors predicting positive outcomes | Intervention |
| Academic literature database searches | Cromarty 2016 | NewAccess for depression and anxiety: Adapting the UK Improving Access to Psychological Therapies Program across Australia | Study design/publication type |
| Academic literature database searches (update search) | Crosland 2024 | Cost-effectiveness of system-level mental health strategies for young people in the Australian Capital Territory: a dynamic simulation modelling study | Outcomes |
| Academic literature database searches | Cross 2014 | A Clinical Staging Model for Early Intervention Youth Mental Health Services | Study design/publication type |
| Academic literature database searches | Cross 2018 | Variability in Clinical Outcomes for Youths Treated for Subthreshold Severe Mental Disorders at an Early Intervention Service | Population |
| Academic literature database searches | Crowe 1981 | Father involvement in early intervention programs | Study design/publication type |
| Academic literature database searches | Cusack 2008 | Targeting trauma-related interventions and improving outcomes for women with co-occurring disorders | Population |
| Academic literature database searches | Daulay 2022 | Family Empowerment Models in Fighting the Problem of Mental Health Children and Adolescent | Study design/publication type |
| Academic literature database searches | Davey 2019 | Early intervention for depression in young people: a blind spot in mental health care | Study design/publication type |
| Academic literature database searches | David 2023 | Are gains in emotional symptoms and emotion-regulation competencies after the REThink therapeutic game maintained in the long run? A 6-month follow-up | Intervention |
| Academic literature database searches | Davidson 2018 | Designing a person-centred care pathway for people with depression in primary care | Study design/publication type |
| Academic literature database searches | Dawson 2005 | Early intervention in mental health | Study design/publication type |
| Academic literature database searches | Diatkine 1982 | New therapeutic pathways in child psychiatry: Work of an evening unit. [French] | Intervention |
| Academic literature database searches | DiBona 2014 | Predictors of patient non-attendance at Improving Access to Psychological Therapy services demonstration sites | Population |
| Academic literature database searches | Dillon-Naftolin 2017 | Implementing Integrated Care in Pediatric Mental Health: Principles, Current Models, and Future Directions | Study design/publication type |
| Academic literature database searches (update search) | Dingwall 2023 | Feasibility and Acceptability of the Aboriginal and Islander Mental Health Inititative for Youth app: Nonrandomized pilot with First Nations young people | Intervention |
| Academic literature database searches | Dixon 2011 | Linking young homeless people to mental health services: An exploration of an outreach clinic at a supported youth accommodation service | Study design/publication type |
| Academic literature database searches | Doey 2008 | Creating primary care access for mental health care clients in a community mental health setting | Population |
| Academic literature database searches | Dubin 2001 | Insights: health. Depression: more than a mood. Children are more depressed than ever, but early detection helps | Study design/publication type |
| Academic literature database searches | Eapen 2012 | Health and education: Service providers in partnership to improve mental health | Study design/publication type |
| Academic literature database searches | Eapen 2023 | Stemming the tide of mental health problems in young people: Challenges and potential solutions | Study design/publication type |
| Academic literature database searches | Eberhart 2015 | Evaluation of California's Statewide Mental Health Prevention and Early Intervention Programs: Summary of Key Year 2 Findings | Study design/publication type |
| Academic literature database searches | Emerson 2021 | Mental health mobile app use: Considerations for serving underserved patients in integrated primary care settings | Study design/publication type |
| Grey literature database searches | Emslie 2009 | Early Treatment Decisions Crucial for Teens with Treatment-resistant Depression (https://www.nimh.nih.gov/news/science-news/2010/early-treatment-decisions-crucial-for-teens-with-treatment-resistant-depression): Contains link to study: ‘Treatment of Resistant Depression in Adolescents (TORDIA): Week 24 Outcome | Population |
| Academic literature database searches | Eniola 2022 | Improving Access to Adolescent Primary Healthcare Services | Study design/publication type |
| Academic literature database searches | Esbjorn 2019 | Increasing access to low-intensity interventions for childhood anxiety: A pilot study of a guided self-help program for Scandinavian parents | Population |
| Academic literature database searches | Etheridge 2004 | Recognising and responding to adolescents with mental illness | Study design/publication type |
| Academic literature database searches | Etherington 2019 | Comparing universal and targeted delivery of a mindfulness-based program for anxiety in children | Intervention |
| Academic literature database searches | Falloon 1992 | Prevention of major depressive episodes: Early intervention | Study design/publication type |
| Academic literature database searches | Fallucco 2021 | Child Psychiatry Consultation Clinic for Pediatricians: Long-Term Outcomes | Study design/publication type |
| Academic literature database searches | Fazel 2021 | How does reorganisation in child and adolescent mental health services affect access to services? An observational study of two services in England | Study design/publication type |
| Academic literature database searches | Felipe 2023 | Integrative community therapy for the promotion of mental health in adolescents: A quasi-experimental study | Intervention |
| Academic literature database searches | Field 2000 | Targeting adolescent mothers with depressive symptoms for early intervention | Intervention |
| Academic literature database searches | Figueroa 2022 | The ABCDE psychological first aid intervention decreases early PTSD symptoms but does not prevent it: Results of a randomized-controlled trial | Population |
| Academic literature database searches | Firth 2015 | Therapist effects and moderators of effectiveness and efficiency in psychological wellbeing practitioners: A multilevel modelling analysis | Population |
| Academic literature database searches | Flygare 2022 | Implementing therapist-guided internet-delivered cognitive behaviour therapy for obsessive-compulsive disorder in theUK's IAPT programme: A pilot trial | Population |
| Academic literature database searches | Folayan 2020 | A proposed one-stop-shop approach for the delivery of integrated oral, mental, sexual and reproductive healthcare to adolescents in Nigeria | Study design/publication type |
| Academic literature database searches | Fortuna 2018 | A treatment development study of a cognitive and mindfulness-based therapy for adolescents with co-occurring post-traumatic stress and substance use disorder | Intervention |
| Academic literature database searches | Fountaine 2023 | Examining the Utility of a Telehealth Warm Handoff in Integrated Primary Care for Improving Patient Engagement in Mental Health Treatment: Randomized Video Vignette Study | Intervention |
| Academic literature database searches | Fox 2020 | Evaluating a low‐intensity cognitive behavioural program for young people in regional Australia | Intervention |
| Academic literature database searches | Fuchs 2016 | Implementation of an acceptance- and mindfulness-based group for depression and anxiety in primary care: Initial outcomes | Population |
| Academic literature database searches | Fuggle 2023 | Outcomes for Adaptive Mentalization Based Integrative Treatment informed care for adolescents using a deployment-based approach | Study design/publication type |
| Academic literature database searches | Gale 2003 | The primary mental health worker within child and adolescent mental health services | Study design/publication type |
| Academic literature database searches | Gallegos 2012 | The FRIENDS for Life program for Mexican girls living in an orphanage: A pilot study | Population |
| Academic literature database searches (update search) | Ganga 2024 | The Impact of a Digital Referral Platform to Improve Access to Child and Adolescent Mental Health Services: A Prospective Observational Study with Real-World Data | Population |
| Academic literature database searches | Gardiner 2022 | Nature-based physical activity as an early intervention for teenagers | Study design/publication type |
| Academic literature database searches | Gardner 2003 | Integrated care pathway for children who attempt deliberate self harm | Study design/publication type |
| Academic literature database searches | Garralda 2016 | What is special about a Paediatric Liaison Child and Adolescent Mental Health service? | Population |
| Academic literature database searches | Gaylord 2015 | Introducing shared mental health care in Northwestern Ontario: An analysis of changing referral patterns of primary care providers | Population |
| Academic literature database searches | Gehue 2021 | Piloting the 'Youth Early-intervention Study ' ('YES'): Preliminary functional outcomes of a randomized controlled trial targeting social participation and physical well-being in young people with emerging mental disorders | Population |
| Academic literature database searches | Geist 2004 | Improving access to mental health services for youth and parents | Study design/publication type |
| Academic literature database searches | Gellatly 2018 | @Home eTherapy service for people with common mental health problems: An evaluation | Population |
| Academic literature database searches | Gershkovich 2021 | Integrating exposure and response prevention with a mobile app to treat obsessive-compulsive disorder: Feasibility, acceptability, and preliminary effects | Population |
| Academic literature database searches | Gibbons 2019 | Predictors of treatment attendance in cognitive and dynamic therapies for major depressive disorder delivered in a community mental health setting | Population |
| Academic literature database searches | Gillis 1984 | Early detection of the suicidal adolescent | Study design/publication type |
| Academic literature database searches | Ginsburg 2016 | An Open Trial of the Anxiety Action Plan ( AxAP): A Brief Pediatrician-Delivered Intervention for Anxious Youth | Population |
| Academic literature database searches | Gobbart 2013 | â€˜Changing Habitsâ€™: an evaluation of a dual diagnosis focused, integrated, multimodal, psychosocial education and skill building group programme delivered in a community-based setting | Population |
| Academic literature database searches | Godoy 2017 | Behavioral Health Integration in Health Care Settings: Lessons Learned from a Pediatric Hospital Primary Care System | Study design/publication type |
| Academic literature database searches | Godoy 2023 | DC Mental Health Access in Pediatrics: Evaluating a Child Psychiatry Access Program in Washington, DC | Study design/publication type |
| Academic literature database searches (update search) | Godoy 2024 | Linking primary care to community-based mental health resources via family navigation and phone-based care coordination | Population |
| Academic literature database searches | Goldston 2021 | Feasibility of an integrated treatment approach for youth with depression, suicide attempts, and substance use problems | Intervention |
| Academic literature database searches (update search) | Gomez 2024 | Expanding the Behavioral Health Workforce: Pediatric Psychologists Training Master's Level Students in Integrated Healthcare | Outcomes |
| Academic literature database searches | Goossen 2008 | Does the introduction of shared care therapists in primary health care impact clients' mental health symptoms and functioning? | Population |
| Academic literature database searches | Gozlan 2009 | [Toward integrated care in mental health: experience in the area of Prepsy] | Study design/publication type |
| Academic literature database searches | Griffiths 2017 | Innovations in practice: Evaluating clinical outcome and service utilization in an AMBIT-trained Tier 4 child and adolescent mental health service | Population |
| Academic literature database searches | Grimes 2018 | Enhanced Child Psychiatry Access and Engagement via Integrated Care: A Collaborative Practice Model With Pediatrics | Population |
| Academic literature database searches | Guerrero 2003 | Early detection and intervention for common causes of psychosocial morbidity and mortality in children and adolescents | Study design/publication type |
| Academic literature database searches | Guess 2006 | Posttraumatic Stress Disorder Early Detection is Key | Study design/publication type |
| Academic literature database searches | Gwynne 2009 | Pilot evaluation of an early intervention programme for children at risk | Population |
| Academic literature database searches | Hadjistavropoulos 2014 | Improving access to psychological services through therapist-assisted, Internet-delivered cognitive behaviour therapy | Study design/publication type |
| Academic literature database searches | Hagell 2016 | Developing an integrated primary health care and youth work service for young people in Lambeth: learning from the Well Centre | Study design/publication type |
| Academic literature database searches | Halsall 2014 | mind your mind : An Overview and Evaluation of a Web-Facilitated Mental Health Program that Applies Empowerment Strategies for Youth | Population |
| Academic literature database searches | Hannan 2000 | The prevention of depression in children: A pilot study | Population |
| Academic literature database searches | Harmon 2021 | Addressing the Long-term Effects of Maternal Depression Through Early Intervention | Study design/publication type |
| Academic literature database searches (update search) | Harris-Lane 2024 | Improving access to child and youth addiction and mental health services in new brunswick: Implementing one-at-a-time therapy within an integrated service delivery model | Outcomes |
| Academic literature database searches (update search) | Harrsen 2024 | Effects of an Integrated Treatment Program on Grief and Distress Among Parentally Bereaved Young Adults | Intervention |
| Academic literature database searches | Harwood 2023 | Variations by ethnicity in referral and treatment pathways for IAPT service users in South London | Intervention |
| Academic literature database searches | Henderson 2020 | From planning to implementation of the YouthCan IMPACT Project: A formative evaluation | Study design/publication type |
| Academic literature database searches | Henderson 2023 | Youth Wellness Hubs Ontario: An Innovative Model for Engagement and Equity-Based Considerations in Funding Allocations in Youth Mental Health | Study design/publication type |
| Academic literature database searches (update search) | Henderson 2023 | Youth wellness hubs Ontario: An innovative model for engagement and equity-based considerations in funding allocations in youth mental health | Outcomes |
| Academic literature database searches | Herman 2019 | The Missouri Prevention Center: A Multidisciplinary Approach to Reducing the Societal Prevalence and Burden of Youth Mental Health Problems | Study design/publication type |
| Academic literature database searches | Hester 1977 | Evaluation and accountability in a parent-implemented early intervention service | Study design/publication type |
| Academic literature database searches | Hodges 2007 | headspace: National Youth Mental Health Foundation: Making headway with rural young people and their mental health | Study design/publication type |
| Academic literature database searches (update search) | Hostutler 2023 | Increasing Access to and Utilization of Behavioral Health Care Through Integrated Primary Care | Population |
| Academic literature database searches | Hoter-Ishay 2022 | Young help-seeker profiles in Israel: The case of the first Israeli headspace centre | Outcome |
| Academic literature database searches | Hui 2022 | LevelMind@JC: Development and evaluation of a community early intervention program for young people in Hong Kong | Study design/publication type |
| Academic literature database searches | Illback 2011 | Transforming youth mental health services and supports in Ireland | Study design/publication type |
| Academic literature database searches | Ion 2017 | Understanding integrated mental health care in "real-world" primary care settings: What matters to health care providers and clients for evaluation and improvement? | Study design/publication type |
| Academic literature database searches | Iorfino 2018 | Delineating the trajectories of social and occupational functioning of young people attending early intervention mental health services in Australia: a longitudinal study | Population |
| Academic literature database searches | Iorfino 2019 | Clinical Stage Transitions in Persons Aged 12 to 25 Years Presenting to Early Intervention Mental Health Services With Anxiety, Mood, and Psychotic Disorders | Population |
| Academic literature database searches | Iorfino 2022 | Social and occupational outcomes for young people who attend early intervention mental health services: a longitudinal study | Population |
| Academic literature database searches (update search) | Iqbal 2024 | Arts-based application of the awareness, agency and motivation framework for community education and mobilisation for early detection and care of children with suicidal ideation in Gilgit-Baltistan | Outcomes |
| Academic literature database searches | Iyer 2019 | A minimum evaluation protocol and stepped-wedge cluster randomized trial of ACCESS Open Minds, a large Canadian youth mental health services transformation project | Study design/publication type |
| Academic literature database searches (update search) | Jackson-Morris 2024 | An investment case analysis for the prevention and treatment of adolescent mental disorders and suicide in England | Outcomes |
| Academic literature database searches | Jacob 2021 | Online counselling and goal achievement: Exploring meaningful change and the types of goals progressed by young people | Intervention |
| Academic literature database searches | Jairam 2014 | Early Intervention in Childhood Disorders | Study design/publication type |
| Academic literature database searches | Jerrell 1999 | Skill, symptom, and satisfaction changes in three service models for people with psychiatric disability | Population |
| Academic literature database searches | Jerrell 2000 | Issues and outcomes in integrated treatment programs for dual disorders.DP - Aug 2000 | Population |
| Academic literature database searches | Jerrott 2022 | Feasibility of Text Messages for Enhancing Therapeutic Engagement Among Youth and Caregivers Initiating Outpatient Mental Health Treatment: Mixed Methods Study | Intervention |
| Academic literature database searches | Jessop 2012 | A consultation service for adult mental health service clients who are parents and their families | Study design/publication type |
| Academic literature database searches | Jones 2010 | Impact of the fast track prevention program on health services use by conduct-problem youth | Population |
| Academic literature database searches | Jones 2018 | Using an Integrated Pediatric Primary Care Program to Serve Low-Income, Urban Families | Study design/publication type |
| Academic literature database searches | Jonker 2020 | Patient referral from primary care to psychological therapy services: A cohort study | Population |
| Academic literature database searches | Jorgensen 2023 | Tracking of depressed mood from adolescence into adulthood and the role of peer and parental support: A partial test of the Adolescent Pathway Model | Population |
| Academic literature database searches (update search) | Joseph 2024 | Child mental health treatment access and retention in integrated primary care and traditional outpatient services | Population |
| Academic literature database searches | Judd 2001 | Improving access for rural Australians to treatment for anxiety and depression: The University of Melbourne Depression and Anxiety Research and Treatment Group-Bendigo Health Care Group initiative | Study design/publication type |
| Academic literature database searches | Jurewicz 2015 | Mental health in young adults and adolescents -supporting general physicians to provide holistic care | Study design/publication type |
| Academic literature database searches | Karkou 2022 | Bringing creative psychotherapies to primary NHS Mental Health Services in the UK: A feasibility study on patient and staff experiences of arts for the blues workshops delivered at Improving Access to Psychological Therapies (IAPT) services | Population |
| Academic literature database searches | Katherine 2012 | Collaborative Care for the Treatment of Depression in Primary Care With a Low-Income, Spanish-Speaking Population: Outcomes From a Community-Based Program Evaluation | Population |
| Academic literature database searches | Kearny 2014 | Children with anxiety disorders: Use of a cognitive behavioral therapy model within a social milieu | Intervention |
| Academic literature database searches | Kennedy 2009 | A selective intervention program for inhibited preschool-aged children of parents with an anxiety disorder: Effects on current anxiety disorders and temperament | Population |
| Academic literature database searches | Kenner 2021 | Individualized Family-Centered Developmental Care: A Model for High-Quality Care | Study design/publication type |
| Academic literature database searches | Kerner 2021 | Trends in the utilization of a peer-supported youth hotline | Intervention |
| Academic literature database searches | Khan 2022 | Mental health considerations of a humanitarian crisis: Identification of needs and delivery of services to Afghan child and adolescent refugees in Qatar | Study design/publication type |
| Academic literature database searches | Kiltackey 2008 | The challenge of integrating employment services with public mental health services in Australia: Progress at the first demonstration site | Study design/publication type |
| Academic literature database searches | Kim 2021 | Mindlink: A stigma-free youth-friendly community-based early-intervention centre in Korea | Study design/publication type |
| Academic literature database searches | Kim 2023 | Association of Integrating Mental Health Into Pediatric Primary Care at Federally Qualified Health Centers With Utilization and Follow-up Care | Population |
| Academic literature database searches | King 2020 | Use of common and unique techniques in the early treatment phase for cognitive-behavioral, interpersonal/emotional, and supportive listening interventions for generalized anxiety disorder | Population |
| Academic literature database searches | Klodnick 2021 | Meeting the developmental needs of young adults diagnosed with serious mental health challenges: The emerge model | Population |
| Academic literature database searches (update search) | Klymkiw 2024 | What do justice-involved youth want from integrated youth services? A conjoint analysis | Outcomes |
| Academic literature database searches | Knapstad 2020 | Effectiveness of Prompt Mental Health Care, the Norwegian version of improving access to psychological therapies: A randomized controlled trial | Population |
| Academic literature database searches | Knapstad 2020 | Prompt Mental Health Care (PMHC): work participation and functional status at 12 months post-treatment | Population |
| Academic literature database searches | Knutson 2018 | Care Coordination for Youth With Mental Health Disorders in Primary Care | Study design/publication type |
| Academic literature database searches (update search) | Koet 2024 | Evaluation of practice nurses' management of paediatric psychosocial problems in general practice | Intervention |
| Academic literature database searches | Kolaas 2023 | Feasibility of a video-delivered mental health course for primary care patients: a single-group prospective cohort study | Population |
| Academic literature database searches | Kolko 2010 | Improving Access to Care and Clinical Outcome for Pediatric Behavioral Problems: A Randomized Trial of a Nurse-Administered Intervention in Primary Care | Population |
| Academic literature database searches | Koper 2024 | Effectiveness of a multidisciplinary treatment with youth-initiated mentoring for youths with mental health needs from multi-problem families: a quasi-experimental study | Population |
| Academic literature database searches | Kosky 1992 | Mental health: Is early intervention the key? | Study design/publication type |
| Academic literature database searches (update search) | Kowalczyk 2024 | Can social prescribing aid childrenâ€™s mental health?: By using social prescribing nurses can offer young people treatments that meet their social and emotional needs | Outcomes |
| Academic literature database searches | Kowitt 2016 | A Pilot Evaluation of an Art Therapy Program for Refugee Youth From Burma | Intervention |
| Academic literature database searches | Kristjansdottir 2016 | Transdiagnostic cognitive behavioural treatment and the impact of co-morbidity: An open trial in a cohort of primary care patients | Population |
| Academic literature database searches | Kuehn 2018 | To Prevent Suicides and Promote Physician Well-Being, Institutions Turn to Early Intervention | Study design/publication type |
| Academic literature database searches | Kuhn 2011 | Improving access to psychological therapies: Systemic therapy in the Newham pilot site | Population |
| Academic literature database searches | Kutash 2014 | Quality Indicators for Multidisciplinary Team Functioning in Community-Based Children's Mental Health Services | Study design/publication type |
| Academic literature database searches | Kutcher 2009 | Child and youth mental health: Integrated health care using contemporary competency-based teams | Study design/publication type |
| Academic literature database searches | Kutcher 2017 | Clinic outcomes of the Pathway Through Care Model: A cross-sectional survey of adolescent depression in Malawi | Intervention |
| Academic literature database searches | Landis 2013 | Effects of different models of integrated collaborative care in a family medicine residency program | Population |
| Academic literature database searches | Leahy 2018 | Role of the general practitioner in providing early intervention for youth mental health: a mixed methods investigation | Population |
| Academic literature database searches | Lederman 2020 | Stepping up early treatment for help-seeking youth with at-risk mental states: Feasibility and acceptability of a real-world exercise program | Population |
| Academic literature database searches | Lee 2013 | Broadening the early intervention paradigm: a one stop shop for youth | Study design/publication type |
| Academic literature database searches | Lee 2016 | Understanding how postnatal depression screening and early intervention works in the real world - A Singaporean perspective | Population |
| Academic literature database searches | Lee 2016 | Understanding How Postnatal Depression Screening and Early Intervention Work in the Real World - A Singaporean Perspective | Population |
| Academic literature database searches | Leech 2020 | eMental health service use among Australian youth: a cross-sectional survey framed by Andersen's model | Intervention |
| Academic literature database searches | Leggatt 2016 | Family peer support work in an early intervention youth mental health service | Study design/publication type |
| Academic literature database searches | Leijdesdorff 2020 | No boundaries: A 2 year experience in a specialized youth mental health care program in the Netherlands | Population |
| Academic literature database searches | Lidbetter 2022 | A reflection on the development and delivery of a community peer support service for clients experiencing anxiety and depression | Intervention |
| Academic literature database searches | Lieberman 2006 | On-site mental health care: a route to improving access to mental health services in an inner-city, adolescent medicine clinic | Intervention |
| Academic literature database searches | Liljeholm 2020 | An integrated mental health and vocational intervention: A longitudinal study on mental health changes among young adults | Intervention |
| Academic literature database searches | Linstead 2022 | Evaluation of the impact and acceptability of Cognitive Behavioural Analysis System of Psychotherapy (CBASP) for chronic depression | Population |
| Academic literature database searches | Lovell 2003 | Improving access to primary mental health care: uncontrolled evaluation of a pilot self-help clinic | Study design/publication type |
| Academic literature database searches | Lynch 2006 | Tackling a long waiting list in a child and adolescent mental health service | Population |
| Academic literature database searches | Lynch 2015 | Feasibility of shelter-based mental health screening for homeless children | Population |
| Academic literature database searches | Lynch 2021 | Cost-effectiveness of Brief Behavioral Therapy for Pediatric Anxiety and Depression in Primary Care | Study design/publication type |
| Academic literature database searches | Malla 2019 | Canadian response to need for transformation of youth mental health services: ACCESS Open Minds (Esprits ouverts) | Study design/publication type |
| Academic literature database searches | Mansell 2021 | A new consultation, Assessment and reflection model (CARM) used in child and adolescence mental health services (CAMHS) | Intervention |
| Academic literature database searches (update search) | Marchini 2024 | From adolescence to adulthood: Understanding care trajectories in an early detection and intervention centre in france | Outcomes |
| Academic literature database searches | Margolis 2018 | A Multidisciplinary, Team-Based Teleconsultation Approach to Enhance Child Mental Health Services in Rural Pediatrics | Population |
| Academic literature database searches | MartÃ­nez 2021 | Feasibility and Acceptability of "Cuida tu animo" (Take Care of Your Mood): An Internet-Based Program for Prevention and Early Intervention of Adolescent Depression in Chile and Colombia | Intervention |
| Academic literature database searches | Martel 2020 | Reaching out to reduce health inequities for MÄori youth | Study design/publication type |
| Academic literature database searches | Martinez 2023 | Optimizing ATTAIN implementation in a federally qualified health center guided by the FRAME-IS | Population |
| Academic literature database searches | Mathias 2022 | Foundry: Early learnings from the implementation of an integrated youth service network | Study design/publication type |
| Academic literature database searches | McCann 2006 | Individual, family, and group therapy for adolescents | Study design/publication type |
| Academic literature database searches | McClatchey 2009 | Efficacy of a camp-based intervention for childhood traumatic grief | Population |
| Academic literature database searches | McCulloch 2014 | Early Intervention in Mental Health Problems: The Role of the Voluntary Sector | Study design/publication type |
| Academic literature database searches | McDevitt-Petrovic 2018 | Preliminary findings of a new primary and community care psychological service in Northern Ireland: Low-intensity cognitive behavioural therapy for common mental health difficulties | Population |
| Academic literature database searches | McGorry 2007 | headspace: Australia's National Youth Mental Health Foundation--where young minds come first | Study design/publication type |
| Academic literature database searches | McGorry 2014 | Early intervention, youth mental health: The value of translational research for reform and investment in mental health | Study design/publication type |
| Academic literature database searches | McGowan 2019 | Pathways to mental health improvement in a community-led area-based empowerment initiative: evidence from the Big Local 'Communities in Control' study, England | Population |
| Grey literature database searches | McShane 2015 | Future in mind: promoting, protecting and improving our children and young people’s mental health and wellbeing | Study design/publication type |
| Academic literature database searches | Mei 2020 | Transdiagnostic early intervention, prevention, and prediction in psychiatry | Study design/publication type |
| Academic literature database searches | Melnyk 2006 | KySS and tell: national nursing initiative helps prevent mental health disorders in children and teens | Study design/publication type |
| Academic literature database searches (update search) | Melnyk 2024 | The evidence-based COPE program: Reducing the time between diagnosing and treating depression and anxiety in youth | Outcomes |
| Academic literature database searches | Meulenbeek 2008 | Early intervention in panic: randomized controlled trial and cost-effectiveness analysis | Study design/publication type |
| Academic literature database searches | Mieloo 2022 | Changes in youth care use after the implementation of community-based support teams: repeated measurement study using registry data and data on team characteristics | Population |
| Academic literature database searches | Milgram 2022 | Youth Top Problems and Early Treatment Response to the Unified Protocols for Transdiagnostic Treatment of Emotional Disorders in Children and Adolescents | Intervention |
| Academic literature database searches | Miller 2014 | Care Coordination Impacts on Access to Care for Children with Special Health Care Needs Enrolled in Medicaid and CHIP | Population |
| Academic literature database searches | Molnar 2018 | Enhancing Early Childhood Mental Health Primary Care Services: Evaluation of MA Project LAUNCH | Population |
| Academic literature database searches | Monson 2011 | Consumer participation in a youth mental health service | Study design/publication type |
| Academic literature database searches | Monson 2021 | How can mental health practitioners collaborate with child welfare practitioners to improve mental health for young people in out of home care? | Study design/publication type |
| Academic literature database searches | Morley 2016 | Is specialized integrated treatment for comorbid anxiety, depression and alcohol dependence better than treatment as usual in a public hospital setting? | Population |
| Academic literature database searches | Mueller-Stierlin 2017 | Does one size really fit all? The effectiveness of a non-diagnosis-specific integrated mental health care program in Germany in a prospective, parallel-group controlled multi-centre trial | Population |
| Academic literature database searches | Mukuria 2013 | Cost-effectiveness of an Improving Access to Psychological Therapies service | Population |
| Academic literature database searches | Muris 2001 | Effects of an early intervention group program for anxious and depressed adolescents: A pilot study | Intervention |
| Academic literature database searches | Muther 2018 | Screening and Assessment for Posttraumatic Stress Disorder in Pediatric Populations in Integrated Primary Care | Study design/publication type |
| Academic literature database searches | Muzik 2012 | Psychiatric illness during pregnancy: Early detection, individualized care can promote health for mother and infant | Study design/publication type |
| Academic literature database searches (update search) | Najm 2023 | Establishing a child and adolescent mental health center in Herat, Afghanistan: a project description | Population |
| Academic literature database searches | Nardi 2007 | Depression in school-aged children - Assessment & early intervention | Study design/publication type |
| Academic literature database searches | Nayak 2022 | Predictors of service utilization of young children and families enrolled in a pediatric primary care mental health promotion and prevention program.PS - First Posting | Population |
| Academic literature database searches | Nazareth 2014 | Prevention and Early Intervention in Depression and Anxiety Disorders | Study design/publication type |
| Academic literature database searches | Nelson 2008 | Description of exemplar cases in the Intensive Mental Health Program: illustrations of application of the therapeutic model | Study design/publication type |
| Academic literature database searches | Neufeld 2012 | Walk-in telemedicine clinics improve access and efficiency: A program evaluation from the perspective of a rural community mental health center | Population |
| Academic literature database searches | Neufeld 2013 | Walk-in telemental health clinics improve access and efficiency: A 2-year follow-up analysis | Population |
| Academic literature database searches | Newman 2022 | Access and attitudinal barriers to engagement in integrated primary care mental health treatment for rural populations | Population |
| Academic literature database searches | Ngo 2020 | Step-up, step-down mental health care service: Evidence from Western Australia's first - A mixed-method cohort study | Population |
| Academic literature database searches | Nikapota 1983 | CONTRIBUTION OF INTEGRATED MENTAL-HEALTH-SERVICES TO CHILD MENTAL-HEALTH | Study design/publication type |
| Academic literature database searches (update search) | Nisarga 2024 | Enhancing child mental health: a stepped care approach by mental health professionals at an early intervention and rehabilitation centre for children, in India | Population |
| Academic literature database searches | Noauthorship 2001 | A unique approach to mental health care for young children-The Multnomah County, Oregon, Early Childhood Mental Health Program.DP - Oct 2001 | Study design/publication type |
| Academic literature database searches | Nordentoft 2017 | Early intervention services are effective and must be defended | Study design/publication type |
| Academic literature database searches | Nuri 2018 | Pathways to care of patients with mental health problems in Bangladesh | Outcome |
| Academic literature database searches | Nuthall 2007 | CBT-based early intervention to prevent panic disorder: A pilot study | Population |
| Grey literature database searches | O'Donnell 2012 | Telephone Administered Cognitive Behavioural Therapy as an Early Intervention for Post-Traumatic Anxiety and Depressive Disorders | Population |
| Academic literature database searches | Olivares 2005 | Early detection and treatment of adolescents with generalized social phobia | Study design/publication type |
| Academic literature database searches | Omylinska-Thurston 2021 | Arts for the Blues: The development of a new evidence-based creative group psychotherapy for depression | Study design/publication type |
| Academic literature database searches | Oostermeijer 2021 | Implementing child and youth mental health services: early lessons from the Australian Primary Health Network Lead Site Project | Study design/publication type |
| Academic literature database searches | Osilla 2022 | Collaboration Leading to Addiction Treatment and Recovery from Other Stresses (CLARO): Process of adapting collaborative care for co-occurring opioid use and mental disorders | Population |
| Academic literature database searches (update search) | Ow 2023 | Patterns of service utilization among youth with substance use service need: A cohort study | Outcomes |
| Academic literature database searches | Pan 2016 | The impact of caregivers on the effectiveness of an early community mental health detection and intervention programme in Hong Kong | Population |
| Academic literature database searches | Parikh 2021 | Integrated Care is Associated With Increased Behavioral Health Access and Utilization for Youth in Crisis | Intervention |
| Academic literature database searches | Parish 2011 | New mental health strategy focuses on early intervention and prevention | Study design/publication type |
| Academic literature database searches | Parker 2016 | The effectiveness of simple psychological and physical activity interventions for high prevalence mental health problems in young people: A factorial randomised controlled trial | Intervention |
| Academic literature database searches | Patel 2021 | Improving access to cognitive behavioural therapy groups for postnatal women following partnership work: a service evaluation | Population |
| Academic literature database searches | Paterson 2001 | Getting in early: A framework for early intervention and prevention in mental health for young people in New South Wales | Study design/publication type |
| Academic literature database searches | Pedrini 2015 | Reasons and pathways of first-time consultations at child and adolescent mental health services in Italy: An observational study | Population |
| Academic literature database searches | Peiper 2017 | Latent class analysis of need descriptors within an Irish youth mental health early intervention program toward a typology of need | Study design/publication type |
| Academic literature database searches | Perez 2020 | The Catalonia Suicide Risk Code: A secondary prevention program for individuals at risk of suicide | Population |
| Academic literature database searches | Picardi 2016 | A randomised controlled trial of the effectiveness of a program for early detection and treatment of depression in primary care | Population |
| Academic literature database searches | Pile 2021 | Harnessing Mental Imagery and Enhancing Memory Specificity: Developing a Brief Early Intervention for Depressive Symptoms in Adolescence | Intervention |
| Academic literature database searches | Pinhas 2014 | Early Intervention in Eating Disorders | Population |
| Academic literature database searches | Pirkis 2004 | Improving access to evidence-based mental health care: General practitioners and allied health professionals collaborate | Population |
| Academic literature database searches | Plamondon 2022 | Integrated group treatment for anxiety disorders: A transdiagnostic effectiveness and feasibility study in a mental health hospital setting | Population |
| Academic literature database searches | Poletti 2021 | Early intervention in psychiatry through a developmental perspective | Study design/publication type |
| Academic literature database searches | Poletti 2022 | Mind the (transition) gap: Youth mental health-oriented early intervention services to overcome the child-adolescent vs. adult hiatus | Study design/publication type |
| Academic literature database searches (update search) | Polihronis 2023 | Closing the referral loop: Piloting a clinical pathway between primary care and community-based mental health and addictions services | Population |
| Academic literature database searches | Pollice 2007 | SMILE: a service for prevention, monitoring and early intervention in psychiatry | Study design/publication type |
| Academic literature database searches | Pollice 2007 | The service for Monitoring and Early Intervention against psychoLogical and mEntal suffering in young people (SMILE) at the University of L'Aquila: first year experience | Population |
| Academic literature database searches | Ponizovsky 2007 | Treatment lag on the way to the mental health clinic among Arab- and Jewish-Israeli patients | Population |
| Academic literature database searches | Poon 2016 | Making in-roads across the youth mental health landscape in Singapore: The Community Health Assessment Team (CHAT) | Population |
| Academic literature database searches | Price 2000 | The treatment of anxiety disorders in a primary care HMO setting.DP - Spr, 2000 | Population |
| Academic literature database searches | Purcell 2011 | Toward a Twenty-First Century Approach to Youth Mental Health Care | Study design/publication type |
| Academic literature database searches | Rassenhofer 2016 | Effectiveness of early intervention in trauma outpatient units. Results of a model project on evaluation of outpatient units according to the Crime Victims Compensation Act | Population |
| Academic literature database searches | Ratnaike 2002 | Reorientation of services toward early intervention in mental health: Brief report.DP - Jun 2002 | Study design/publication type |
| Academic literature database searches | Reininghaus 2023 | Effects of a Novel, Transdiagnostic Ecological Momentary Intervention for Prevention, and Early Intervention of Severe Mental Disorder in Youth (EMIcompass): Findings From an Exploratory Randomized Controlled Trial | Population |
| Academic literature database searches | Rice 2020 | Leveraging the social network for treatment of social anxiety: Pilot study of a youth-specific digital intervention with a focus on engagement of young men | Intervention |
| Academic literature database searches | Richards 2009 | Improving access to psychological therapies: Phase IV prospective cohort study | Population |
| Academic literature database searches | Richards 2011 | Implementation of psychological therapies for anxiety and depression in routine practice: Two year prospective cohort study | Population |
| Academic literature database searches | Rickwood 2019 | Australia's innovation in youth mental health care: The headspace centre model | Study design/publication type |
| Academic literature database searches | Riggs 2002 | A clinical approach to integrating treatment for adolescent depression and substance abuse | Study design/publication type |
| Academic literature database searches | Roberts 2019 | Mind and Body: an early intervention group programme for adolescents with self-harm thoughts and behaviours | Intervention |
| Academic literature database searches | Robinson 2020 | The impact of primary care behavioral health services on patient behaviors: A randomized controlled trial | Population |
| Academic literature database searches | Rocks 2020 | Introducing a single point of access (SPA) to child and adolescent mental health services in England: a mixed-methods observational study | Population |
| Academic literature database searches | Rohde 2014 | Sequenced Versus Coordinated Treatment for Adolescents With Comorbid Depressive and Substance Use Disorders | Population |
| Academic literature database searches | Rosello 2021 | Early intervention in child and adolescent eating disorders: The role of a parenting group | Population |
| Academic literature database searches | Rowe 2020 | Co-designing the InnoWell Platform to deliver the right mental health care first time to regional youth | Study design/publication type |
| Academic literature database searches | Ruglass 2017 | Concurrent treatment with prolonged exposure for co-occurring full or subthreshold posttraumatic stress disorder and substance use disorders: A randomized clinical trial | Population |
| Academic literature database searches | Russell 2021 | Homelessness youth and mental health service utilization: A long-term follow-up study | Outcome |
| Academic literature database searches | Russell 2022 | Improving access for the vulnerable: a mixed-methods feasibility study of a pop-up model of care in south-eastern Melbourne, Australia | Population |
| Academic literature database searches | Rust-Overman 2024 | Enhancing Pediatric Mental Health Care in an Outpatient Primary Care Setting Using the Keep Your Children/Yourself Safe and Secure (KySS) Program: A Multidisciplinary Quality Improvement Project | Study design/publication type |
| Academic literature database searches | Ryall 2008 | Intensive youth outreach in mental health: an integrated framework for understanding and intervention | Study design/publication type |
| Academic literature database searches | SÃ¡nchez-GarcÃ­a 2009 | Effectiveness of a program for early detection/intervention in children/adolescents with generalized social phobia | Population |
| Academic literature database searches | Sadock 2017 | Initial and follow-up evaluations of integrated psychological services for anxiety and depression in a safety net primary care clinic | Population |
| Academic literature database searches | Saether 2019 | Twelve months post-treatment results from the Norwegian version of improving access to psychological therapies | Population |
| Academic literature database searches | Salisbury 2016 | Effectiveness of an integrated telehealth service for patients with depression: A pragmatic randomised controlled trial of a complex intervention | Population |
| Academic literature database searches | Samokhvalov 2018 | Outcomes of an integrated care pathway for concurrent major depressive and alcohol use disorders: A multisite prospective cohort study | Population |
| Academic literature database searches | SanPio 2023 | A randomized controlled implementation trial of a multicomponent integrated care program to empower mental health service users and their relatives throughout the recovery process | Population |
| Academic literature database searches | Sarvet 2010 | Improving Access to Mental Health Care for Children: The Massachusetts Child Psychiatry Access Project | Population |
| Academic literature database searches | Saunders 2021 | Factors associated with initiation of community-based therapy for emerging adults with mood and anxiety disorders | Study design/publication type |
| Academic literature database searches | Saxon 2023 | Telephone treatments in improving access to psychological therapies services: An analysis of use and impact on treatment uptake | Population |
| Academic literature database searches | Scanlon 2000 | Building capacity for promotion, prevention and early intervention in mental health | Study design/publication type |
| Academic literature database searches | Schell 2012 | Outdoor adventure for young people with a mental illness | Population |
| Academic literature database searches | Schmidt 2016 | Will a comprehensive, person-centered, team-based early intervention approach to first episode illness improve outcomes in eating disorders? | Population |
| Academic literature database searches | Schotanus-Dijkstra 2017 | An early intervention to promote well-being and flourishing and reduce anxiety and depression: A randomized controlled trial | Population |
| Academic literature database searches | Schweickle 2024 | Potential moderators and mediators of intervention effects in a sport-based mental health literacy and resilience program for adolescent men | Intervention |
| Academic literature database searches | Schweitzer 2023 | Developing an innovative pediatric integrated mental health care program: interdisciplinary team successes and challenges | Outcome |
| Academic literature database searches | Scott 2020 | Early intervention, prevention, and prediction in mood disorders: Tracking multidimensional outcomes in young people presenting for mental health care | Population |
| Academic literature database searches | Seaton 2022 | Mental health outcomes in patients with a long-term condition: Analysis of an Improving Access to Psychological Therapies service | Population |
| Academic literature database searches | Serneels 2017 | An Intervention Supporting the Mental Health of Children with a Refugee Background | Study design/publication type |
| Academic literature database searches | Shah 2023 | Return on investment from service transformation for young people experiencing mental health problems: Approach to economic evaluations in ACCESS Open Minds (Esprits ouverts), a multi-site pan-Canadian youth mental health project | Study design/publication type |
| Academic literature database searches | Sharifi 2023 | Effect of general practitioner training in a collaborative child mental health care program on children's mental health outcomes in a low-resource setting: A cluster randomized trial.DP - Jan 2023 | Population |
| Academic literature database searches | Shepherd 2009 | Nursing practice. Developing an outreach service in adolescent mental health to improve engagement | Study design/publication type |
| Academic literature database searches | Shidhaye 2016 | Development and piloting of a plan for integrating mental health in primary care in Sehore district, Madhya Pradesh, India | Study design/publication type |
| Academic literature database searches | Simmons 2021 | Inside the black box of youth participation and engagement: Development and implementation of an organization-wide strategy for Orygen, a national youth mental health organization in Australia | Study design/publication type |
| Academic literature database searches | Simmons 2023 | Implementing a combined individual placement and support  and vocational peer work program in integrated youth mental  health settings | Outcomes |
| Academic literature database searches | Simon 2020 | Development of 'learn to dare!': An online assessment and intervention platform for anxious children | Population |
| Academic literature database searches (update search) | Soliemannjad 2024 | Integrated Care for Adolescents in Child Welfare: Association of Extended Adverse Childhood Experiences with Youth Depressive Symptoms and Substance Use | Study design/Publication type |
| Academic literature database searches | Sookman 1999 | Integrative cognitive therapy for obsessive-compulsive disorder: A focus on multiple schemas | Study design/publication type |
| Grey literature database searches | Sorensen 2013 | Early detection and treatment of mental illness in the workplace: an intervention study | Study design/publication type |
| Academic literature database searches | Sorensen 2019 | An outreach collaborative model for early identification and treatment of mental disorder in Danish workplaces | Population |
| Academic literature database searches | Spencer 2019 | Utilization of Child Psychiatry Consultation Embedded in Primary Care for an Urban, Latino Population | Population |
| Academic literature database searches | Stadnick 2022 | Implementation outcomes from a pilot of "Access to Tailored Autism Integrated Care" for children with autism and mental health needs | Population |
| Academic literature database searches | Stafford 2020 | Improving access and flow within Child and Adolescent Mental Health Services: a collaborative learning system approach | Study design/publication type |
| Academic literature database searches | Stevens 2019 | Detecting and reducing post-traumatic stress among children exposed to domestic violence: A multi-agency early intervention program | Population |
| Academic literature database searches | Storch 2023 | Improving access in 2023: Evidence-based psychotherapy for autistic youth with anxiety | Study design/publication type |
| Academic literature database searches | Straus 2014 | Behavioral health care for children: the massachusetts child psychiatry access project | Study design/publication type |
| Academic literature database searches | Summerhurst 2017 | Youth perspectives on the mental health treatment process: What helps, what hinders? | Study design/publication type |
| Academic literature database searches | Szigethy 2001 | Depression among pregnant adolescents: An integrated treatment approach | Study design/publication type |
| Academic literature database searches | Szymanski 2013 | Integrated care: Treatment initiation following positive depression screens | Population |
| Academic literature database searches | Tandon 2015 | 21. An Intervention for Low-Income Adolescents and Young Adults in Employment Training Programs Reduces Depressive Symptoms and Improves Coping Strategies | Study design/publication type |
| Academic literature database searches | Tanzer 2021 | Implementing Coordinated Specialty Care in CMHC Youth and Young Adults with Severe Mental Illness: Preliminary Outcome Assessment | Population |
| Academic literature database searches | Tarnowski 1991 | DISADVANTAGED-CHILDREN AND FAMILIES IN PEDIATRIC PRIMARY CARE SETTINGS .1. BROADENING THE SCOPE OF INTEGRATED MENTAL-HEALTH-SERVICE | Study design/publication type |
| Academic literature database searches | Tay 2022 | Online HOPE intervention on help-seeking attitudes and intentions among young adults in Singapore: A randomized controlled trial and process evaluation | Intervention |
| Academic literature database searches | Taylor 2011 | Infusing Early Intervention for Substance Use Into Community Mental Health Services for Transitioning Youth | Population |
| Grey literature database searches | Teitelbaum 2005 | Integrating Mental Health Services for Adolescents in Juvenile Detention | Study design/publication type |
| Academic literature database searches | Theiss 2023 | Routine assessment of anxiety among adolescents in a primary care clinic | Intervention |
| Academic literature database searches (update search) | Thompson 2024 | Bridging the gap: Can single session interventions help enhance mental health treatment delivery for young people in Australia? | Study design/Publication type |
| Academic literature database searches | Thoren 2021 | The collaborative care model: Improving access to children's mental health care | Population |
| Academic literature database searches | Thummathai 2020 | Depression prevention in adolescents based on buddhism and sufficiency economy philosophy | Study design/publication type |
| Academic literature database searches | Titov 2017 | The first 30 months of the MindSpot Clinic: Evaluation of a national e-mental health service against project objectives | Population |
| Academic literature database searches | Tovey 2012 | The benefits of an adolescent psychiatric day service: the Harrow experience â€“ a pilot study | Population |
| Academic literature database searches | Towbin 2018 | Elements of Integrated Treatment of Refractory Major Depression in Youth | Study design/publication type |
| Academic literature database searches | Tucker 2016 | The Impact of Wilderness Therapy: Utilizing an Integrated Care Approach | Study design/publication type |
| Academic literature database searches | Turnbull 2023 | A pilot evaluation of the role of a children's wellbeing practitioner (CWP) in a child and adolescent mental health service (CAMHS) | Population |
| Academic literature database searches (update search) | Turpin 2024 | Leveraging integrated youth services for social prescribing: a case study of Youth Wellness Hubs Ontario | Outcomes |
| Academic literature database searches | Turton 2011 | Systematic Function-Based Intervention for Adolescents with Emotional and Behavioral Disorders in an Alternative Setting: Broadening the Context | Intervention |
| Academic literature database searches | Tyrer 1990 | A pilot study of the effects of early intervention on clinical symptoms and social functioning in psychiatric emergencies | Population |
| Academic literature database searches | Unknown | Team has cut waiting times for children | Study design/publication type |
| Academic literature database searches | Unknown | Specialization and integration in mental health care | Study design/publication type |
| Academic literature database searches | Unknown | Clinical digest. Early intervention could reduce suicide numbers in teenagers who self-harm | Study design/publication type |
| Grey literature database searches | Unknown 2006 | Research to Practice: Depression in the Lives of Early Head Start Families: Early Head Start Research and Evolution Project | Population |
| Academic literature database searches | Unknown 2007 | Program for early detection and referral of youth mental illness goes national | Study design/publication type |
| Grey literature database searches | Unknown 2008 | Evaluating the development and impact of Early Intervention Services (EIS) in the West Midlands | Population |
| Grey literature database searches | Unknown 2008 | Brief Integrative Therapy for Post-Traumatic Stress Disorder | Intervention |
| Academic literature database searches | Unknown 2009 | Early intervention key to prevent MH disorders in young people | Study design/publication type |
| Grey literature database searches | Unknown 2009 | Improving access to child and adolescent mental health services: reducing waiting times policy and practice guide (including guidance on the 18 weeks referral to treatment standard) | Study design/publication type |
| Grey literature database searches | Unknown 2009 | Improving the psychological wellbeing and mental health of children and young people: commissioning early intervention support services: requirements of PSA 12, indicator 4, 4th proxy measure | Study design/publication type |
| Academic literature database searches | Unknown 2010 | TORDIA depression trial suggests potential of early intervention for nonresponders | Population |
| Grey literature database searches | Unknown 2010 | Child and Family Traumatic Stress Intervention (CFTSI) | Study design/publication type |
| Academic literature database searches | Unknown 2012 | Program seeks to intervene before mental illness takes toll | Study design/publication type |
| Academic literature database searches | Unknown 2014 | Early intervention needed | Study design/publication type |
| Grey literature database searches | Unknown 2015 | Healthy minds, healthy Londoners: improving access to mental health services for London’s young and black, Asian and minority ethnic population | Study design/publication type |
| Academic literature database searches | Usacheva 2021 | Long-term mental health services use in children referred to a clinical intervention | Population |
| Academic literature database searches (update search) | van Doorn 2022 | Usability, feasibility, and effect of a biocueing intervention in addition to a moderated digital social therapy-platform in young people with emerging mental health problems: A mixed-method approach | Outcomes |
| Academic literature database searches | Vance 2023 | Exploring Service Use Disparities among Suicidal Black Youth in a Suicide Prevention Care Coordination Intervention | Intervention |
| Academic literature database searches | vanDoorn 2023 | The Effects of a Digital, Transdiagnostic, Clinically and Peer-Moderated Treatment Platform for Young People With Emerging Mental Health Complaints: Repeated Measures Within-Subjects Study | Intervention |
| Academic literature database searches | VanMeter 2019 | Online help-seeking prior to diagnosis: Can web-based resources reduce the duration of untreated mood disorders in young people? | Study design/publication type |
| Academic literature database searches | vanOrden 2009 | Collaborative mental health care versus care as usual in a primary care setting: A randomized controlled trial | Population |
| Academic literature database searches | VanVoorhees 2010 | Adolescents in primary care with sub-threshold depressed mood screened for participation in a depression prevention study: Co-morbidity and factors associated with depressive symptoms | Intervention |
| Academic literature database searches | Vella 2021 | An Intervention for Mental Health Literacy and Resilience in Organized Sports | Intervention |
| Academic literature database searches | Vickers 2013 | Integration of mental health resources in a primary care setting leads to increased provider satisfaction and patient access | Study design/publication type |
| Academic literature database searches | Vigano 2007 | Psychodynamic psychotherapy in the integrated treatment model for panic disorder: A 3-year follow-up clinical study | Intervention |
| Academic literature database searches | Vusio 2021 | After the storm, Solar comes out: A new service model for children and adolescent mental health | Study design/publication type |
| Academic literature database searches | Vyas 2015 | Youth services: Meeting the mental health needs of adolescents | Study design/publication type |
| Academic literature database searches | Wade 2007 | Early intervention services in youth mental health | Study design/publication type |
| Academic literature database searches | Waldfogel 1959 | LEARNING-PROBLEMS .3. A PROGRAM FOR EARLY INTERVENTION IN SCHOOL PHOBIA | Intervention |
| Academic literature database searches | Walitza 2020 | Early detection and intervention for obsessive-compulsive disorder in childhood and adolescence | Study design/publication type |
| Academic literature database searches | Wallace 2014 | Early Intervention for Young Children at Risk for Developmental Mental Health Disorders | Study design/publication type |
| Academic literature database searches | Walters 2007 | Do mail-shots improve access to primary care for young men with depression? | Intervention |
| Academic literature database searches | Wang 2020 | Rethinking service design for youth with mental health needs: The development of the Youth Wellness Centre, St. Joseph's Healthcare Hamilton | Study design/publication type |
| Academic literature database searches | Weersing 2008 | Brief Behavioral therapy for pediatric anxiety and depression: Piloting an integrated treatment approach | Intervention |
| Academic literature database searches | White 2022 | Bridging the gap: A new integrated early intervention service for young people with complex mental health issues | Population |
| Academic literature database searches | Wiener 2006 | Evaluation of a CAMHS in Primary Care Service for General Practice | Study design/publication type |
| Academic literature database searches | Wiles 2005 | DEVELOPING INTEGRATED MENTAL HEALTH SERVICES FOR CHILDREN AND YOUNG PEOPLE IN MORAY | Study design/publication type |
| Academic literature database searches | Wilson 2015 | Rural nurses: A convenient co-location strategy for the rural mental health care of young people | Population |
| Academic literature database searches | Wolfersdorf 2017 | [Early detection and early treatment of depressive disorder] | Study design/publication type |
| Academic literature database searches | Wolitzky-Taylor 2022 | Development and Initial Pilot Testing of a fully integrated treatment for comorbid social anxiety disorder and alcohol use disorder in a community-based SUD clinic setting | Study design/publication type |
| Academic literature database searches | Worrall-Davies 2004 | Evaluation of an early intervention Tier 2 child and adolescent mental health service | Study design/publication type |
| Academic literature database searches | Wright 2016 | The Costs and Cost-effectiveness of Collaborative Care for Adolescents With Depression in Primary Care Settings | Outcome |
| Academic literature database searches | Wright 2019 | Computerised cognitive-behavioural therapy for depression in adolescents: 12-month outcomes of a UK randomised controlled trial pilot study | Intervention |
| Academic literature database searches | Wright 2020 | Computerised cognitive behavioural therapy for depression in adolescents: 12-month outcomes of a UK randomised controlled trial pilot study | Intervention |
| Academic literature database searches | Wyatt 1983 | PARENTS AND MEDICAL-STUDENT THERAPISTS PERCEPTIONS OF CHILD MENTAL-HEALTH-SERVICES - A TEACHING PROGRAM IN PREVENTION AND EARLY INTERVENTION | Population |
| Academic literature database searches | Yearous 2008 | Adolescent mental health screening | Study design/publication type |
| Academic literature database searches | Yonek 2018 | Patient-centered medical home care for adolescents in need of mental health treatment | Study design/publication type |
| Academic literature database searches | Zenone 2022 | Implementing integrated-youth services virtually in British Columbia during the COVID-19 pandemic | Outcome |
| Academic literature database searches (update search) | Zhang 2022 | Screening depressive symptoms and incident major depressive disorder among Chinese community residents using a mobile app-based integrated mental health care model: Cohort study | Population |
