## Appendix 4 for "Approaches to early intervention for common mental health problems in young people: a systematic review"

### Appendix 4: Study characteristics by goal

| Intervention | Author, year and country | Study aim | Setting, target population and provider | Study sample | Lived experience involvement | Study design | Quality rating | Results |
| --- | --- | --- | --- | --- | --- | --- | --- | --- |
| Models aimed at making care more comprehensive and joined up | | | | | | | | |
| Name: Collaborative Care Model (CoCM) | Chang et al. (2023)  Country: USA | Evaluate outcomes of implementing behavioural health services into a rural and underserved paediatric primary care clinic, and to understand clinical staff’s perspectives regarding the Collaborative Care Model (CoCM) implementation. | Setting: Primary care  Target population: Adolescents aged 12-18 presenting to primary care meeting screening criteria on depression and/or anxiety screening measures.  Delivered by: A behavioural health care manager, psychiatric consultant and primary care provider. | n = 101 young people  **Demographics**  Age range: 11-19  Ages 11-13: n = 17 (16.8%);  Ages 14-16: n = 49 (48.5%);  Ages 17-19: n = 35 (34.7%);  Gender:  Male: n = 22 (21.8%); Female: n = 79 (78.2%)  Ethnicity: Caucasian: n = 101 (100%)  **Mental health problems:**  Depression only: 23 (22.8%);  Anxiety only: 44 (43.6%)  Both depression and anxiety: 34 (33.6%) | No | Pre-post single group study | 9 | RQ2:  Pre-post comparisons:  Patients enrolled in services showed significant improvement in anxiety and depression symptoms. During the 1.5-year timeframe, There was an average improvement rate of 83% for depression symptoms using the PHQ-9 and an average improvement rate of 76% for anxiety symptoms using the GAD-7. |
| Name: Collaborative Care Model (CoCM) | Khasanov et al. (2024)  Country: USA | To examine changes in suicidal ideation, depression, and anxiety following CoCM among patients with and without suicidal ideation, comparing differences in these changes across demographic subgroups; and examine the relationship between the amount of CoCM services provided and degree of symptom reduction among patients with and without suicidal ideation. | Setting: Primary care  Target population: Anyone with mild to moderate mental health problems, mainly anxiety, or alcohol misuse, and in this paper, those at risk of suicide  Delivered by: A behavioural health care manager, psychiatric consultant and primary care provider. | n = 3,487 of which 339 were aged 18-24  **Demographics**  Not reported for included population separately  **Mental health problems:**  100% of sample had symptoms of anxiety and depression. 62/339 young people also had suicidal ideation | No | Pre-post single group study | 19 | RQ2:  Pre-post comparisons:  Post intervention, significantly lower PHQ scores were reported for both participants with and without suicidal ideation.  With suicidal ideation: mean difference: 5.68 (p < .05)  Without suicidal ideation: mean difference: 4.20 (p < .05)  There were also significantly lower GAD scores for both participants with and without suicidal ideation.  With suicidal ideation: mean difference: 2.88 (p < .05)  Without suicidal ideation: mean difference: 4.14 (p < .05) |
| Name: Community-based mental health hub | Parry et al. (2023)  Country: UK | Evaluate the services offered by a low-intensity community mental health hub in one of the most deprived boroughs of England, UK. | Setting: Community-based  Target population: CYP aged 8-18 and their parents/carers  Delivered by: Paper mentions a 'staff team' but no further detail provided. | n = 2,384 (from clinical records)  n = 40 (in qualitative component)  **Demographics:**  *Quantitative component:*  Age: Not reported Gender:  Female:male = 47:39 ‘Other’: 0.1% “Prefer not to say”: 0.4% Ethnicity:  White: 59.4% White-British: 57.6% Pakistani: 4% Bangladeshi: 2%  *Qualitative component:*  Mean age: Not reported  Gender:  Female: n = 27 (68%);  Male: n = 10 (25%);  Non-binary: n = 3 (8%);  Ethnicity (n = 36 with completed data)  White British: n = 31 (86%);  Pakistani: n = 3 (8%);  Asian: n = 1 (3%);  Black British: n = 1 (3%) | No | Pre-post single group study with comparisons to before implementation | 13 | RQ1:  Pre-post comparisons:  In terms of the other key performance indicators of the Hub, waiting lists have reduced through the implementation of the Hub model from an average of eight weeks from referral to initial appointment to three weeks as of March 2021.  RQ2:  Pre-post comparisons:  YP-CORE10 scores indicated that the majority of young people who accessed the Hub experienced a benefit, with the severity of their reported symptoms reducing from “moderate” (M=16.4, SD=0.2) to “mild” (M=11.09, SD=0.2). Young people reported a significant reduction in psychological distress following attendance to the Hub (z=-18.419, p=< 0.001) with a medium effect size (r=0.0.62) and a median YPCORE10 score reduction from pre-intervention to post-intervention.  Predictors of RQ2 results: Neither *the type of presenting issue motivating the referral to the Hub* (rs=0.005, p=0.900; rs=-0.098, p=0.854), nor *the number of interventions attended* (rs=0.037, p=0.285; rs=0.059, p=0.639) were significantly related to the improvements reported by the service users (assessed via YPCORE10 and SCORE15), indicating that other aspects of the service offered (e.g. quality of the therapeutic support, the relationship between clinicians and service users, the contents of the interventions proposed) may have contributed to the large-scale improvements outlined by the outcome measures used.  RQ3:  Pre-post comparisons:  There was a significant difference in pre- (M=43.9, SD=7.3) and post- (M=31.25, SD=13.2) SCORE-15 scores, (z=-5.405, p=<0.001) indicating an improvement in young people’s perception of their family life. |
| Name: First Episode Mood and Anxiety Program (FEMAP) | Osuch et al. (2015; 2016)  Country: Canada | 1. Conduct a process evaluation of an early intervention program developed for mood and anxiety disorders among transition-age youths. 2. Determine if this delivery model was being implemented as intended in terms of assessment activities, treatment, and referrals and was providing an appropriate level of coverage of its target audience. | Setting: Community-based (face-to-face)  Target population: Youth aged 16-25 with concerns related to mood and/or anxiety with or without substance use.  Delivered by: a team of mental health professionals including licensed, master's-level social workers and psychologists. | Total n accepted into service = 390  N with symptom severity effectiveness outcomes compared pre-post = 88  **Demographics:**  Total sample accepted onto the programme:  Age: M = 19.2, SD = 2.7  Gender: Female n = 243 (61%)  Ethnicity:  White, not Hispanic: n = 339 (85%);  Black, not Hispanic: n = 4 (1%);  Hispanic: n = 3 (1%);  Asian/Pacific Islander: n = 19 (5%);  Native: n = 5 (1.3%); Other: n = 29 (7.3%)  **Mental health problems:**  Total sample accepted into the service:  Depression & anxiety: n = 133 (34%);  Depression: n = 120 (30%);  Anxiety: n = 62 (16%);  Bipolar: n = 38 (10%);  Substance use: n = 15 (4%);  Trauma/PTSD: n = 12 (3%)  Other (incl. ADHD): n = 3 (1%) | No | Process evaluation using a single group cohort and comparisons before implementation | 12 | RQ1:  Pre-post comparisons:  There was a 65% increase in requests for service between March 2013 and May 2015, with over 35 new contacts per month, over 80% of whom are appropriate for the service. However, this meant that with no resources forthcoming for stable clinical salaries and no way to hire new clinical staff, this has led to long wait times. The wait for entry into FEMAP changed from two weeks to over five months  RQ2:  Pre-post comparisons:  There were significant improvements in depression symptoms (MADRS scores) and anxiety symptoms (ASI scores) at 4-month follow-up in the first 88 FEMAP patients to be evaluated in their ongoing longitudinal, prospective treatment-effectiveness study (p < 0.001).  RQ3:  Pre-post comparisons:  There were significant improvements in functioning and quality of life scores (all p values <.001)  RQ4:  Estimated average cost of 4 months of treatment at FEMAP (based on the first 63 patients in the study using chart review)=$1634 per patient  Cost of psychiatric evaluation in ER using Form 1 of the mental health act in Ontario plus ER visit (based on 330 cases at the London Health Sciences Centre 2013-2014) = approximately $2188 per patient  Cost of being on the Ontario Disability Support Program for 4 months, as of September/October of 2014, = $4392 per patient.  Estimates therefore suggested that it is reasonable to suspect that effective, early, outpatient treatment of youth with mood/anxiety disorders may be more cost-effective. |
| Name: First Episode Mood and Anxiety Program (FEMAP) | Anderson et al. (2019)  Country: Canada | Evaluate the impact of an early intervention program for emerging adults with mood and anxiety disorders in the larger health system context, relative to standard care. | Setting: Community-based (face-to-face)  Target population: Young people aged 16-25 who had at least two physician billing claims or emergency department visits with a diagnostic code for a mood or anxiety disorder in any 12-month period or an inpatient hospitalisation with a primary discharge diagnosis of a mood or anxiety disorder.  Delivered by: Multidisciplinary team of mental health professionals (including psychiatrists and/or physician) | **Unmatched sample:** FEMAP users: n = 497 Non-users: n = 29,389  Demographics Age at index date: M = 19.3, SD = 2.4 Gender: Female n = 332 (67.8%) Ethnicity: not reported  **Mental health problems:** Mood disorder: n = 211 (42.5%)  Anxiety disorder: n = 286 (57.5%)  Prior alcohol related disorder: n = 14, (2.8%)  Prior substance related disorder: n = 16 (3.2%)  **Matched Sample:** FEMAP users: n = 490 Non-users: n = 967  **Demographics:**  Age at index date: M = 19.3, SD = 2.4 Gender: Female n = 338 (68%) F; Ethnicity: not reported  **Mental health problems:** Mood disorder: n = 209 (42.7%)  Anxiety disorder: n = 281 (57.3%)  Prior alcohol related disorder: n = 14 (2.9%)  Prior substance related disorder: n = 16 (3.3%) | No | Retrospective cohort study with control group | 25 | RQ1:  Contemporaneous control comparisons:  PFEMAP users had more rapid access to a psychiatrist relative to nonusers (hazard ratio [HR], 2.82; 95% confidence interval, 2.45 to 3.26; median time, 16 vs. 71 days). In the year following admission, FEMAP users also had lower rates of emergency department use for mental health reasons (HR, 0.73; 95% CI, 0.53 to 0.99). No difference in psychiatric hospitalisations between groups (HR, 0.80; 95% CI, 0.48 to 1.33), but the overall event rate was low (FEMAP users = 4.5%, nonusers = 5.6%). |
| Name: First Episode Mood and Anxiety Program (FEMAP) | Osuch et al. (2019)  Country: Canada | Evaluate uptake in an independent sample and investigate engagement and medium-term outcomes of a program for mood and anxiety disorders among emerging adults; the First Episode Mood and Anxiety Program (FEMAP) | Setting: Community-based (face-to-face)  Target population: Young people aged 16-25 with concerns related to mood or anxiety with or without substance use.  Delivered by: Psychiatrists, psychologists, social workers, addictions, and family therapists, and licensed clinicians. | Total n = 398;  Intervention arm: n = 370  Wait-list controls: n = 210  **Demographics:**  Age: M = 19.2, SD = 2.6  Gender: Female n = 249 (67%)  Ethnicity:  White Canadian/ American: n = 292 (80%)  White European: n = 85 (23%)  East Asian: n = 19 (5%)  Aboriginal: n = 16 (4%)  Middle Eastern: n = 14 (4%)  Black Canadian/ American: n = 12 (3%)  Latin American: n = 11 (3%)  South Asian (e.g., India, Sri Lanka, Pakistan): n = 8 (2%)  Black African: n = 5 (1%)  Indo-Caribbean: n = 5 (1%)  South-East Asian: n = 5 (1%)  Missing data: n = 3 (1%)  **Mental health problems:**  The group had clinically moderate levels of depression and clinically significant levels of dysfunction.  Substance use (one or two times per month or less): n = 265 (72%) | No | Pre-post study with post-hoc waitlist control comparison | 15 | RQ2:  Contemporaneous control comparisons:  When outcomes were compared post-hoc to a wait-list control group, the treatment group (n=210, no additional information) had significantly greater improvement in depression (F=80.64, df=1,383, p<.001) and anxiety (F=32.81, df=1,373, p<.001).  Pre-post comparisons:  Depression and anxiety both improved significantly (p<.001) in the treatment group.  RQ3:  Contemporaneous control comparisons:  When outcomes were compared post-hoc to a wait-list control group, the treatment group (n=210, no additional information) had significantly greater improvement in functioning (F=52.61, df=1,376, p<.001).  Pre-post comparisons:  Functioning and quality of life both improved significantly (p<.001) in the treatment group.  RQ6:  Satisfaction:  All but one participant (99%) gave the program a mean score more positive than neutral. 47% (N=91) of participants gave it the highest possible score. |
| Name: Mindspace Mayo | Corley et al. (2023)  Country: Ireland | Provide information about pathways to care and clinical response to community-based brief interventions for improving youth mental health through evaluating the Mindspace Mayo service. | Setting: Primary care (face-to-face and telehealth)  Target population: Young people aged 12-25 experiencing psychological distress or at-risk of developing mental health disorders.  Delivered by: Staff had various training backgrounds, including mental health nursing, social work, psychology (clinical, counselling, educational), counselling/psychotherapy and occupational therapy, with experience in mental health. | n = 1184  **Demographics:**  Age: M = 17.92, SD = 2.661  Gender:  Male: 426, (34.3%);  Female: 732 (58.9%);  Non-binary: 3 (0.2%)  **Mental health problems:** Psychological/affective disorders (including anxiety, low mood, low self-esteem, suicidal thoughts, social anxiety, panic attacks, thoughts of hurting oneself, stress, elated mood, isolating/withdrawal, grief/loss, sleep changes): n = 1080; (91%);  Anxiety: n = 1,015 (86%);  Behavioural (including alcohol use, drug use, gambling, behavioural problems, self-injury): 17% | No | Pre-post single group study | 19 | RQ2:  Pre-post comparisons:  After the intervention, psychological distress (measured using the CORE) was significantly reduced in both those aged 12-16 (t(306)=12.81, p=<.001, Cohens d = 0.53) and those aged over 17 (t(273)=15.20, p=<.001, Cohens d = 1.38) compared to pre-intervention  Predictors of RQ2 outcome:  Predictors of Psychological distress  *Living situation, referral pathway, and source of referral* were not significant predictors of improvement in psychological distress.  *Complexity of presenting issue:*  Was a significant predictor of post-intervention psychological distress in participants aged 17+ (Wald=2.04, p=.049) but not in participants aged 12-16 (p=.153). Presenting issue was not a significant predictor of post intervention psychological distress. |
| Name: Quality improvement intervention | Peters et al. (2018)  Country: USA | Implement and evaluate a system for improving use and show rates of mental health appointments with an integrated behavioural healthcare model. | Setting: Primary care (face-to-face, phone)  Target population: Young people aged 12-26 years seeking support from primary care for sexual and reproductive health care, menstrual concerns, sexuality and gender identity concerns, eating disorders, behavioural health concerns, and complex psychosocial situations or chronic care management.  Delivered by: A multidisciplinary team comprised of adolescent health care providers and social workers. | n = not reported  **Demographics:** Not reported  **Mental health problems:** Not reported | No | Quality improvement project with comparisons to before implementation | 10 | RQ6:  Pre-post comparisons:  Drop-out rate:  The first appointment show rate improved from a monthly average of 51% to 78%; the overall (new and returning) show rate improved from 67% to 77%. Use rates initially improved and then decreased because of an emphasis on scheduling only patients ready to engage in treatment, determined by provider interview |
| Name: Reaching Out to Adolescents in Distress (ROAD) intervention. | Richardson et al. (2014)  Country: USA | Determine whether a collaborative care intervention for adolescents with depression improves depressive outcomes compared with usual care. | Setting: Primary care (face-to-face, phone)  Target population: Adolescents who screened positive for depression in primary care.  Delivered by: Intervention components were delivered by depression care managers (DCMs), master’s-level clinicians employed by the study | Total n = 101  n (intervention group) = 50  n (control group) = 51  **Demographics:**  Age: M = 15.3, SD = 1.3  Gender: Female n = 73 (72%)  Ethnicity:  White: n = 70 (69%); Black: n = 5 (5%); Asian/Pacific Islander: n = 2 (2%); Other/multiracial: n = 24 (24%)  **Mental health problems:** Major Depression (K-SADS): n = 61 (60%) | No | RCT | 21 | RQ1:  Contemporaneous control comparisons:  Overall, 86% of patients in the intervention group received either psychotherapy or medications that met study quality standards, compared with 27% of the (TAU) control group. Intervention youth were significantly more likely than control youth to receive 4 or more psychotherapy sessions in the first 6 months of the study (84.0% vs 15.7%; OR = 28.2; 95% CI, 9.7–82.1; P < .001), but differences were nonsignificant during months 6 through 12 (20.0% vs 15.7%; OR = 1.3, 95% CI, 0.5–3.7; P = .57). Although intervention youth were significantly more likely than control youth to have received antidepressants in the first 6 months of the study (44.0% vs 17.7%; OR = 3.7, 95% CI, 1.5–9.1; P = .005), there were no significant differences between intervention and control youth in receipt of 90 days or more of antidepressants at either 6 or 12 months. There were also no significant differences between the groups in use of specialty mental health care through Group Health  RQ2:  Contemporaneous control comparisons:  Depression: There was a 8.5-point greater decrease in mean CDRS-R depression scores from baseline than control youth (95% CI, −13.4 to −3.6; P = .001) at 6 months, and a 9.4-point greater decrease from baseline at 12months (95% CI, −15.0 to −3.8;P = .001).  Depression remission rates: at 12 months was 50.4% (95% CI, 34.7%-66.1%) for intervention youth compared with 20.7% (95% CI, 8.2%-33.2%) for control youth.  Functioning: Differences between intervention and control youth were not significant at P ≤ .01 at 6 months (mean difference, −4.4; 95%CI, −8.4 to −0.5; P = .03)  or 12 months (mean difference, −4.3; 95% CI, −8.3 to −0.3; P = .04).  RQ6:  Satisfaction:  Contemporaneous control comparisons:  When patients were asked to report their satisfaction with the treatment, those in the intervention group were significantly more likely to be “moderately to very satisfied” with care at 6 months (85.8% vs 52.2%; OR = 5.6, 95% CI, 1.9–16.0; P = .001) but not at 12 months (82.2% vs 68.5%; OR = 2.1, 95% CI, 0.7–6.1; P = .16)  Drop-out rates:  Contemporaneous control comparisons:  7/50 dropped out of the intervention group, 2/51 dropped out of enhanced usual care. Reasons for this not provided. |
| Name: Remote Collaborative Depression Care (RCDC) | Martínez et al. (2018)  Country: Chile | To assess the feasibility, acceptability, and effectiveness of a Remote Collaborative Depression Care (RCDC) intervention for adolescents with major depressive disorder living in the Araucanía Region, Chile. | Setting: Primary care (telephone)  Target population: Young people aged 13-19 presenting to primary care with suspected major depressive disorder.  Delivered by: Primary care teams at the participating health centres, mental health specialists from Universidad de Chile, and a psychologist responsible for conducting the phone monitoring. | Total n = 143  Follow-up n = 138  **Demographics:**  Age: M = 15.4 years, SD 1.6;  Gender: Females: n = 116 (81.1%)  Ethnicity: Mapuche (indigenous people of the region): n = 34 (23%); the rest not reported  **Mental health problems:**  Suspected major depressive disorder: n = 143 (100%) | No | RCT | 17 | RQ2:  Contemporaneous control comparisons:  Depressive symptoms: There was no significant difference between participants who received the remote collaborative depression care intervention and control participants in improvement in depression at 12 weeks (Non-adjusted difference between means (95% CI): 1.1 (−2.9 to 5.2); Adjusted difference between means (95% CI): 1.5 (−2.4 to 5.6))  Psychological wellbeing: There was no significant difference between participants who received the remote collaborative depression care intervention and control participants in improvement in psychological wellbeing at 12 weeks (Non-adjusted difference between means: 0.8 (−3.9 to 5.8); Adjusted difference between means: 1.0 (−3.4 to 5.8)  Health related quality of life: There was no significant difference between participants who received the remote collaborative depression care intervention and control participants in improvement in psychological wellbeing at 12 weeks (Non-adjusted difference between means: 1.0 (−2.8 to 4.8); Adjusted difference between means: 1.2 (−3.2 to 5.2)  RQ6:  Adherence to treatment: No significant differences were observed across groups at 12-week follow-up in terms of adherence to pharmacological treatment (Fisher exact test P value=.98)  Satisfaction: There was no significant difference in satisfaction ratings between the intervention and control group across satisfaction with treatment, facilities, medical care and non-professional staff treatment. However, participants who received the intervention were significantly more satisfied with their psychological care (Wilcoxon rank-sum test P value=.04) |
| Name: Support with One-stop care on Demand for Adolescents and young adults in Adachi (SODA) | Uchino et al. (2022)  Country: Japan | Clarify the services of Support with One-stop care on Demand for Adolescents and young adults in Adachi (SODA), which have the potential to overcome the challenges faced by existing systems, focusing on the role of clinical case management (CCM). | Setting: Community-based (face-to-face, telephone, online video, outreach)  Target population: Young people aged 12-35 with a wide variety of difficulties – diagnosis of a mental illness not needed.  Delivered by: A multidisciplinary team consisting of psychiatrist, psychiatric social workers, psychologist, public health nurse and registered nurse. | n = 105  **Demographics:**  Age: M = 20.5, SD = 6.8;  Gender: Female: n = 58 (55.2%)  Ethnicity: Not reported  **Mental health problems:**  Neurotic disorder: n = 8 (53.3%);  Mood disorder: n = 2 (13.3%);  Psychotic disorder: n = 1 (6.7%);  Other: n = 4 (26.7%) | No | Case-control study | 4 | RQ1:  Pre-post comparisons:  The rate of the subjects receiving psychiatric treatments significantly increased from 42.9% at initial interview to 71.4% at 6 months (p = 0.031)  RQ3:  Pre-post comparisons:  The mean level of global functioning improved significantly, from 46.6 (SD = 15.9) at the initial interview session to 59.3 (SD = 11.0) at 6 months (p = .001). |
| Name: The Young Adult Service (YAS) | Bond & Power (2020)  Country: Ireland | 1. Describe sociodemographic characteristics, clinical characteristics, and treatment pathways of CYP attending the Youth Adult Service (YAS). 2. Explore which treatment or clinical factors may affect outcomes. 3. Examine engagement rates and possible factors that may predict dropouts before completing treatment and follow-up. | Setting: Community and inpatient-based (face-to-face)  Target population: Young people aged 18-25.  Delivered by: Multidisciplinary team consisting of psychiatry, psychology, nursing, occupational therapy, social work, family therapy and cognitive behavioural therapy professionals. The clinical lead was a consultant dual trained and registered as both a child and adolescent and adult psychiatrist. | n = 567  **Demographics:**  Age: M = 20.6, SD = 2.0;  Gender: Female n = 302 (53.3%);  Ethnicity (birth country):  Ireland: n = 551 (97.2%);  Outside Ireland: n = 16 (2.8%)  **Mental health problems:** Anxiety disorder: n = 104 (18.3%);  Eating disorder: n = 32 (5.6%); Mixed anxiety/depressive disorder: n = 86 (15.2%);  Mood disorder: n = 148 (26.1%);  Psychotic disorder: n = 38 (6.7%);  Stress related disorder: n = 37 (6.5%);  No Axis I disorder: n = 122 (21.5%);  Diagnosis of any comorbid Axis I or II disorder (e.g., substance dependence or developmental disorder): n = 368, (64.9%);  Diagnosis of Axis I comorbidity (other than addictions): n = 100 (17.6%);  Diagnosis of alcohol/substance abuse/dependence: n = 177 (31.2%); Diagnosis of personality disorder: n = 238 (42.0%); Diagnosis of developmental disorder: n = 93, (16.4%) | No | Retrospective (single group) cohort study | 18 | RQ2:  Pre-post comparisons:  356 out of 567 (62.8%) were recorded as improving on the Clinical Global Impression scale between baseline and post-treatment, while 211 out of 567 (37.2%) were recorded as worsened or without improvement.  Predictors of RQ2 outcome:  Predictors of mental health symptoms:  *Employment:*  Unemployment was significantly associated with having equivocal or worse outcomes (X2 = 6.6, p=.007)  *Route to, and receipt of care:*  Patients who had been referred from inpatient (compared to outpatient) services (X2 =25, p<.001) and who had at least one admission during their time in the young adult service (X2 =24, p<.001) were more likely to show improvement  *Diagnosis:*  There was no effect of diagnosis on outcomes, including the impact of comorbidities, however, there was an impact of attending a treatment programme specifically developed for their diagnosis for all diagnoses except anxiety (p<.05 for all except anxiety).  *Medication:*  Patients treated with medication had higher rates of improvement compared with those who had never received medication (X2 =10.2, p=.001)  Predictors of RQ6 outcome:  Drop-out  *Route to, and receipt of care:*  Time spent in the service positively predicted drop-out (OR=1.01, p< .001), while directly transferring from CAMHS (OR=2.01, p=.044), having a developmental disorder (OR=2.66, p=.001) and being prescribed psychotropic medication (OR=1.73, p=.012) negatively predicted drop-out. |
| Name: Youth Partners in Care Quality Improvement Intervention | Asarnow et al. (2005; 2009); Ngo et al. (2009); Wells et al. (2012); Rapp et al. (2017)  Country: USA | 1. Evaluate the effectiveness of a quality improvement intervention aimed at increasing access to evidence-based treatments for depression (particularly CBT and antidepressant medication), relative to usual care, among adolescents in primary care practices. 2. Explore racial-ethnic differences in impact of the intervention and predictors of access to care. | Setting: Primary care (face-to-face)  Target population: Youths aged 13-21 presenting at clinics for a primary care visit with depression.  Delivered by: Master's- or doctoral-level care managers who supported primary care providers (clinicians) with patient evaluation, education, medication, and psychosocial treatment and linkage to specialty mental health services. Cultural researchers provided consultation to primary care providers. | Total n = 418 Quality Improvement (QI) arm: n = 211  Usual care (UC) arm: n = 207  **Demographics:**  Age: M = 17.2; SD = 2.1;  Gender: Female n = 326 (78%);  Ethnicity:  African American: n = 56 (13.4%);  Asian: n = 5 (1.2%);  Hispanic: n = 234 (56%);  Mixed: n = 57 (13.6%);  White: n = 53 (12.7%);  Other: n = 13 (3.1%)  **Mental health problems:**  Depression: n = 178 (42.6%);  Externalising symptoms or conduct problems: n = 117 (28%);  PTSD symptoms: n = 93 (22.3%);  Substance use: n = 103 (24.6%);  Suicide attempt or deliberate self-harm: n = 54 (12.9%); | No | RCT | 19 | RQ1:  Contemporaneous control comparisons:  At 6-month follow-up, participants who had received the QI intervention were significantly more likely to receive mental health care than those receiving usual care (32% vs 17%; OR=2.8 (95% CI: 1.6 to 4.9, p<.001). This was due to higher rates of psychotherapy (Arsarnow 2005) However by 18 months there was no significant difference between the two groups (p=.427)  Participants who received the QI intervention were more likely to have "appropriate treatment" (defined by use of guideline recommended antidepressants or 6 or more speciality counselling visits over 6 months) than the usual care control group (probit coefficient=0.32 (SE=0.13), t=2.46, p=0.01) at 6 months follow up. This remained when broadening appropriate treatment to include a mental health specialty visit (p<.001) (Wells 2012)  Impact of the intervention in sub-groups (ethnicity):  At six months follow-up, more participants from minority ethnic groups accessed mental health treatment from the QI intervention arm than the usual care control arm. Black participants were significantly more likely to receive any counselling (37% vs 18%, OR=3.65, 95% CI: 1.09-12.23, p=.036) and to receive speciality mental health care (42% vs 9%, OR=9.37, 95% CI: 1.58-55.71, p=.014). Latino participants were more likely to receive any speciality mental health care (28% vs 17%, OR: 2.03, 95% CI: 1.01-4.06, p=0.046). However, there were no significant differences for any ethnic group between the intervention and control group for use of any mental health care by a primary care clinician or medication. (Ngo 2009)  Impact of the intervention in sub-groups (language): Treatment access rates were higher in the QI intervention group for those whose primary language was English, however, they were not in participants with an alternative primary language (p=.023) (Rapp 2017)  Impact of the intervention in sub-groups (age):  At six months follow-up, the quality improvement intervention mitigated the effect of older age on reduced treatment access (p<.001) (Rapp 2017)  RQ2:  Contemporaneous control comparisons  Mental health symptoms: Participants who received the QI intervention had significantly lower depression scores (measured using the CES-D) at six months follow up compared to the usual care control group (QI intervention: M=19, SD=11.9, control: M=21.4, SD=13.1, p=.02). The participants in the QI intervention group were also significantly less likely to reach the threshold for severe depression (CES-D score of ≥24) compared to the control group (QI intervention: 31%, Control: 42%; OR=0.6, 95% CI: 0.4 to 0.9, p = .02) (Arsarnow 2005; 2009)  However, at the 18 month follow up, There were no differences between the two groups in mental health symptom scores and the trend towards decreased rates of severe depression in participants who recieved the QI intervention no longer reached statistical significance (p=.06) (Arsarnow 2009)  Suicide and self harm: There was no significant difference between the two groups in youth-reported suicidal ideation and suicide attempts or deliberate self-harm at either 6 or 18 months. (Arsarnow 2005; 2009)  Impact of the intervention in sub-groups (ethnicity):  At six months follow-up, black participants had significantly reduced depression scores in the QI intervention group but not in the usual care control group (difference between groups = -7.55, CI: -12.17 to -2.93, p=.001), however there was no effect of the intervention in Latino participants (p=.191) or white participants (p=.959) (Ngo 2009)  Wellbeing and quality of life: The participants in the QI intervention reported higher mental health–related quality of life compared with the usual care control group at 6 months follow up (QI intervention: M=44.6, SD=11.3, control: M=42.8, SD=12.9, p=.03) (Arsarnow 2005; 2009), However at 18 months follow up there were no significant differences in mental health-related quality of life (p=.245) (Arsarnow 2009)  RQ2: Quality of life  Impact of the intervention in sub-groups (ethnicity):  There was no differential effect of the intervention in specific ethnic groups (p=.195, p=.069 and p=.213 for Black, Latino and White participants, respectively) (Ngo 2009)  RQ6:  Contemporaneous control comparisons  Participants who received the QI intervention reported significantly higher satisfaction with care scores at 6 months follow up compared to the usual care control M=3.8, SD=0.9, control: M=3.5, SD=1.0, p=.004) (Arsarnow 2005; 2009) However by 18 months follow up there was no longer a significant difference in satisfaction with care between the two groups (p=0.280) (Arsarnow 2009)  Impact of the intervention in sub-groups (ethnicity):  Only the Latino sub-group in the QI intervention had significantly higher ratings of satisfaction at 6 months follow up compared to Latino participants in the control group (mean difference =0.28, ci: 0.06-0.80, p=.015) (Ngo 2009). |
| Name: Jigsaw | O’Keeffe et al. (2015)  Country: Ireland | Profile the young people who avail of a service in Jigsaw and to provide emerging evidence that Jigsaw’s goal-focused early intervention model facilitates the reduction of psychological distress. | Setting: Community-based (face-to-face)  Target population: Young people aged 12-25 with mild to moderate mental health difficulties.  Delivered by: Staff consists of multi-disciplinary teams of allied health professionals with varying professional backgrounds. | n = 2,420  **Demographics:**  Age: Not reported  Ethnicity: Not reported  Gender: Female: n = 1367 (56.5%)  **Mental health problems:** Anxiety: 33.5%  Feelings of depression: 19.35%  Drug use: 7.55%  Thoughts of hurting self: 13.95% | No | Pre-post single group study | 16 | RQ2:  Pre-post comparisons:  Psychological distress:  CORE-10: Young people showed a significant difference in pre- and post-intervention levels of psychological distress as measured by the CORE-10, t(157) = 22.249; p < 0.001, CI95 = 11.63, 13.89 (Cohen’s d = 0.8), with lower levels of psychological distress reported post-intervention (M = 7.56; S.D. = 5.37) compared with pre-intervention (M = 20.33; S.D. = 6.92). 62% of young people showed a reliable and clinically significant improvement on the CORE-10. 22% showed a reliable improvement only.  YP-CORE: Young people also showed a significant reduction in psychological distress on the YP-CORE, t(149) = 16.839; p < 0.001. CI95 = 9.33, 11.81 (Cohen’s d = 0.7), with scores being significantly lower post-intervention (M = 8.98; S.D. = 7.05) compared with pre-intervention (M = 19.55; S.D. = 6.92). 68% participants showed a reliable improvement on the YP-CORE.    Predictors of RQ2 outcomes:  There was no significant difference in changes in psychological distress by age and gender.  RQ5: There was a consistent finding of “definite agreement” with the evaluative questions on the post-treatment qualitative rating scales for youths.  Satisfaction: 95% youths completing the scales indicated that they were entirely satisfied with their experience in the BIC. |
| Name: Jigsaw | O’Reilly et al. (2022)  Country: Ireland | 1. Examine the effectiveness of Jigsaw’s brief intervention model of support using an idiographic tool, the goal-based outcome (GBO) measure. 2. Explore the type of goals set by young people engaging with this service. | Setting: Community-based (face-to-face, video, and phone)  Target population: Young people aged 12-25 with mild to moderate mental health difficulties.  Delivered by: Jigsaw clinicians, who are trained mental health professionals. | n = 4839  **Demographics:**  Age (years) (N, %):  12–14 = 1506 (31.1%)  15–17 = 2045, (42.3%)  18–21 = 933, (19.3%)  22–25 = 355, (7.3%)  Gender:  Female = 3072 (63.5%)  Male = 1767 (36.5%)  **Mental health problems:**  Diagnoses not reported. Focuses on young people experiencing mild to moderate mental health difficulties. | No | Pre-post single group study | 17 | RQ3:  Pre-post comparisons:  There was a substantial main effect for time, with all demographic groups showing an increase in Goal-Based Outcome scores from pre- to postintervention. All models show a significant effect of time, which were reported by different participant groups: Gender: F(1, 3308) = 11,221.74, p < .001, gp2 = .77; Age: (1, 3306) = 7753.17, p < .001, gp2 = .70; Referral source: F(1, 3305) = 4000.47, p < .001, gp2 = .54  Over three-quarters (78.7%) of participants showed reliable improvement in progress towards their goals, 21.1% showed no reliable change, and 0.2% displayed a reliable deterioration.  Predictors of RQ3:  Changes in goal-based outcome scores from pre- to post-intervention did not vary significantly by age group, gender or referral source (self-referral vs family/friends vs medical). However, there was a significant association between referral source and reliable change categories; those referred by school were significantly more likely to fall into the reliable deterioration category. |
| Models aimed at increasing speed or ease of access to support | | | | | | | |  |
| Intervention: Enhanced Moderated Online Social Therapy (MOST+) | Alvarez-Jimenez et al. (2020)  Country: Australia | Primary: Determine the feasibility, acceptability, and safety of MOST+ for young people seeking online mental health support.  Secondary:   1. Assess changes in psychological distress, well-being, depression, stress, social support, loneliness, basic psychological needs (self-competence, relatedness, and autonomy), strengths usage, and mindfulness skills from the point of engagement to post intervention 2. Explore the associations between system usage, perceived helpfulness, and secondary outcome variables. | Setting: Online  Target population: Young people aged 16 - 25 years who are help-seeking with concerns about their own mental health  Delivered by: Web counselling provided by qualified clinicians. Moderation provided by clinician and peer moderators. | n = 157 young people  Age: 19.1 (2.3), Gender: 77% female  Ethnicity: Aboriginals and/or Torres Strait Islander: 3%  Reasons for help-seeking:  Sadness: n = 60 (38%) Anxiety: n = 35 (22%) Distress: n = 15 (9.6%)  Previous support: n = 93 (59%) had not previously used mental health services n = 58 (37%) had never received any mental health support | Yes – ‘end users’ were involved in the development of the MOST+ platform, and trained peer moderators (young people with lived experience of mental illness) moderated the online social networking platform. | Pre-post single group study | 12 | RQ2:  Pre-post comparisons  Psychological distress: There was a significant reduction in psychological distress between baseline and post-intervention (d=−0.39; p<.001). The proportion of participants with psychological distress scores on the K10 which were indicative of severe mental health disorder was significantly lower after the intervention (72%) compared with baseline (82%; X2(91)=18.8; p<.001).  Depression: There was a significant decrease in depression between baseline and post-intervention (d=−0.29; p=.008). The proportion of participants with depression scores on the PHQ-9 which were indicative of moderately severe depression was significantly lower after the intervention (53%) compared with baseline (58%; X2(91)=19.7; p<.001).  Perceived stress: There was a significant reduction in perceived stress between baseline and post-intervention (d=−0.44; p<.001).  Psychological well-being: There was a significant improvement in psychological well-being between baseline and post-intervention (d=0.51; p<.001).  Predictors of RQ2:  Analyses showed similar results when only including participants who had received full vs partial access to all available aspects of the intervention based on safety assessments at intake.  RQ3:  Pre-post comparisons  Loneliness: Loneliness significantly improved between baseline and post-treatment (d=−0.23; p=.04).  Social support: Social support significantly increased between baseline and post-treatment (d=0.30; p<.001).  Autonomy: Autonomy significantly increased between baseline and post-treatment (d=0.36; P=.001).  Self-competence: Self-competency significantly improved between baseline and post-intervention (d=0.30; P<.001).  Predictors of RQ3:  Analyses which separated those given full vs partial access to the intervention showed that those given full access only showed improvement in loneliness (d=−0.33; p=.02), social support (d=0.25; p=.05), and autonomy (d=0.50; p<.001), while those given partial access only showed significant improvements in social support (d=0.39; p=.03), and self-competence (d=0.42; p<.001).  RQ6:  Self-report satisfaction  Participants gave positive ratings of their experience using MOST+, with mean scores of 3.5 or more (out of 5) on each of the core domains of ease of use (M=3.7, SD=1.1), relevancy (M=3.9, SD=1.0), helpfulness (M=3.5, SD=0.9), and overall experience (M=3.9, SD=0.8).  98% (91/93) of participants reported a positive experience using MOST+.  86% (80/93) considered it easy to use.  88% (88/93) reported that MOST+ was relevant to their needs.  86% (80/93) considered it helpful, 82% (76/93) reported that using MOST+ helped them feel better.  86% (70/93) felt more socially connected using it.  92% (86/93) said that they would recommend it to other young people experiencing similar difficulties. |
| Name: Acceptance and Commitment Therapy implemented within Integrated Primary Care (ACT-IPC) | O’Dell et al. (2020)  Country: USA | Preliminary effectiveness study of a novel acceptance and commitment therapy group treatment implemented within paediatric integrated primary care (ACT-IPC) to address limited access to behavioural health services in primary care. | Setting: Primary care (face-to-face)  Target population: Youth aged 12-19 attending an integrated primary care clinic in rural USA  Delivered by: Each group was co-facilitated by a licensed psychologist and postdoctoral fellow. | n = 110  **Demographics:** Age: M = 15.1 (SD = 1.5)  Gender: Female: n = 90 (81.8%)  Ethnicity:  White, non-hispanic: 103 (93.6%)  Black, non-hispanic: 4 (3.6%) Hispanic: 3 (2.7%)  **Mental health problems:** Primary diagnosis: Adjustment disorder: 40 (36.4%), Depression: 34 (30.9%) Anxiety: 31 (28.2%)  Conduct disorder: 4 (3.6%)  ADHD: 1 (0.91%). | Yes – the initial intervention protocol was refined over several group cycles based on feedback from adolescent group members | Retrospective single group cohort study | 18 | RQ2:  Pre-post comparisons:  Anxiety: Anxiety significantly reduced post-intervention (RCADS Anxiety: mean change -7.35 (SE: 1.58). D=-0.52, p<.001).  Depression: Depression significantly reduced post-intervention (RCADS Depression: mean change -5.15 (SE: 1.43). D=-0.54, p<.001).  Predictors of RQ2 outcomes:  Age, sex, race and ethnicity, primary diagnosis, psychiatric comorbidity, psychotropic medication, concurrent individual psychotherapy, overall participation by clinic, average number of sessions, and which ACT-IPC group a participant attended did not significantly predict RCADS Anxiety or RCADS Depression subscale scores. |
| Name: Entourage | Rice et al. (2020)  Country: Australia | Examine the acceptability, feasibility, safety and potential clinical benefit of a novel digital intervention for young people with social anxiety. | Setting: Community-based and online digital  Target population: Young people aged 12–25 years inclusive with “probable” social phobia.  Delivered by:  Entourage platform: use is self-directed with moderation from expert youth mental health clinicians. Peer support is also provided by peer support workers who completed a one-day workshop of training specific to providing peer support in an online environment.  Concomitant in-person therapy at Headspace centres is delivered by psychologists, occupational therapists, social workers and psychiatrists. | n = 89  **Demographics:**  Age: M = 19.8 years (SD not reported)  Gender:  Male: n = 43 (48.3%);  Female: n = 42 (47.2%); Non-conforming: n = 4 (4.5%)  Ethnicity:  Born in Australia: n = 65 (73%)  **Mental health problems:**  Mild anxiety: 9.9%; Moderate anxiety: 12.3%;  Marked anxiety symptoms: 21.0%;  Severe social anxiety: 56.8%  Suicide ideation: 40.4%  Minimal depression: 6.8%;  Mild depression: 13.6%;  Moderate depression:  31.8%;  Moderately severe depression: 30.7%;  Severe depression: 17.0% | Yes – Entourage represents an adaptation of the MOST model (Alvarez-Jimenez et al., 2016), an online platform developed in collaboration between a multidisciplinary team and young people experiencing mental health problems. The Entourage intervention also includes moderation and support from peer workers. | Pre-post single group study | 17 | RQ2:  Pre-post comparisons:  Anxiety: largest clinical improvement observed for social anxiety symptoms (LSAS); with effect size of d = 0.73 (p < .001) and 48.33% (n = 29) of participants showing reliable improvement. n = 4 (6.67%) participants experienced reliable worsening of social anxiety symptoms (LSAS).  Depression: Young people’s symptoms of depression significantly decreased (PHQ-9: d = 0.66, p < .001). n = 2 (2.74%) participants showed reliable deterioration in depressive symptoms (PHQ-9).  Suicidality: Young people’s suicidality significantly reduced (PHQ-9 suicidality item: d = 0.27, p = .026).  Wellbeing: Young people’s wellbeing significantly increased (SWEMWBS; d = 0.50, p < .001)  Self-esteem: Young people’s self-esteem significantly improved (RSES; d = 0.47, p < .001).  RQ3:  Pre-post comparisons:  Loneliness, as measured by the UCLA, had a significant and reliable improvement (d = 0.63, p < .001, 39.13% improved). Social connectedness as measured by the Social Connectedness Scale showed an effect size of d = 0.63 (p < .001) and 44.12% of participants had significantly improved during the intervention period. Participants showed decreased feelings of thwarted belongingness, measured by the Interpersonal Needs Questionnaire (INQ; d = 0.58, p < .001) with 32.94% showing significant improvement.  Predictors of RQ2 and RQ3 outcomes:  *Gender differences:* Overall, males showed reliable improvement on 14/22 variables, whereas non-male young people reliably improved on 18/22 variables. Males had a slightly lower median effect size overall (Males med. d = 0.39, non-males med. d = 0.56).  *Use of intervention:*  Participant ratings of the development of new social connections occurring via Entourage were positively associated with logins to the system (rs = 0.39, p = .001), number of therapy steps completed (rs = 0.31, p = .007), number of actions completed (rs = 0.35, p = .002), as was the usage composite score summing logins, steps and actions (rs = 0.37, p = .001).  RQ6:  Acceptability:  A total of 25.8% (n = 23) participants met the a priori acceptability criteria (logging on to Entourage at least 10 times over 10 different weeks), however 60.7% (n = 54) logged in weekly over 5 weeks. In addition, at post treatment, 74.4% of participants reported that Entourage provided timely support; 62.2% of participants reported the therapy content in Entourage was relevant to developing control over social anxiety symptoms; and 77.0% of participants found Entourage to be at least somewhat helpful.  Safety:  All participants reported feeling safe and adequately supported by clinicians on the Entourage intervention: measured using a 5-point Likert scale from “not at all safe” to “very safe”, 94.1% (n = 32) of participants reported feeling safe while using the Entourage intervention and no participants reported feeling unsafe. No serious adverse events were reported during the intervention. |
| Name: Headspace Brief Intervention Clinic (BIC) | Schley et al. (2019)  Country: Australia | Not clearly stated. | Setting: Community-based  Target population: Young people aged 12-25 presenting with a general mental health problem or mild to moderate levels of mental disorder, low level of risk to self/others and limited complexity (i.e. "simple" rather than multiple needs).  Delivered by: Initial headspace biopsychosocial assessment is conducted by a postgraduate mental health clinician. The BIC intervention was delivered by trained allied-health trainee clinicians trained who received individual and group supervision with a senior clinical psychologist. | n = 122  **Demographics:**  Age:  The "younger" cohort of 12-17 year olds made up roughly 70% of the BIC graduates  Gender:  Identifying as male gender: 62%  Identifying as female gender: 38%  Identifying as other gender = 0%  Ethnicity: not reported  **Mental health problems:**   - 17% presented with either no specific symptoms of mental disorder or mild-moderate general symptoms of mental health problems. - 27.1% young people who were engaged with the BIC had subthreshold mental disorders or met criteria for one or more diagnosable mental illnesses. | Yes – the BIC was refined with input from Youth Advisory Groups | Retrospective observational study with comparison to before implementation | 13 | RQ1:  Pre-post comparisons:  Time to starting treatment: The time between initial contact with the centre and commencing treatment improved to M = 17.6 days; SD = 11.9; min = 0; max = 49. Before, this was M = 5.31 weeks; SD = 3.23, min = 0.14; max = 14.  RQ2:  Pre-post comparisons:  Overall: Significantly more young people scored in the “non-clinical” range on a given psychometric measure compared to the time of the initial session.  Clinical stage of illness: More young people were in a "lower" clinical stage of illness by the end of their treatment in the BIC, but this shift was not significant (λ2 = 15.19; df = 12; P = 0.231).  Psychological distress: There were significant reductions on K10 (d = 0.75; p < 0.000), QIDS (d = 0.66; p < 0.000) and OASIS (d = 0.61; p < 0.000) following treatment. These were medium effect sizes.  Predictors of RQ2 outcomes  Gender, age, centre attended did not predict changes in symptoms, except for a significant correlation between centre and K10 scores. Authors state this is likely a reflection of the very low numbers of young people with available data at this centre (n = 5) rather than a true representation for "worse outcome".  RQ3  Social and occupational functioning: Scores on the SOFAS significantly improved following treatment (d = 0.59; p < 0.000). This was a medium effect size.  Following involvement in the intervention, significantly more young people scored in the “non-clinical” range on a given psychometric measure compared to the time of the initial session.  Predictors of RQ3 outcomes  The occupational and social functioning of young people improved equally across age groups, genders, and centre attended, irrespectively of the number of completed sessions and whether participants dropped out or completed treatment in a planned manner. |
| Name: RISE Rugby League Development Programme | Dowell et al. (2020)  Country: Australia | 1. Examine mental health outcomes and program acceptability in 12-15-year-old adolescent boys who participated in RISE within an urban versus a rural setting. 2. Describe implementation feasibility, defined in terms of participant attrition, obstacles, and challenges. | Setting: Community-based (face-to-face)  Target population: Male rugby league players aged 12-15 years old.  Delivered by: Clinical psychologists | Total n = 74 Urban areas: n = 44 Rural areas: n = 30  Completed follow up: Total n = 36 Urba cohort: n = 24  Rural cohort: n = 12  **Demographics:**  Age:  Urban: M = 13.29, SD = 0.95 Rural: M = 13.67, SD = 0.99;  Non-completers: M = 13.38, SD = 0.97;  Gender: 100% male  Ethnicity (Country of birth):  Australia:  Urban: n = 2 (87.5%);  Rural: n = 12 (100%); Non-completers: n = 18 (85.7%);  New Zealand:  Urban n = 3 (12.5%);  Rural n = 0;  Non-completers: n = 1 (4.8%);  Great Britain:  Urban: n = 0;  Rural: n = 0;  Non-completers: n = 2 (9.5%)  **Mental health problems:** Not specified | No | Pre-post single group study | 9 | RQ2:  Pre-post comparisons  Anxiety: Mean anxiety scores significantly declined from pre- to post-intervention for both the urban and rural site (F(1,34)=6.25, p=0.017, η2=0.16), but there was no significant difference in the decline in anxiety between the urban and rural sites (F(1,34)=0.25, p=0.62, η2=0.01).  Depression: There was a marginally significant decrease in depression symptoms over time, from pre- to post-assessment (F(1,34)=3.97, p=0.054, η2=0.11), but the decline in depression did not significantly differ between the urban and rural sites (F(1,34)=2.30, p=0.13, η2=0.07).  Anger and conduct: There were no significant changes in anger or conduct problems at either the urbal or rural site (F(1,34) = 0.65, p = 0.43, η2 = 0.02), and no significant differences in changes in anger or conduct problems between the urban and rural site (F(1,34)=1.46, p=0.24, η2=0.04).  Parents’ ratings of their child’s mental health: 5/7 contactable parents (71%) of high-risk participants reported some or much improvement in their son's mental health. 2/7 (29%) reported no change.  At post-assessment, 5/8 (63%) participants identified as high-risk in the pre-assessment were in the normal range on all areas that were in the risk-range at pre-assessment, and 3/8 (37%) remained in the high-risk range for one area at post-assessment (two of these participants for anger/conduct problems, and one for anxiety).  RQ6:  Contemporaneous control comparisons:  Acceptability  The urban boys rated the sessions as significantly more enjoyable than rural boys (t(68) = 2.29, p = 0.025).  There were no significant differences between sites for helpfulness of sessions, ease of understanding content, or usefulness of workbook content (all p's > 0.12).  The mean session ratings were within the 'a lot' to 'very much' range on average at both sites. |
| Name: RISE Rugby League Development Programme | Waters et al. (2022)  Country: Australia | 1. Compare changes in strengths-based attributes from pre-to post-assessment 2. Determine whether supporting parents of players participating in the RISE program who were at high-risk for mental health problems to engage in and access further care reduced the mental health symptoms of players to within healthy ranges. | Setting: Community-based (face-to-face workshops in the clubhouse of the rugby league groups, Access element had online and phone-based referral options)  Target population: 12–15-year-old boys participating in a rugby league development programme.  Delivered by: Workshops were co-delivered by two of three provisionally registered psychologists completing postgraduate clinical psychology training at Griffith University. | Total n = 251;  Included in study: n = 176;  RISE group/ intervention group: n = 94;  Control group/group who did not receive the RISE/Life-Fit-Learning program: n = 82.  **Demographics:**  *Intervention group:* Age: M = 13.12, SD = 0.788  Gender: Female n = 0 (0%);  *Comparison group:* Age: M = 13.32, SD = 1.029  Gender: Female n = 0, (0%)  Ethnicity: Not reported for either group.  **Mental health problems:** *Intervention group:*  Of those 94 RISE participants, 24 players (25.5%) reported anxiety, depression and/or behavioural problems scores in the high-risk range (i.e., RISE high-risk participants) and 70 players were in the healthy range on all measures (i.e., RISE healthy range participants);  *Comparison group:* Of those 82 controls, 21 players (25.6%) self-reported scores in the high-risk range for anxiety, depression and/or behavioural problems (i.e., comparison high-risk participants) and 61 were in the healthy range (i.e., comparison healthy range participants). | Yes – states in the introduction that community-based participatory research underpins their approach, and that they adopt a collaborative approach to research and program delivery that involves all partners in the process. No specific details provided. | Non-randomised controlled trial | 14 | RQ2:  Contemporaneous control:  Anxiety: Anxiety symptoms significantly reduced from pre- to post-assessment in participants categorised as 'high risk' in the RISE (p = 0.02) and comparison (p = 0.017) groups. Anxiety symptoms did not significantly change amongst 'healthy range' participants in either the RISE or comparison group (both p > 0.35).  Depression: Depression symptoms significantly improved over time in the intervention group participants classified as 'high risk' (p = 0.016) but not the control group participants classified as 'high risk' (p = 0.12). Depression symptoms did not significantly change in healthy range participants in the RISE (p = 0.63) or comparison (p = 0.19) groups.  Behavioural problems: Behavioural problems improved significantly in the RISE (p = 0.003) but not the comparison (p = 0.13) high risk groups over time.  Behavioural problems did not significantly change in the RISE healthy range participants (p = 0.23) but did significantly increase in the comparison healthy range participants (p = 0.019).  Overall: In both groups, 26% of players scored in the high-risk range on at least one mental health measure.  Predictors of RQ2:  *Parental action*  Among the RISE high-risk participants, it was observed that parent-reported and youth self-reported improvements were greater for players whose parents took more action, rather than less, to assist their children between pre- and post-assessment.  RQ6:  Contemporaneous control:  Self-satisfaction:  Self-satisfaction declined over time in comparator group, but did not decline over time in the RISE intervention group (Time X Condition ANOVA, f (1,174)=4.85, p=0.029. The results showed that self-satisfaction significantly declined from pre- to post-assessment in the comparison group (p = 0.01), which differed from the RISE group who did not change on self-satisfaction over time (p = 0.66). |
| Name: RISE Rugby League Development Programme | Waters et al. (2024)  Country: Australia | Determine the comparative effectiveness of in-person and remote delivery of the Life-Fit system within a junior sport development program. | Setting: Community-based (face-to-face, remote video, pre-recorded)  Target population: 12–15-year-old boys participating in a rugby league development programme  Delivered by: Wellbeing officers with training in elite athlete well-being training, or registered psychologists. | Total n = 671  n =195 players completed the program including both the pre- and post-RISE Assess steps  n = 482 players completed the program and satisfaction ratings  **Demographics** (total sample including non-completers and healthy sample_  Age: M = 13.35, SD = 0.35  Gender: Male n = 671, (100%)  Ethnicity: Born in Australia: n = 624 (93%)  **Mental health problems:**  Total in high-risk range for anxiety or depression: n = 46;  Anxiety symptoms: n = 6;  Depressive symptoms: n = 7;  Anxiety and depression symptoms: n = 1;  Behavioural problems n = 19;  Anxiety and behavioural problems: n = 2;  Depression and behavioural problems: n = 7;  Anxiety, depression and behavioural problems: n = 4 | Yes – states in introduction that there have been over five years of research on this co-designed and co-delivered youth rugby league development program, and that it was developed based on community-based participatory research frameworks. | Pre-post single group study | 15 | RQ2:  Pre-post comparisons:  The 'high-risk range' group had significant reductions from pre- to post-RISE in anxiety (p < 0.001), depression (p < 0.001) and behavioural problem scores (p < 0.001).  There was no significant change in the 'health range' group pre- to post-RISE in anxiety (p = 0.74), depression (p = 0.56) or behavioural problem scores (p = 0.52).  RQ6:  Helpfulness ratings:  Mean ratings of helpfulness were between 3.5-3.81, and there was no significant difference in helpfulness ratings between delivery format groups.  Enjoyableness ratings:  Mean ratings of enjoyableness were between 3.4-3.9, and these significantly differed between delivery conditions (F(3, 478) = 3.23, p = 0.022, np2 = 0.02).  Post-hoc tests showed significantly higher enjoyment ratings in the 'in-person + remote real-time' condition (p = 0.045) and the 'remote delivery only' condition (p = 0.039), compared to the 'in-person delivery only' condition. No other comparisons were significant (all p > 0.125). |
| Name: Single-session family therapy programme (SST) | Hopkins et al. (2016)  Country: Australia | Explore the experience of single-session family therapy programme (SST) sessions and the impact of participation on the young persons’ functioning. | Setting: Community-based (face-to-face)  Target population: Young people presenting to a mental health service and their families or caregivers.  Delivered by: Headspace clinicians | Total n = 265  Follow up (4-5 weeks post-session):  ORS: n = 238  SRS: n = 265  **Demographics:** Not reported  **Mental health problems:** Not reported | No | Pre-post single group study | 4 | RQ3:  Pre-post comparisons:  Functioning: Both mothers and fathers rated the young-person’s functioning at 4-5 weeks post session as improved, with a Two-way ANOVA finding a significant time effect, F(1,40)=18.55, p<.001, but no interaction of role (p=.14).  RQ6:  Acceptability: There was an overall mean score of 33.75 (SD 5.73) out of a total score of 40 indicating that participants rated the sessions highly, although for 56% this was not at least a score of 36, indicating a possible source of concern. A significant main effect of role was also found, F(3,261)=11.17, p<.001. Tukey’s post-hoc tests found that mothers rated the sessions significantly higher than siblings/others (p=.015) and young people (p<.001). Fathers also rated the sessions as significantly higher than young people (p<.001). No other comparisons were significant. |
| Name: Single-session therapy (SST) | Kachor et al. (2020)  Country: Canada | To evaluate the implementation of a single-session therapy (SST) pilot project in a youth community-based mental health clinic | Setting: Community-based (face-to-face)  Target population: Youth aged 12-18 presenting with mild to moderate anxiety or depression, and their families.  Delivered by: SST-trained clinicians | n = 338 attending a single session therapy  n = 179 reached for follow-up phone call  **Demographics:** Not reported  **Mental health problems:** Not reported | No | Pre-post study with single group and comparisons to before implementation | 6 | RQ1:  Pre-post comparisons:  Wait time: Youth and families who agreed to attend a single session of therapy received service within 30 days. This resulted in a 90-day reduction in the wait times for the initial session of therapy compared to the previous 120-day wait.  RQ3:  Pre-post comparisons:  Functioning: Between the start of the single session therapy and one-month follow-up, families reported improved functioning (average improvement score of 9.59 using the Outcome Rating Scale). Any score greater than 6 is considered reliable change. 37% of post-test scores revealed clinically significant change.  RQ6:  Acceptability: 100% of families reached reported that the single session therapy had been helpful in some way. |
| Name: The Intensive Mobile Youth Outreach Service (IMYOS) team (a sub programme of ORYGEN Youth Health (OYH)) | Schley et al. (2008)  Country: Australia | Explore client characteristics and treatment effects in a group of difficult to engage, ‘high-risk’ young people, seen by the Intensive Mobile Youth Outreach Service (IMYOS, ORYGEN Youth Heath) in Western Metropolitan Melbourne. | Setting: Community-based (face-to-face)  Target population: Young people aged 15-24 who display signs of mental illness, are considered at ‘high-risk’ (e.g. of suicide) and have a history of poor engagement with clinic-based services.  Delivered by: The service is staffed by three psychologists, two social workers, one occupational therapist and one psychiatric nurse plus a consultant psychiatrist is available for 2 days a week. | n = 47  **Demographics:**  Overall age: M = 15.5  Females’ age: M = 14.9, SD = 1.39  Males:’ age: M = 15.0, SD = 2.5  Gender:  Females: n = 22 (47%) Males: n = 25 (53%)  **Mental health problems (n = 45):**  Mood disorders: 40%;  Attention-deficit and disruptive behaviour disorders: 37.8%; Substance-related disorders: 31.1%; Anxiety disorders: 22.2%;  Eating disorders: 8.9%;  Psychotic disorders: 8.9%;  Other: 17.8% | No | Pre-post single group study | 15 | RQ2:  Pre-post comparisons:  Psychiatric hospital admission rates: Significantly reduced at all measurement periods except the 12-month follow-up interval (reduced by 17.0% at 3-months, 29.4% at 6-months, 27.6% at 9-months, and at 12-months was still lower than the 12-months before but not statistically significantly so).  Time as inpatients: Significantly reduced at all measurement periods except the 12-month follow-up interval (clients spent on average 6.7 days less in hospital at 3-months, 6.2 days less at 6-months, 5.5 days less at 9-month post-referral, and spent less time as inpatients at 12-months post-referral but not statistically significantly so.  Suicidal ideation: Significantly reduced following IMYOS involvement (at referral: n=41, 90.2%; At discharge: n=28, 17.9%; McNemar test: n=24, p=0.000).  Deliberate self-harm: Significantly reduced following IMYOS involvement (at referral: n=36, 86.1%; At discharge: n=31, 6.5%; McNemar test: n=25, p=0.000).  Substance abuse: Did not significantly change between referral to and discharge from IMYOS (at referral: n=40, 77.5%; At discharge: n=30, 70%; McNemar test: n=29, p=0.453).  RQ3:  Pre-post comparisons:  Violence: Significantly reduced following IMYOS involvement (at referral: n=40, 85%; At discharge: n=14, 42.9%; McNemar test: n=14, p=0.016).  Crime: (at referral: n=42, 59.5%; At discharge: n=19, 26.3%; McNemar test: n=18, p=0.031). |
| Name: Voluntary and community sector counselling services for CYP (Youth Information Advice and Counselling Services [YIACS]). | Duncan et al. (2020)  Country: UK | 1. Conduct the first systematic evaluation of the outcomes of counselling in the voluntary and community sector in England. 2. Collect outcome data using a common set of measures in accordance with a pre-defined protocol and across a large number of participants at several voluntary and community sector sites. 3. Establish the demographic profile of service users accessing voluntary and community sector counselling services for young people and young adults. | Setting: Community-based (face-to-face)  Target population: Young people aged 11-25.  Delivered by: Counsellors who have completed a diploma in counselling or psychotherapy. | Total n = 2144 Included in analyses: n = 1241  **Demographics:**  Gender  Male: 638 (29.8%);  Female: 1384 (64.6%);  Transgender: 1 (0.0%);  Missing: 121 (5.6%)  Age:  Mean age: 18.2 years (SD = 3.5);  Aged 11-15: 508 (23.7%);  Aged 16-20: 957 (44.6%);  Aged 21-25: 570 (26.6%);  Missing: 109 (5.1%)  Ethnicity  White British: 1047 (48.8%);  White Other: 109 (5.1%);  Asian/Asian British: 126 (5.9%);  Mixed ethnicity: 176 (8.2%);  Black/Black British: 311 (14.5%);  Other ethnic background: 55 (2.6%);  Missing: 320 (14.9%)  **Mental health problems:**  The most prevalent presenting problem was generalized anxiety (n = 980, 67.8%), followed by depression/low mood (n = 875, 60.5%) | No | Pre-post single group study | 17 | RQ2:  Pre-post comparisons:  Psychological distress: There was a clinically significant reduction in psychological distress in both participants aged <17 (YP-CORE; t(451) = 13.6, p < .001, mean difference=5.2 (SD = 8.1, 95% CI: 4.4, 5.90), Cohen’s d=0.64) and aged 17+ (CORE-10; t(788) = 22.4, p <.001, mean difference= 6.5 (SD = 8.2, 95% CI: 5.96, 7.11) Cohen’s d=0.8). 37.2% and 52.2% of those aged <17 and 17+, respectively, showed reliable improvement. 36.4% and 29.4%, respectively, showed clinically significant change (recovery).    Predictors of RQ2:  Number of sessions**:** Number of sessions positively predicted post-therapy psychological distress on the CORE-10 (b = 0.15, 95% CI: 0.32, 0.13, p < .001) but not the YP-CORE.  Age: Age positively predicted post therapy psychological distress on the CORE-10 (b = .08, 95% CI: 0.45, 0.06, p = .011) and YP-CORE (b = .10, 95% CI: 0.09, 0.82, p = .015).  Gender: Gender positively predicted post therapy psychological distress only on the YP-CORE (b = 1.00, 95% CI: 0.28, 3.44, p = .021), males were more likely to show improvement or recovery. |
| Name: '@ease' Dutch youth-initiative | Boonstra et al. (2024)  Country: The Netherlands | To evaluate outcomes and support use in 12- to 25-year-old visitors of the @ease mental health walk-in centres, a Dutch initiative offering free counselling by trained and supervised peers. | Setting: Community  Target population: Young people aged 12-25  Delivered by: Trained peer counsellors, health care professional, psychiatrist | Total N= 754  **Demographics:**  Gender: Female n=382 (62%)  Age: Visited once M 19.98, SD. = 3.53, returned M 20.85, SD = 3.31  **Mental health problems:** NR | Peer counsellors with lived experience support delivery of the intervention | Pre-post single group study | 13 | RQ2: Pre-post comparison:  Psychological distress: of the 95 participants who answered the CORE-10 on the first and last visit (which typically occurred over 6 weeks and within 3 visits), 28.4% improved reliably (defined as a decline of at least 6 points on the scale), compared to 6.3% who deteriorated reliably (increase in 6 or more points) while 65.3 showed no reliable change. Changes mostly occurred over a short period of time as 49.1% made their last visit within 6 weeks of their first visit.  Social functioning: Among the 53 participants who had a SOFAS score at first and last visit, 39.6% improved reliably (10+ point increase) while 24.5% decreased reliably (10+ point decrease) and 35.8% did not change reliably.  RQ6:  Satisfaction with the @ease visit was rated 4.5 out of 5 on average; 59.8% were very satisfied, 33.5% were satisfied, 4.2% were neutral and 2.5% were not satisfied. |
| Name: ENYOY- Platform | Van Doorn et al. (2023)  Country: The Netherlands | To investigate the effects on young people using the ENYOY-platform in relation to psychological distress, psychosocial functioning, and positive health parameters. | Setting: Online  Target population: Young people aged 16 to 25 with emerging mental health complaints  Delivered by: Therapist, peer workers, clinical moderators | Total N = 131  **Demographics:**  Gender: Female 116 (88.5%); Male 14 (10.7%); Intersex 1 (0.8%)  Age: mean 21.61 (SD 2.2) years  Ethnicity: Dutch or of Dutch and other ethnic and racial backgrounds 117 (89.3%)  Surinamese 6 (4.6%)  Other 3 (2.3%)  Unknown 5 (3.8%)  **Mental health problems:** NR | Peer workers with lived experience support delivery of the intervention | Pre-post single group study | 16 | RQ2:  Pre-post comparisons:  Psychological distress: There was a significant improvement over time from baseline to 12 months (p<.001). This was due to significant improvements in K10 scores between baseline and 3 months (P<.001; d=0.62), and between 3 and 6 months (P<.001, d=0.37), while there was no significant difference between 6 and 12 months (p=0.54). 77/102 (75.5% )of cases with data showed clinically significant changes at 6 months, 63/82 (77%) at 12 months.  Functioning: There was a significant improvement over time from baseline to 12 months (p<.001). There were significant improvements in SOFAS scores between baseline and 3 months (P<.001; d=0.50), and between 3 and 6 months (P<.001, d=0.50). There was no significant difference between 6 and 12 months (p=0.21). 91/102 (89.2%) of cases with data showed clinically significant changes at 6 months, 75/82 (91%) at 12 months.  Quality of life: There was a significant improvement over time from baseline to 12 months (p<.001). There were significant improvements in QoL scores between baseline and 3 months (P=.01; d=0.38), and between 3 and 6 months (P=.02, d=0.23). There was no significant difference between 6 and 12 months (p=0.28).  Predictors of RQ2: Improvement in both psychological distress and social functioning was predicted at the 6-month measurement by higher psychological distress (P<.001) and lower social functioning (P=.04) at baseline, respectively. This means that both severity of mental health complaints and impairment of social functioning predicted greater improvement in young people. In addition, age (P=.24), sex (P=.89), educational level (P=.19), and clinical stage (P>.99) did not predict improvement. |
| Models aimed at providing targeted support for specific needs beyond anxiety or depression | | | | | | | | |
| Name: Self-Help for Alcohol/other Drug use and Depression for Young People (SHADEY) | Hides et al. (2010)  Country: Australia | To determine the outcomes of an integrated CBT intervention for co-occurring depression and substance misuse in young people presenting to a mental health setting. | Setting: Community-based (face-to-face)  Target population: Young people aged 15-25.  Delivered by: Not reported. | n = 60  **Demographics:**  Age: M = 20.7, SD = 2.7;  Gender: Female: n = 26 (43.3%);  Ethnicity not reported  **Mental health problems:**  Axis I disorders: Generalised Anxiety Disorder: n = 12 (20%);  Post-Traumatic Stress Disorder: n = 12 (20%);  Social phobia: n = 9 (15%);  Panic disorder: n = 6 (10%);  Obsessive Compulsive Disorder: n = 2 (3.3%);  Specific phobia: n = 2 (3.3%);  Substance use Disorders:  Cannabis: n = 32 (53.3%)  Alcohol: n = 38, (63.3%);  Amphetamines: n = 8 (13.3%)  Opiate: n = 4 (6.7%);  Hallucinogen (includes ecstasy): n = 2 (3.3%);  Inhalant, sedative, poly: n = 4 )6.7%);  Axis II disorders: Childhood conduct disorder: n = 21 (41.5%);  Antisocial personality disorder: n = 9 (17.0%);  Borderline personality disorder: n = 2 (3.8%) | No | Pre-post single group study | 17 | RQ2:  Pre-post comparisons:  Depression:  Diagnosis: At baseline, 100% of participants had a diagnosis of major depressive disorder. At 20 weeks, 17.3% (n=9) had a diagnosis and at 44 weeks, 16% (n=8) had a diagnosis. This meant that at 20 weeks, 82.7% (n=43) of cases had a full or partial remission of MDD, and this figure increased to 84.0% (n = 42) at 44 weeks.  Symptoms: mean scores (measured using HAM-D) at week 10, (M=12.8, SE=0.7), 20 (M= 10.5, SE=0.7), and 44 (M= 10.5, SE=0.7) were significantly lower than baseline (M=18.9, SE=0.6; all p<.001)  Anxiety:  Symptoms: mean scores (measured using HAM-A) at week 10 (M=11.2, SE=0.8); week 20 (M=9.5, SE=0.8); week 44 (M=10.1, SE=0.8) were all significantly lower than baseline (M=16.2, SE=0.7; all p<.001)  Substance use disorder:  Diagnosis: There was a significant reduction in the number of SUDs from baseline to week 20 (Wilcoxon z= −4.92, p<0.001) and week 44 (Wilcoxon z=−2.77, p<0.01), and reduction in current use (both p<.001).  Symptoms: There were no significant differences in the total days of alcohol and drug combined use (p=0.074) and total days of abstinence (p=0.060). There was a significant reduction on the AUDIT alcohol use measure at week 44 (p=.001) |
| Name: Housing Outreach Program-Collaboration (HOP-C) | Kidd et al. (2020)  Country: Canada | Describe the first RCT of Housing Outreach-Program-Collaboration (HOP-C), comparing its outcomes with the delivery of transitional case management alone | Setting: Community-based (face-to-face, texts, telephone calls)  Target population: Young people aged 16-26, residing within city limits, who had experienced at least 6 months (not necessarily consecutive) of homelessness, and had been housed in a stable arrangement (i.e., not a crisis shelter, not couch surfing) between 1 day and 1 year since their last homeless episode.  Delivered by: 2-3 peer support workers, 2 transitional case managers (one child and youth worker, one social worker), and a clinical psychologist (an expert in mindfulness-based interventions). | Total n = 65 Intervention group: n = 34  Control group: n = 31)  **Demographics:**  ***All participants:***  Age M = 21.75, SD = 2.07;  ***HOP-C group:***  Age:  M = 21.76, SD = 2.14;  Gender:  Female (n = 12, 35%);  Ethnicity:  White-North American: n = 5 (14%), Black-African: n = 3, (8%)  Black-Caribbean: n = 4 (11%); Mixed heritage: n = 4, (11%);  ***Control group:***  Age M = 21.74, SD = 2.03;   Gender:  Female: n = 14 (46%);  Ethnicity:  White-North American: n = 5 (16%);  Black-African: n = 5, (16%);  Black-Caribbean: n = 3 (10%);  Mixed heritage: n = 2 (6%)  **Mental health problems:** Not reported | Yes – the intervention involves peer support workers, but there was no description of people with lived experience being involved in the design of the intervention, or evaluation of it in this study. | RCT | 14 | RQ2:  Contemporaneous control comparisons:  At six month follow-up, participants who had received the HOP-C intervention were not significantly more likely to have improved their mental health (on any measure) compared to the control condition.  RQ3:  Contemporaneous control comparisons:  Housing: At 6 months follow-up, there was no difference in likelihood of gaining or sustaining housing between participants who received the intervention and participants in the control group  Employment & Education: At 6 months follow up, there was no statistical difference in the likelihood of employment or education.  RQ6:  Programme engagement: There was no significant difference between the number of case management contacts for treatment (M=36.55, range=5-212, SD=40.14) and control participants M=35, range=5-99, SD=27.6). |
| Name: The Primary Care Intervention for Post-traumatic stress disorder (PCIP) | Ng et al. (2023)  Country: USA | To assess the feasibility and acceptability of the Primary Care Intervention for Post-traumatic stress disorder (PCIP), an Integrated Behavioural Health Care treatment for post-traumatic stress disorder (PTSD) in adolescents. | Setting: Primary care (face-to-face)  Target population: Young people aged 12-26 presenting to primary care suspected of having PTSD symptoms.  Delivered by: Social workers providing integrated care. | n = 20 consented;  n = 19 completed pre-assessment;  n = 8 completed post-assessment  **Demographics:**  Of the 19 at pre-assessment:  Age: M = 19.32, SD = 2.11  Gender:  Female: n = 17;  Male: n = 2; Transgender: n = 3.  Ethnicity:  Black or African-American: n = 8 (40%);  Hispanic: 31.58% (n not reported).  **Mental health problems:**  All participants were diagnosed with at least one mental disorder and 7/19 (36.84%) participants were diagnosed with 2 or more comorbid mental health disorders.  PTSD: 68.4%;  Major depression: 31.58%;  Trauma and stress related disorder: 31.58%;  Anxiety: 15.79%;  ADHD: 5.26%. | No | Pre-post single group study | 12 | RQ2:  Pre-post comparisons:  Mental health symptoms: Despite the small sample size (n=8 at post-assessment, n=4 at post-assessment), exploratory analyses suggested that there was significant improvement in symptom scores of anxiety (measured using the RCADS); hedges g =0.68, p =0.02) and substance use (hedges g = 0.36, p =0.04) from pre to post, and depression symptoms (measured using the CES-D; hedges g =0.38, p =0.04) from pre to follow-up. In addition, there were trends in symptom improvement for re- experiencing (hedges g = 0.52, p = 0.09) and low mood (hedges g = 0.38, p = 0.08) symptoms.  PTSD: Of the eight participants who completed the post-assessment (seven of whom completed three PCIP sessions, and one completed two sessions) four had PTSD symptoms that showed clinical improvement: two improved without clinical significance, three worsened, three had severe PTSD symptoms at both baseline and post-assessment.  RQ6:  Satisfaction/Acceptability: Post-treatment qualitative interviews indicated that 11/14 (79%) found the intervention helpful. All 8 patients who participated in exit interviews said they were satisfied with the treatment and would recommend it to others. |
| Name: Screening, brief intervention, and referral to treatment (SBIRT) | Sterling et al. (2018)  Country: USA | To examine patient outcomes from a pragmatic trial of two modalities of delivering screening, brief intervention, and referral to treatment (SBIRT) and usual care (UC) in paediatric primary care. | Setting: Primary care (face-to-face)  Target population: Adolescents (aged 12-18) presenting to primary care with comorbid mental health and substance use difficulties  Delivered by: There were two intervention conditions in this study:   a) Paediatrician-only: Paediatricians were trained to assess substance use and depression using evidence-based screening tools, deliver brief interventions, and refer patients to speciality mental health treatment. b) Embedded behavioural clinician: Paediatricians were trained to assess and refer patients to an embedded behavioural clinician for further assessment, brief interventions, brief CBT, and referral to treatment, as well as providing brief CBT-based treatment and crisis management for substance use and mood problems. | Screened: n = 5183; Eligible for further assessment: n = 1,871;  Total n = 648  Analytical sample: *Paediatrician only:*  Paediatricians: n = 15 Patients: n = 224  *Behavioural clinician:*  Paediatricians: n = 12 Patients: n = 215  *Usual care:*  Paediatricians: n = 12 Patients: n = 209  **Demographics:** *Embedded behavioural clinician group:*  Age: M = 15.1, SD = 1.2,  Gender: Male: n = 98 (43.8%)  Ethnicity;  Asian: n = 21 (9.4%),  Black: n = 68 (30.4%), Hispanic: n = 60, (26.8%);  White: n = 63 (28.1%)l  Other/unknown: n = 12 (5.4%);  *Paediatrician-only group:*  Age: n = 15.2, SD = 1.2  Gender: Male n = 82 (38.1%)  Ethnicity:  Asian: n = 31 (14.4%);  Black: n = 71 (33.0%);  Hispanic: n = 39, (18.1%);  White: n = 55 (25.6%);  Other/unknown: n = 19 (8.8%);  *Usual care group:*  Age: n = 15.3, SD = 1.2  Gender: Male n = 110 (52.6%)  Ethnicity:  Asian: n = 19 (9.1%);  Black: n = 61 (29.2%);  Hispanic: n = 36 (17.2%);  White: n = 81 (38.8%);  Other/unknown: n = 12 (5.7%)  **Mental health problems:**   - Substance use symptoms: Among all patients, 11% endorsed substance use symptoms at the index visit only, 15% at the follow-up visit only and 48% at both visits; - Depression symptoms: 5% of patients endorsed mood symptoms only at the index visit, 29% endorsed at the follow-up visit only, 17% endorsed symptoms at both visits | No | RCT | 11 | RQ2:  Contemporaneous control comparisons:  Depression symptom endorsement at baseline only: (1) embedded behavioural clinician group: 6.7%; (2) paediatrician-only group: 3.3%; Endorsement at follow-up only: (1) embedded behavioural clinician group: 27.2%; (2) paediatrician-only group: 31.6%. Patients in the embedded BC arm had lower odds of endorsing depression symptoms than those in the paediatrician-only arm (AOR = .71, 95% CI = .50–.99); the paediatrician-only arm did not differ from UC (AOR = 1.02, 95% CI = .72–1.43).  Pre-post comparisons:  The odds of endorsing depression symptoms increased between visits for all patients (AOR = 3.53, 95% CI = 2.78–4.40).  Predictors of RQ2 outcomes:  *Ethnicity:* Participants who were Black or of other/unknown ethnicity had higher odds of endorsing depression symptoms over time than participants who were White  *Age: O*lder adolescents had lower odds of endorsing depression symptoms over time.  *Gender:* Males had lower odds of endorsing depression symptoms over time. |
| Name: Screening, brief intervention, and referral to treatment (SBIRT) | Parthasarathy et al. (2021)  Country: USA | Examine the relationship between access to a screening, brief intervention, and referral to treatment (SBIRT) and substance use, depression and medical diagnoses, and health services use at 1- and 3-years post-screening for adolescents. | Setting: Primary care (face-to-face)  Target population: Adolescents aged 12 to 18 years endorsing past-year substance use and recent mood symptoms during visits to a general paediatric clinic.  Delivered by: Paediatricians and behavioural clinicians. | n = 1,851  Adolescents with past-year substance use and recent mood problems: n = 289 Adolescents with either past-year substance use or recent mood problems: n = 1582  **Demographics:**  *Analytic sample (SBIRT):*  Age: M = 16.1, SD = 0.1’  Gender: Female n = 134 (72.4%)  Ethnicity:  White: n = 37 (20%); Black: n = 72 (38.9%);  Asian American: n = 21 (11.4%);  Hispanic: n = 42 (22.7%);  Other: n = 2 (1%)  *Analytic sample (Usual care):*  Age: M = 16.1, SD = 0.1  Gender:  Female n = 65 (62.5%)  Ethnicity:  White: n = 28 (26.9%);  Black: n = 39 (37.5%);  Asian American: n = 9 (8.7%);  Hispanic: n = 20 (19.2%);  Other: n = 3 (2.9%)  **Mental health problems:**  Comorbidity in depression diagnosis: n = 53 (18.3%)  Comorbidity in substance use disorder diagnosis: n = 8 (2.8%)  Comorbidity in any chronic condition: n = 52 (18.0%) | No | Cluster RCT | 17 | Subsample of Sterling 2018 sample  RQ2:  Contemporaneous control comparisons:  Emergency Department (ED) use was lower in the 1-year post screening but did not differ between the SBIRT and UC groups (5.9% vs 10.5% for SBIRT and UC, respectively; P = .15). However, 3-year post screening data revealed the SBIRT group had significantly fewer ED visits (rate ratio = 0.65; CI = 0.44–0.97) compared with the usual care group.  Depression: The SBIRT group had lower odds of depression diagnoses (OR = 0.31; CI = 0.11–0.87) than the usual care group at 1-year post screening after adjusting for patient characteristics. Similar results were observed at 3 years post screening, with the odds of a depression diagnosis (OR = 0.51; CI = 0.28–0.94) being almost 50% lower in the SBIRT group than in the usual care group.  Substance use: Prevalence of unadjusted substance use diagnoses were similar at the 1-year post-screening and did not differ between the 2 groups (1 year post screening: 2.2% vs 2.9% for SBIRT and usual care, respectively; P = .70). Prevalence of substance use diagnoses increased to 11.9% for SBIRT and 20% for UC by 3 years, but the difference was not statistically significant (P = .06). |
| Name: Trauma Systems Therapy (TST) | Saxe et al. (2012)  Country: USA | Obtain preliminary evidence for the extent to which a novel intervention embedded within a systems-oriented treatment model – Trauma Systems Therapy (TST) – engages and retains traumatised children and their families in treatment. | Setting: Child psychiatry clinic of a large, urban hospital (face-to-face)  Target population: Young people with prominent symptoms of PTSD and their families.  Delivered by: A multidisciplinary team | Total n = 20  TST: n = 10  Care as usual (CAU): n = 10  **Demographics:**  Age: M = 13.7 (SD = 3.6)  Gender: Female n = 11 (55%)  Ethnicity:  Black: 68% Hispanic: 18% White: 10% Mixed race/ethnicity: 4%  **Mental health problems:**  PTSD: 100% | No | Quasi-experimental study | 14 | RQ2:  Pre-post comparisons:  In the TST group, there was a significant reduction in arousal symptoms (PTSD-RI Criterion D subscale scores; t = 2.65; p = 0.04) and aggression (Child Behaviour Checklist aggressive behaviour subscale; t = 2.85; p = 0.03), and improvement on the home safety subscale of the Child Assessment of Needs and Strengths-Trauma Exposure and Adaptation Version approached significance (t = 2.00; p = 0.08).  There were no other significant within-group differences (outcomes analysed unclear).  RQ6:  Retention: At the 3-month reassessment, 9 of 10 (90%) patients receiving TST were still enrolled in treatment compared with only 1 of 10 (10%) patients in the CAU condition. |
| Name: Integrated family-based outpatient treatment for adolescents (OPT-A). | Sheidow et al. (2021)  Country: USA | Examine the difference between an experimental treatment (OutPatient Treatment for Adolescents; OPT-A) to treatment as usual (TAU) through 18-months post-referral, and to investigate improvements in outcomes. | Setting: Community-based (face-to-face)  Target population: Young people aged 10-17 requiring treatment for current comorbid substance use and internalising (mood or anxiety) disorders.  Delivered by: Master's level clinicians, trained therapists and physicians (for medication prescriptions). | n = 134 youth/families  **Demographics:** Age: M = 16.0, SD = 1.1  Gender: Female = 37.9%;  Ethnicity:  White: 76.4%; African American: 15.7%; More than one race: 5.7%;  Not reported: 2.1%;  Hispanic: 2.1%.  **Mental health problems:**  Participants had comorbid substance use and internalizing (mood or anxiety) disorders, although baseline measures of internalising symptoms suggested that only 17% reached borderline level. | No | RCT | 17 | RQ2:  Contemporaneous control:  Substance use (controlled for sex and age at each time point):  From baseline to month 3, there was a significant between-group difference in the amount substance use changed over time, with OPT-A decreasing to 29% and TAU increasing to 56% (OR = 0.25; 95% CI: 0.08, 0.75).  There was no significant between-group difference in change in substance use between baseline and month 6 (OR = 0.49; 95% CI: 0.14, 1.64) and baseline and month 12 (OR = 0.26, 95% CI: 0.07, 1.00).  From baseline to month 18, substance use increased in both groups but the levels of substance use increased significantly less in the OPT-A group compared to the TAU group (52% vs 81%; OR = 0.19; 95% CI: 0.04, 0.96)  RQ3  Contemporaneous control comparisons:  School enrolment: At baseline, and from baseline to later assessments (3-, 6-, 12- and 18-month follow-ups), OPT-A and TAU did not differ on the log-odds of school enrolment  RQ6:  Contemporaneous control comparisons:  Parental satisfaction: At month 1, OPT-A had significantly higher satisfaction compared to TAU (3.82 vs 3.64), and the groups did not differ in change over time.  Treatment motivation: From baseline to month 1, OPT-A and TAU did not differ significantly in treatment motivation. From baseline to months 2, OPT-A increased more (to 2.26 for parents; 2.11 for youth) than TAU (to 1.99 for parents; to 1.63 for youth). From baseline to month 3, OPT-A had less of a decrease in treatment motivation (to 2.22 for parents) than did TAU (to 1.90 for parents). Both groups did not differ at baseline or in change from baseline to later assessments in motivation to cut down substance use. |
| Name: Multidimensional Treatment Foster Care adapted for girls with co-occurring trauma and delinquency (MTFC+T) | Smith et al. (2012)  Country: USA | Present a theoretical rationale for adapting a community-based intervention - Multidimensional Treatment Foster Care - to treat adolescent girls with co-occurring trauma and delinquency, describe the intervention approach, and present outcomes from a small-scale pilot study to show its impact on mental health outcomes and delinquency. | Setting: Community-based (face-to-face)  Target population: Females, ages 12–17, at least one arrest in the year prior to referral, court mandated for out-of-home placement, at least one traumatic experience, and not currently pregnant.  Delivered by: Trained and supervised foster parents (who had undergone 20 hours of pre-service training), programme supervisors. | Total n = 30; Intervention (MTFC + Trauma) group: n = 13;  Control (TAU): n = 17  **Demographics:** Age: M = 15.31, SD 1.29);  Gender: Female: 100% Ethnicity:  European American: 71% African-American: 4% Native American: 7% Latino: 7% Multiracial: 11%  **Mental health problems:** All had at least one traumatic experience | No | RCT | 13 | RQ2:  Contemporaneous control:  Girls taking part in the community fostering intervention had significantly fewer trauma-related mental health symptoms compared to the usual group (β = −.48, p < .05) at 12-months post-baseline.  Predictors of RQ2 outcomes:  *Baseline trauma* (β = .57, p < .05) and *prebaseline* *arrests* (β =−.49, p <.05) were significant predictors of trauma-related mental health symptoms at 12-month post-baseline.  There was no additive effect of *age or baseline mental health symptoms*.  RQ3:  Contemporaneous control:  Delinquency:  Girls taking part in the community fostering intervention had significantly lower levels of delinquency compared to TAU (β = −.44, p < .05) at 12-months post-baseline.  Predictors of RQ3 outcomes:  There was a trend for baseline delinquency to predict delinquency at 12 months postbaseline (β = .33, p < .10). There was no additive effect of age or baseline trauma. |
| Name: Multidisciplinary care provided in a drop-in centre | Souza et al. (2011)  Country: Honduras | Describe the uptake and outcomes of a multidisciplinary case management approach for street children and youth attending a drop-in day care centre in Tegucigalpa, Honduras. | Setting: Community-based (face-to-face)  Target population: Youth, specifically homeless children, under 25 years old  Delivered by: Trained clinical psychologists, social workers, and an education team. | Total n = 800; N  Contacted psychological service: n = 400 No contact with psychological service: n = 400  **Demographics:**  Age: M = 17.5, 95% CI: 17.0–17.9, SD not reported  Gender: Female (95% CI): 38.0 % (33.2–42.8) Ethnicity: Not reported  **Mental health problems:**  No specific diagnosis, but clients contacted services for medical or psychological consultations:   - 219 (54.8%) individuals reported feeling depressed and hopeless; - 312 (78%) reported anxiety and tension; - 24 (6%) reported having suicidal ideas - 5 (1%) reported having made a suicide attempt in the month prior to the first consultation; - 110 (25.3%) reported having trouble controlling violent behaviour. | No | Pre-post single group study | 20 | RQ2:  Pre-post comparisons:  Psychological distress: Between the first and last visit, there were significant reductions in symptoms of psychological distress (mean difference = -0.44, p = 0.0001).  The estimated reduction in psychological scores per year was -0.45 (95% CI: 0.23–0.67) for males and -1.00 for female patients (95% CI: 0.76–1.24).  Substance use: Between the first and last visit, there was a significant increase in days without substance use (mean difference = -2.21, p <= 0.0001).  - In 236 patients who had baseline and follow-up visits, 114 (48%) reported using substances consistently throughout both periods, 19 (8%) reported never to have used substances; 78 (33%) intermittently took substances during follow-up and 19 stopped substance use (8%). A further six (3%), who reported no substance taking at baseline, started taking substances during follow-up.  - Survival analysis showed that the probability of remaining on substances was 0.76 (95% CI: 0.69-0.81) and 0.51 (95% CI: 0.42-0.59) at 24-months.  Predictors of RQ2 outcome:  Age did not predict improvements in psychological distress however there was a significant main effect of gender (p < 0.001) and a significant interaction of gender over time (p < 0.001) on improvements in levels of psychological distress.  At 12-months, significantly fewer female patients remained using substances compared to male (p < 0.01).  RQ3:  Pre-post comparison:  122/373 participants reported improvements in their social situation (sleeping arrangements), compared to 174 who reported no change, 77 who reported negative changes. |
| Name: Individual Placement and Support (IPS) added to HEADSPACE | Telford et al. (2024)  Country: Australia | To investigate the effectiveness of the headspace IPS program in achieving mental health outcomes in comparison to those achieved with mental health treatment alone. | Setting: Community  Target population: Young people aged 15-25 years old | Total n = 2128; IPS: 544 Non-IPS: 1584  **Demographics:**  Age:  IPS group: mean 19.7; SD 2.7  Non-IPS group: mean 19.4; SD 2.7  Gender:  IPS group: Female n= 346; 63.6%; Male n=182 33.5%; Gender diverse n=16 2.9%  Non-IPS group Female n= 980 61.9%; Male n=555 35%; Gender diverse n=49 3.1%  Ethnicity: (indigenous population):  IPS group: n= 52; 9.6%  Non-IPS group: n=92; 5.8%  **Mental health problems:** NR | No | Matched cohort study | 17 | RQ2:  Contemporaneous control comparisons:  Quality of life: IPS was associated with significant improvements (greater than at least half a standard deviation of the respective measure’s baseline for the entire headspace population in the sampling period) in quality of life (58.3% vs 50.3%; OR=1.24, p=.043).  Psychosocial functioning and distress: Although a higher percentage of participants who received IPS improved in psychosocial functioning (53.9 vs 46.1%) and psychological distress (39 vs 37.4%), when controlling for matching variables and baseline severity, IPS was not associated with a significantly increased odds of improvement (OR: 1.24 and 1.04 for psychosocial functioning and psychological distress, respectively; p>0.05).  Predictors of RQ2:  Males were more likely to see improvement in quality of life than those not receiving IPS, although females were no more likely.  Although underpowered, Aboriginal or Torres strait Islander young people had a 47% to 64% higher likelihood of improving significantly on at least one mental health outcome compared to non-IPS counterparts.  RQ3:  Contemporaneous control comparisons:  At the conclusion of their IPS episode, the vocational outcome achieved for almost two thirds (65%) of IPS clients was that they had obtained a job (56%) or commenced study (9%). An additional almost 8% had other positive work or study outcomes recorded. Just over one quarter did not achieve a work and study outcome. For the non-IPS clients, 12% reported a positive change in their work and study status. |
