## Appendix 5 for "Approaches to early intervention for common mental health problems in young people: a systematic review"

### Appendix 5a: Associations with outcome by variable

| **High-level factor** | **Factor** | **Results** |
| --- | --- | --- |
| Demographics | Ethnicity | The Youth Partners in Care QI intervention (Ngo, 2009; Rapp, 2017) suggested that when comparing sub-groups by ethnicity, more participants from minority ethnic groups accessed mental health treatment through the QI programme compared to usual care, particularly through improved access to counselling or improved speciality mental health care for Black and Latino participants, and that Black participants experienced significantly larger improvements in mental health compared to Black participants in the usual care condition. This difference was not present for Latino or White participants. Khasanov (2024) also reported that in participants receiving the collaborative care model (CoCM), declines in depression and anxiety were higher among individuals identifying as Black (4.54 mean difference from pre-to post-) compared to White (4.06 mean difference) participants. In contrast, Sterling et al. (2018) reported that SBIRT participants who were Black or of other unknown ethnicity had higher odds of endorsing depression symptoms over time compared to White participants. Although underpowered, Aboriginal or Torres strait Islander young people had a 47% to 64% higher likelihood of improving significantly on at least one mental health outcome compared to non-IPS counterparts suggesting a particular benefit for this group when IPS is added to HEADSPACE early intervention (Telford, 2024). |
|  | Age and Gender | Most studies reported that age and gender had limited impact on mental health and social outcomes (O’Keeffe, 2015; O’Reilly, 2022; Smith, 2012; Schley, 2019; Souza, 2011; Van Doorn 2023), although Sterling et al. (2018) reported that younger or female adolescents were more likely to report depression symptoms after the screening, brief intervention and referral intervention for comorbid substance use compared to older or male participants, and the Youth Partners in Care QI intervention appeared to mitigate the negative effect of age on treatment access (Rapp, 2017). Khasanov (2024) also reported marginally higher pre- to post- declines in depression for females (4.36 mean difference) compared to males (4.02), while conversely males were more likely to see quality of life improvements than females following IPS support integrated with headspace (Telford, 2024). |
|  | English as first language | Access rates were only higher for those whose primary language was English in the Youth Partners in Care QI intervention (Rapp, 2017). |
|  | Living situation and location | One study reported that living situation was not a predictor of improvement in mental health outcomes (Corley, 2023). Furthermore, rural compared to urban intervention sites also did not demonstrate significant differences in effects, although participants in urban sites rated participant in the RISE rugby league as more enjoyable than those in the rural sites (Waters, 2024). |
| Severity | Baseline mental health symptoms | Neither baseline mental health symptoms nor diagnosis were reported to predict mental health symptoms post intervention in two studies (Smith, 2012; Parry, 2023), although another online intervention produced greater improvements in psychological distress and social functioning improvements in those with higher severity at baseline (Van Doorn 2023), and one study reported that complexity of presenting problems interacted with age such that this was only a significant predictor in participants aged 17+ (Corley, 2023). However, attendance at diagnosis-specific interventions did predict more positive mental health outcomes (Bond, 2020). Medication prescription and the presence of a neurodevelopmental disorder were reported as significant predictors of drop out in one study (Bond and Power, 2020). |
| Service-related | Number of interventions provided or sessions completed | Although time spent in the service was a significant predictor of drop out in one study (Bond and Power, 2020), the number of completed sessions or interventions given did not predict mental health or functioning outcomes of interventions (Schley, 2019; Corley, 2023; Parry, 2023). However, Khasanov (2024) reported in a pre- to post- comparison that patients receiving a collaborative care model for longer periods of time showed significantly greater declines in depression severity. |
|  | Referral route | Referral route was not reported to influence mental health outcomes (Corley, 2023; Parry, 2023), although referral from CAMHS (Bond and Power, 2020) was reported as a significant predictor of dropout. Another study found that being referred by schools was associated with a significantly higher likelihood of reliable deterioration in goal-based outcome scores (O’Reilly, 2022). |
|  | Level and format of intervention access | Full (including peer-to-peer web-based social networking chat for those assessed as safe to do so) versus partial access to a web-based intervention did not predict mental health symptoms. However, those with full access had improved loneliness, social support, and autonomy, while those with only partial access had improved self-competency alongside social support (Alvarez-Jimenez et al., 2020). Participants who received ‘remote’ or ‘in-person and remote real-time' sessions (versus in-person delivery only) rated RISE Rugby League sessions as significantly more enjoyable (Waters, 2024). |

### Appendix 5b: Associations with outcome by outcome

Associations with access and waiting time outcomes

Factors associated with access and waiting time outcomes were examined in two publications from one RCT evaluating the Youth Partners in Care QI intervention (Ngo, 2009; Rapp, 2017). One reported that at six months (when most outcomes of the QI intervention were favourable compared to the usual care group), more participants from minority ethnic groups accessed mental health treatment through the QI programme compared to usual care, including increased odds of counselling and speciality mental health care for Black and Latino participants. However, these groups were not more likely to access mental health care through a primary care clinician or medication (Ngo, 2009). The intervention also mitigated the negative effect of age on treatment access (Rapp, 2017), although access rates were only higher for those whose primary language was English (Rapp, 2017).

Factors associated with mental health and wellbeing outcomes:

Fifteen studies in total examined factors associated with mental health and wellbeing outcomes (Alvarez-Jimenez et al., 2020; Bond, 2020; Corley, 2023; Dowell 2020; Ngo, 2009; O’Keeffe, 2015; Parry, 2023; Schley 2019; Smith, 2012; Sterling, 2018; Souza 2011; Van doorn 2023; Telford 2024; Khasanov 2024).

Three RCTs and one matched cohort study with contemporaneous controls examined factors associated with mental health and wellbeing outcomes. These included one study examining a model designed to make care more comprehensive and joined up (Youth Partners in Care QI intervention; Ngo, 2009) and three evaluating models aiming to provide targeted support for specific needs beyond anxiety or depression (SBIRT: Sterling, 2018; MTFC+T: Smith, 2012; IPS integrated with HEADSPACE; Telford 2024). Ngo et al. (2009) found that the Youth Partners in Care QI intervention was predominantly beneficial for Black participants, who experienced significantly larger improvements in mental health compared to Black participants in the usual care condition, while Latino and White participants did not (Ngo, 2009). Telford (2024) also reported that although the finding did not reach statistical significance, those of aboriginal ethnicity in Australia appeared to benefit more from an IPS intervention integrated into early intervention. In contrast, Sterling et al. (2018) reported that SBIRT participants who were Black or of other unknown ethnicity had higher odds of endorsing depression symptoms over time compared to White participants. Additionally, younger adolescents and females were more likely to report depression symptoms than older or male participants (Sterling, 2018) and males benefited more compared to controls than females from IPS support (Telford 2024). Smith et al. (2012) identified baseline trauma and pre-baseline arrests as significant predictors of trauma-related symptoms at 12 months post-baseline for MTFC+T participants, while age and baseline mental health symptoms did not have an additive effect (Smith, 2012).

Nine pre-post studies explored factors associated with mental health and wellbeing outcomes of models aiming to make care more comprehensive and joined up (Bond, 2020; Corley, 2023; O’Keeffe, 2015; Parry, 2023; Khasanov 2024), models increasing speed or ease of access to care (Alvarez-Jimenez, 2020; Dowell, 2020; Schley, 2019; Van Doorn 2023), and models providing targeted support for specific needs beyond anxiety or depression (Souza, 2011). Living situation (Corley, 2023), referral route (Corley, 2023), diagnosis (Parry, 2023), urban versus rural location (Dowell, 2020), full versus partial intervention access (Alvarez-Jimenez, 2020), and age (O’Keeffe, 2015; Schley, 2019; Souza, 2011; Van Doorn 2023) did not significantly predict mental health outcomes. There were mixed results regarding ethnicity and gender (O’Keeffe, 2015; Schley, 2019; Souza, 2011, Khasanov 2024) and intensity of services received (Number of interventions received did not predict outcomes in one study (Parry, 2023) but time in service did in another (Khasanov 2024)).. However, attendance at diagnosis-specific interventions did predict more positive mental health outcomes (Bond, 2020), as did complexity of presenting problems in participants aged 17+ (Corley, 2023) and severity of psychological distress and psychosocial functioning at baseline (Van Doorn 2023).

Factors associated with social or functioning outcomes

Factors associated with social or functioning outcomes were explored in two pre-post studies evaluating approaches aiming to increase speed or ease of access to care (Alvarez-Jimenez, 2020; Schley, 2019) and one approach aiming to make care more comprehensive and joined up (O’Reilly, 2022). One reported that age group, gender, centre, number of completed sessions, and dropout versus planned treatment completion status were not significant predictors of occupational and social functioning (Schley, 2019). Likewise, O’Reilly et al. (2022) found no significant association between changes in goal-based outcome scores and either age group or gender. However, young people referred by schools were significantly more likely to experience reliable deterioration in goal-based outcomes than those referred by other sources (O’Reilly, 2022). Alvarez-Jimenez et al. (2020) found that participants with full intervention access only showed improvements in loneliness, social support, and autonomy, while those given partial access only showed significant improvements in social support and self-competency.

Factors associated with acceptability outcomes

Three pre-post studies evaluated factors associated with acceptability, including one study evaluating a model aiming to make care more comprehensive and joined up (YAS: Bond & Power, 2020) and two studies evaluating a model aiming to increase speed or ease of access to care (RISE: Dowell, 2020; Waters, 2024). Significant predictors of YAS dropout rates included: time in the service, referral from CAMHS, medication prescription, and presence of developmental disorder (Bond & Power, 2020). RISE participants at urban sites (versus rural) and receiving ‘remote’ or ‘in-person and remote real-time' sessions (versus in-person delivery only) rated sessions as significantly more enjoyable (Waters, 2024). There were no differences in ratings of helpfulness, usefulness, or ease of understanding between participants at rural versus urban sites (Waters, 2024).
